# Hierarchical genetic regulatory architecture of the human immune system informs complex disease mechanisms

**DOI:** 10.64898/2026.11.17.26366583

**Authors:** Nicola Pirastu, Sodbo Sharapov, Danai Vagiaki, Eva Holtkamp, Arianna Landini, Matiss Ozols, Edoardo Giacopuzzi, Manuel Tardáguila Sancho, Paola Benaglio, Alessandro Raveane, Stephen Watt, Bruno Ariano, Olli Dufva, Jan P. Engelmann, Daniela Zanotti, Giuditta Clerici, Davide Bolognini, Lucas Arnoldt, Beatrice Costa, Shuang Li, Andrei Cristea, Eljas Roellin, Ruth Eberhardt, Andrea Senacheribbe, Dominique Von Schiller, Eve Coomber, May Hu, Gabriella Ivanova, Fereshteh Torabi, Nima Subasinghe, Laura Durrant, Vahid Shariati, Chiara Chiereghin, Michela Colombo, Ilaria Gori, Matilde I. Conte, Emanuele Saitto, Federica Santonastaso, Cecilia Dominguez Conde, Blagoje Soskic, Alice Giustacchini, Craig A. Glastonbury, Carl A. Anderson, Genes & Health Research Team, Nicole Staudt, John Wright, Richard Trembath, Julien Gagneur, Sarah Finer, Fabian J. Theis, Francesco Paolo Casale, Fabiola Curion, David A. van Heel, Vivek Iyer, Brian Clarke, Sarah A. Teichmann, Oliver Stegle, Marc Jan Bonder, Nicole Soranzo

**Author notes:** Correspondence to be addressed to NS, Fondazione Human Technopole, Via Levi Montalcini 1, 20157 Milano, Italy. MJB, University Medical Center Groningen, Groningen, Netherlands. NP, Fondazione Human Technopole, Via Levi Montalcini 1, 20157 Milano, Italy. SAT, Department of Medicine, Cambridge Stem Cell Institute, University of Cambridge, Cambridge, UK. OS., Division of Computational Genomics and Systems Genetics German Cancer Research Center (DKFZ) Heidelberg, Germany. These authors contributed equally to the manuscript (Nicola Pirastu, Sodbo Sharapov, Danai Vagiaki, Eva Holtkamp, Arianna Landini, Matiss Ozols, Edoardo Giacopuzzi, Manuel Tardáguila Sancho, Paola Benaglio). These authors contributed equally to the manuscript (Nicola Pirastu, Vivek Iyer, Brian Clarke, Sarah A. Teichmann, Oliver Stegle, Marc Jan Bonder, Nicole Soranzo).

## Abstract

How inherited genetic variation propagates through multicellular regulatory systems to influence human disease remains poorly understood. To reconstruct the regulatory architecture of genetic variation on circulating immune cells, we analysed single-cell transcriptomes from 6.7 million peripheral blood mononuclear cells from 4,452 European-ancestry UK Biobank participants, identifying 114,063 independent cis-regulatory effects affecting 15,419 genes. Using a new statistical workflow (FLANDERS), we map the hierarchical organisation of these effects and the underlying regulatory architecture across tissues, immune lineages, cell types and cell states. We find that disease-associated variants predominantly act through regulatory programmes shared within immune lineages and broad cellular compartments, rather than through highly specific cellular states. We show that local *cis*-regulatory effects commonly act through coherent regulatory pathways implicating transcription factor cascades and intercellular signalling, which allows linking together multiple disease genes. We demonstrate independent support for these genes and mechanisms through rare variant associations. Our findings reveal a highly organised structure of immune regulatory variation shaping human disease, and provide a highly effective framework for interpreting disease associations and prioritising therapeutic targets.

## Introduction

Complex traits emerge through the coordinated activity of multicellular systems, yet how inherited genetic variation propagates through these systems to influence human phenotypes remains poorly understood ^1–3^. Genome-wide association studies (GWAS) have identified hundreds of thousands of variants associated with human traits and diseases, showing that genetic variation is a fundamental determinant of phenotypic diversity ^4^. However, translating these associations into biological mechanisms remains a major challenge because most variants reside outside protein-coding sequences and are thought to act by altering gene regulation ^5^. Understanding how local regulatory perturbations are transmitted across cells and biological pathways to shape organismal phenotypes is currently a central goal of human genetics.

Expression quantitative trait locus (eQTL) studies have established altered gene expression as a fundamental mechanism through which common genetic variation influences complex traits ^6,7^. By linking regulatory variants to nearby genes, these studies have substantially improved interpretation of disease-associated loci and have accelerated the discovery of candidate therapeutic targets ^8–10^. However, most regulatory effects have been characterised in bulk tissues, where measurements represent averages across heterogeneous cellular populations ^11–13^. As a consequence, it remains poorly characterised how regulatory effects are distributed across the cellular diversity of human tissues, whether they act broadly across related cell populations or are restricted to specialised cellular states, and at what biological scale genetic variation influences disease.

Single-cell genomics has transformed the study of gene regulation by enabling genetic effects to be resolved at cellular resolution ^14,15^. Single-cell eQTL studies at population scale have revealed extensive cell-type-specific and context-dependent genetic regulation of gene expression.^15,16^. By linking GWAS loci to regulatory effects that are restricted to specific immune cell types or activation states, these studies provided novel insights into the cellular contexts and molecular mechanisms underlying disease associations ^17,18^. These findings have provided increasingly precise maps linking genetic variation to specific immune populations. Although the sharing of these effects between cell types has been explored, they are limited to pairwise relationships and do not fully explore pleiotropy ^19–23^. Whether genetic regulation instead follows broader organisational principles across tissues, developmental lineages and cellular states, and how local regulatory perturbations propagate beyond individual genes to coordinated multicellular processes, remain of great importance for the interpretation of their disease context and thus their translational value.

Addressing these questions requires considering two complementary dimensions of genetic regulation. First, how regulatory effects are shared across tissues, immune lineages, cell types and cellular states. Second, how local cis-regulatory effects relate to broader transcriptional programmes operating within and across cell populations. Understanding both dimensions may help connect disease-associated variants not only to their target genes and cellular contexts, but also to the broader regulatory processes through which they influence disease.

The human immune system provides an ideal model for addressing these questions. Circulating immune cells comprise a rich hierarchy of lineages, specialised cell types and dynamic functional states that continuously respond to environmental exposures, infection and ageing ^24,25^. Because immune dysregulation contributes to the pathogenesis of numerous chronic inflammatory, autoimmune, metabolic and cardiovascular diseases, peripheral blood mononuclear cells (PBMCs) represent an accessible system in which to study how inherited variation is translated into disease through multicellular regulatory processes ^19–23^, particularly those driven by immune dysfunction ^10^. Recent advances in population-scale single-cell transcriptomics have made it possible to characterise these processes across thousands of individuals and millions of cells, enabling regulatory architecture to be examined across multiple levels of biological organisation.

Here we integrate population-scale single-cell transcriptomes from 6.7 million PBMCs across 4,452 UK Biobank participants of European ancestry with a newly developed statistical genetics framework to reconstruct the regulatory architecture of human peripheral immune cells. We identify nearly 114,063 independent *cis*-regulatory effects affecting 15,419 genes and show that regulatory variation is organised across successive biological scales, from multi-tissue regulation to lineage-, cell-type-and cell-state-specific programmes. We show that disease-associated variants predominantly act through regulatory programmes shared across related immune cell populations rather than highly restricted cellular states. Further, using single-cell deep learning models, we show that local *cis*-regulatory perturbations propagate through transcription factor networks and intercellular signalling to shape multicellular gene programmes, connecting independent disease loci to coherent regulatory pathways. Finally, we show that PBMC *cis*-eQTLs can be used to improve therapeutic target prioritisation and are independently supported by rare variant associations. Together, our findings establish a framework for understanding how inherited variation is organised across multicellular regulatory systems to shape human disease.

## Results

### Hierarchical organisation of human *cis*-regulatory variation across immune cell types

To define how genetic variation shapes transcription across the human immune system, we used *cis*-expression quantitative trait loci (*cis*-eQTLs) from the CARDINAL UKB dataset, comprising approximately 6.7 million peripheral blood mononuclear cells collected from 4,452 UK Biobank (UKB) participants as described in the cohort reference manuscript ^26^. We focus here on participants of European ancestry to facilitate comparison with well powered disease GWAS datasets. The dataset consists of 88,513 *cis*-eQTLs (i.e., gene–cell-type pairs having at least one associated genetic variant within a +/-1Mb of the gene’s transcription start site-TSS) affecting 15,419 genes across 23 immune cell types (**Fig. 1**, **Fig. 2a**).

**Figure 1.**
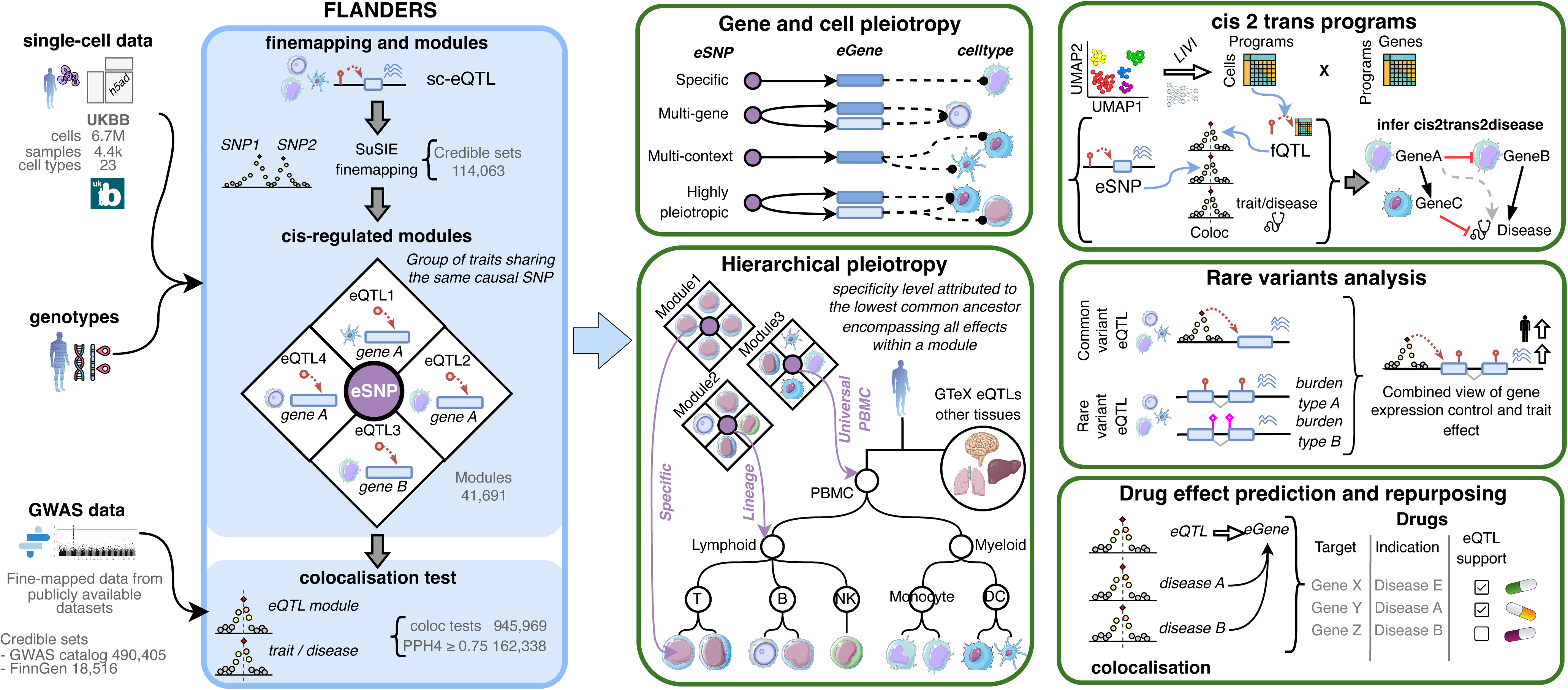
Reconstruction of the hierarchical regulatory architecture linking genetic variation to human disease. Overview of the analytical framework integrating population-scale single-cell transcriptomes from UK Biobank (UKB) with genotype data. The FLANDERS framework performs statistical fine-mapping, groups gene–cell-type associations driven by shared causal variants into cis-regulatory modules, and systematically tests colocalisation with disease-associated loci. We derived classification of fine-mapped regulatory variants according to target genes numbers and immune cell types, within and across biological scales. FLANDERS was also used to integrate fine-mapped cis-regulatory variants with LIVI multicellular gene programmes to identify cis-to-trans regulatory relationships. Rare variant burden analyses were integrated with common-variant regulatory associations to identify constrained genes inaccessible to conventional eQTL mapping and to provide independent validation of disease mechanisms inferred from common eSNPs. Integration with disease and with genetically inferred direction of effect was used to prioritise therapeutic targets, predict pharmacological direction, identify opportunities for therapeutic repositioning and anticipate potential adverse effects.

**Figure 2.**
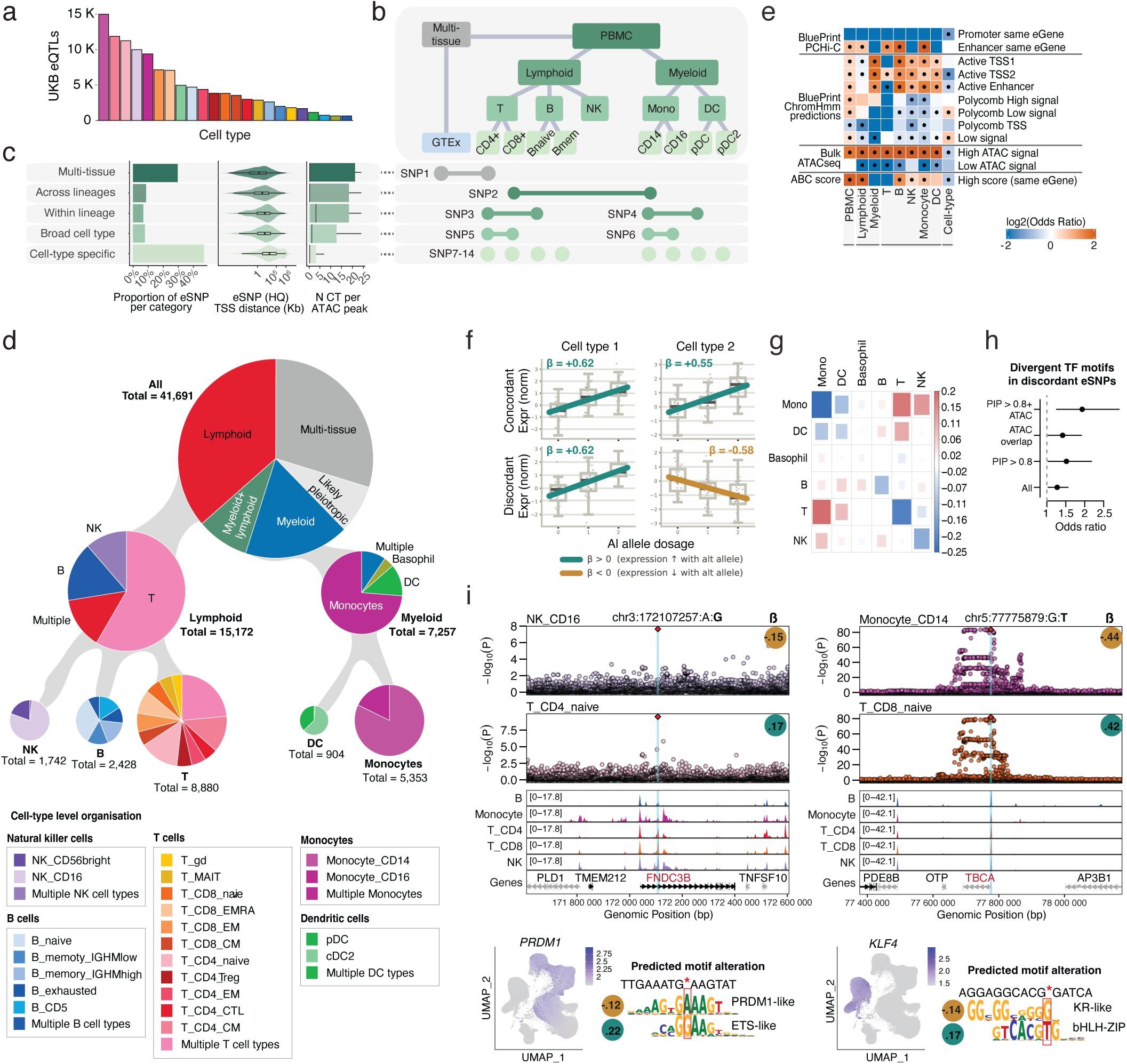
Hierarchical organisation of human cis-regulatory variation across the immune system. a, Number of discovered eQTLs. **b,** Hierarchical deployment of regulatory variation across multi-tissue, lineage-, cell-type- and cell-state-specific programmes, each with distinct genomic architectures. **c,** Properties of eSNPs across hierarchical classes: class proportions (left), distances from target-gene transcription start sites (TSS; centre) and sharing of accessible chromatin peaks across cell types (right). **d,** Distribution of PBMC-restricted eSNPs across biological compartments. Most are confined to lymphoid or myeloid lineages, with few shared between them. **e,** Enrichment or depletion (log₂ odds ratio) of promoter interactions, ChromHMM states, chromatin accessibility and activity-by-contact scores across pleiotropy classes. Black circles indicate FDR < 0.05. Broadly shared eQTLs favour promoter-associated, constitutively active elements, whereas specific variants favour distal, context-dependent chromatin. **f,** Illustrative directionally concordant and discordant eQTLs. **g,** Deviation from the expected probability of opposite allelic effects between pleiotropy levels. Discordance occurs preferentially between lymphoid and myeloid lineages and is uncommon between related cell types. **h,** Enrichment of discordant motif predictions among switching variants under increasingly stringent fine-mapping and chromatin-accessibility criteria. **i,** Regional association plots for two discordant eQTLs with opposing motif-alteration mechanisms. Effect sizes are shown for the indicated effect allele (bold) in two cell types. UMAPs show expression of relevant transcription factors and their predicted disrupted motifs, illustrating how cell-type-specific transcription-factor activity can reverse the regulatory effect of the same variant.

To determine how regulatory variants are shared across genes and immune cell types, we developed FLANDERS, a scalable fine-mapping and colocalisation framework that groups gene–cell-type associations likely to be driven by the same causal regulatory variant (eSNP) into *cis*-regulatory modules, which are treated as discrete units in downstream analyses. Details and benchmarking of the approach can be found in **Supplementary Note** and **Fig. 1**. Fine-mapping identified 114,063 statistically independent regulatory signals (i.e. independent associations between SNPs and the expression of a specific gene expression in a single cell type**; Table S1**), corresponding to 1.29 conditionally independent signals per gene–cell-type pair.

Using FLANDERS, we grouped 114,063 independent eQTL signals into 41,691 cis-regulatory modules, each represented by a single eSNP (Table S1). Most eSNPs (∼85%) regulated a single gene. When regulatory effects were shared, this occurred primarily across cell types rather than across genes: 38.6% of eSNPs regulated either a single gene (24.1%) or multiple genes (14.5%) across cell types, whereas only 1.2% regulated multiple genes within a single cell type. The remaining 60.2% were associated with a single gene-cell-type pair, of which 46% were confidently classified as cell-type-specific after accounting for differences in discovery power using multivariate adaptive shrinkage (mashr) ^27^. The proportion classified as cell-type-specific was unrelated to sample size or cell count (**Fig. S1**), indicating that mashr effectively accounted for these sources of variation in statistical power. Overall, approximately 85% of causal variants regulated a single target gene, indicating that differences in regulatory sharing arise primarily from the number of cell types in which they acted rather than the number of genes they influenced.

We next examined the organisation of regulatory pleiotropy across tissues, immune lineages and cell types, considering five distinct hierarchical levels of sharing, from multi-tissue to cell-type restricted (**Fig. 2b**). Comparison with 47 non-blood tissues from the GTEx project ^8^ revealed that nearly one-third of immune eQTLs were active in at least one non-blood tissue (**Fig. 2c**; 12,361 multi-tissue eQTLs, 29.7%), of which the majority regulated the same target gene (74.5%, **Table S2**). Multi-tissue activity was most frequent among broadly pleiotropic immune eQTLs, but was also detected for almost one-quarter of variants classified as cell-type-specific within PBMCs (**Table S2**), indicating that tissue- and cell-level specificity represent partly independent dimensions of regulatory activity. Among PBMC-restricted variants, regulatory effects showed strong lineage specificity: 58.8% of eQTLs were restricted to lymphoid cells and 28.9% to myeloid cells, while only 12.3% were shared across the two compartments (**Fig. 2d, Table S2**). This compartmentalisation extended to individual cell types. In contrast, lineage specificity was much less pronounced at the gene level. Most eGenes were shared across tissues or immune lineages, with only 24.2% (3,746 of 15,419) restricted to a single lineage (**Fig. S2**). Thus, lineage specificity appears to arise primarily from variants that are selectively active in defined cellular contexts, rather than from lineage-restricted expression of the genes themselves.

To test whether these patterns reflect distinct regulatory mechanisms, we compared the genomic architecture of eQTLs across classes of regulatory pleiotropy. Multi-tissue eQTLs were predominantly promoter-proximal, whereas increasingly restricted classes occurred progressively farther from their target genes and showed greater overlap with cell-type-specific accessible chromatin (**Fig. 2c**; median TSS distance 1.4 kb vs 23.4 kb; ANOVA p-value=2×10^-231^, **Fig. S3**). Increasing specificity was also associated with progressive depletion from promoter contacts, active transcription start sites, active enhancers, highly accessible chromatin and regions with high activity-by-contact scores, together with enrichment in low-signal and weak Polycomb-repressed chromatin marked by H3K27me3 (**Fig. 2e**, **Table S3**). This interpretation is consistent with previous evidence linking cell-type-specific regulation to distal regions with dynamic H3K27me3 states ^28^. However, when annotations were matched to the relevant lineage or broad cell class, each pleiotropy category remained enriched in accessible chromatin within its corresponding compartment (**Fig. 2e**, **Table S3**). Similarly, eGenes in each immune compartment were significantly enriched for pathways characteristic of that compartment’s known function (e.g., T and B cell categories enriched in adaptive_immune_response GO_BP; **Fig. S4**, **Table S4**). These findings suggest that hierarchical pleiotropy arises from a similarly organised regulatory landscape, ranging from constitutively active promoter-proximal elements to distal elements whose activity is progressively restricted across tissues, lineages and cell types.

Finally, to further characterise functional convergence for pleiotropic SNPs, we investigated whether eQTLs exerted concordant effects on gene expression (i.e. trait-increasing or trait-decreasing) across cell types, or whether their direction differed across cellular contexts. Regulatory effects were highly conserved across contexts, with approximately 95% of pleiotropic eQTLs having concordant effects (as measured from the signed beta of the eQTL) in every cell type in which they were active. The remaining 5% showed directional divergence (**Fig. 2f)**, predominantly between myeloid and lymphoid lineages rather than between closely related cell types (for example, monocyte-to-T-cell comparisons: 12.2% observed versus 8.4% expected; *r* = 0.18, p-value*=*4 × 10^-13^; **Fig. 2g; Table S1** and **Table S5**). Variants with discordant effects were enriched for disruption of transcription-factor binding motifs (OR=1.92; two-sided Fisher’s exact test, p-value*=*0.0078; **Fig. 2h Table S6**), including ETS-, SPI-, RUNX- and KR-like motifs which recur in highly confident finemapped variants (PIP>0.8)(**Fig. 2i**; **Fig. S5)**. At representative loci, lineage-specific expression of transcription factors including *PRDM1*, *EGR1* and *KLF4* reversed the transcriptional effect of the same variant across immune contexts (**Fig. 2h**). Thus, although regulatory effects are generally directionally stable, a small subset functions differently in lineage-specific transcription-factor environments.

These analyses establish regulatory pleiotropy as a hierarchy extending from multi-tissue programmes to terminal cell-type-specific effects. This hierarchy connects the genomic organisation of regulatory elements to the functional diversification of genes across innate and adaptive immune compartments, with lineage-specific transcription-factor logic reversing the direction of effect at only a small minority of loci.

### Blood regulatory variation shapes the human phenome and resolves immune components of complex disease loci

Having established a map of immune regulatory variation, we next asked whether its hierarchical organisation across tissues and cell types was reflected in the regulatory mechanisms underlying susceptibility to human disease. We used FLANDERS to integrate our single-cell *cis*-eQTL atlas with association signals from the GWAS Catalog and FinnGen, spanning 3,628 phenotypes (1,188 diseases and 2,440 quantitative traits) from 18,511 genome-wide association studies (**Table S7**), and systematically tested whether disease-associated variants and immune eQTLs shared a common causal SNP.

This analysis identified widespread sharing between regulatory and phenotype-associated variants (**Fig. 3a, Fig. S6),** with 162,338 colocalisations (PP.H4 ≥ 0.75) among 945,969 tests, successfully linking 2,089 unique diseases and quantitative traits to *cis*-eQTLs for 40,175 gene–cell-type pairs (comprising 9,077 unique genes; **Table S8**). Each phenotype locus typically implicated one or two candidate genes across four immune cell types. Some signals exhibited extensive phenotypic pleiotropy, including *TMEM258*, *EML2* and *PSRC1*, each colocalising with hundreds of diseases or quantitative traits (448, 256 and 214, respectively) (**Table S8**). Although primary *cis*-eQTL association signals were more likely to colocalise with disease than secondary signals, 15.4% of disease colocalisations were explained exclusively by secondary signals, highlighting the value of comprehensive fine-mapping for resolving complex disease loci (**Fig. S7**).

**Figure 3.**
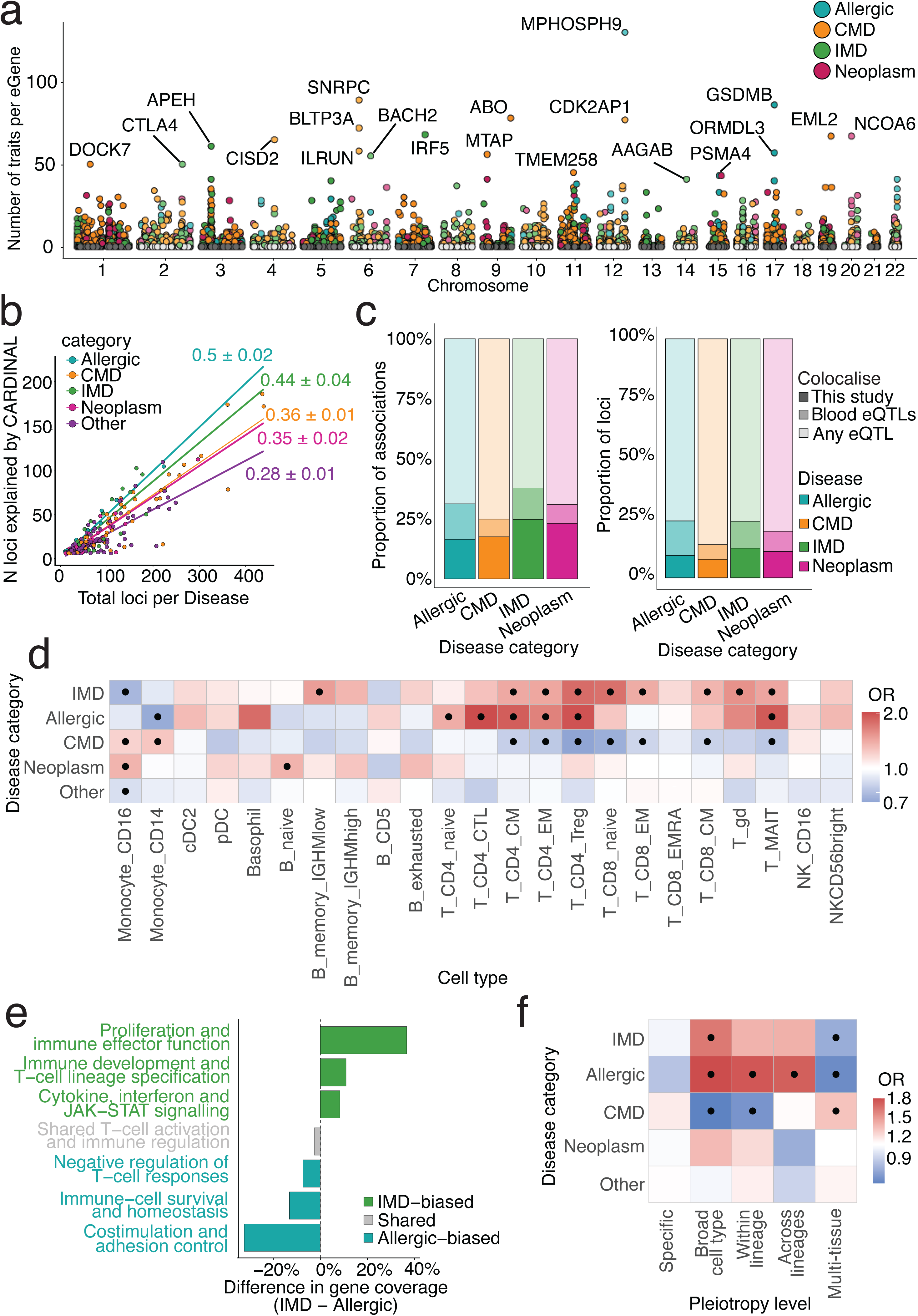
Hierarchical immune regulatory programmes explain genetic susceptibility across the human phenome. a, Disease pleiotropy among colocalising eGenes. Points represent eGenes, positioned by genomic location and number of colocalising traits; colours indicate the predominant disease class. Selected pleiotropic genes are labelled. **b,** Number of disease loci colocalising with CARDINAL PBMC eQTLs versus total GWAS loci. Colours indicate disease classes; slopes (± s.e.m.) represent the proportion explained by PBMC eSNPs. **c,** Proportions of GWAS associations (left) and independent loci (right) explained uniquely by CARDINAL PBMC eQTLs (dark), also by blood eQTLs (intermediate) or by broader eQTL resources (light). **d,** Cell-type enrichment of disease-colocalising eQTLs. Immune-mediated and allergic diseases predominantly involve adaptive immune cells, cardiometabolic diseases involve monocytes, and neoplasms show broader distributions. **e**, Biological processes differentially enriched between immune-mediated (IMD) and allergic diseases. Bars show differences in gene coverage. IMDs favour cytokine, interferon, developmental and proliferative programmes; allergic diseases favour co-stimulation, adhesion and immune homeostasis. **f,** Enrichment of disease-colocalising eQTLs across regulatory hierarchies. Immune-mediated and allergic diseases favour intermediate immune programmes, whereas cardiometabolic diseases favour organism-wide effects. In **d, f**, colours indicate odds ratios and black circles denote significant enrichments after multiple-testing correction.

A substantial proportion of association signals across immune-mediated, allergic, cardiometabolic and neoplastic diseases could be linked to genetically regulated gene expression in PBMCs. Specifically, 19–31% of disease-association signals, representing 28–50% of independent loci depending on disease class, shared a likely causal variant with a PBMC eQTL (**Fig. S8**; **Fig. 3b**). These findings implicate altered expression of specific genes in defined immune-cell populations as a potential mechanism through which these loci influence disease risk. Notably, 1,883 disease associations were linked exclusively to *cis*-eQTLs detected in our single-cell atlas and were not resolved by the bulk-tissue eQTL resources available through the Open Targets Platform ^29^ (34 studies across 50 tissues/cell types; **Table S9**). These associations accounted for up to one-quarter of all colocalised signals in immune-mediated diseases (**Fig. 3c**), revealing regulatory mechanisms that are masked when expression is averaged across heterogeneous tissues or cell populations. For example, at the chr11:128154056:128651691 asthma locus, *ETS1* had previously been prioritised on the basis of its position and experimental evidence, although no colocalisation had been detected in existing datasets. Here, we show that the asthma association colocalises with *ETS1* expression in several CD4⁺ T-cell subtypes, highlighting the importance of cell-type-resolved eQTL data for dissecting disease loci (**Table S10**).

We applied stringent filtering criteria (**Methods**) to prioritise the most likely effector gene and immune-cell context at each locus, and used Mendelian randomisation to infer whether genetically increased gene expression was associated with higher or lower disease risk. This approach reduced locus complexity by approximately one-third: the mean number of candidate genes per locus decreased from 1.84 to 1.34, and the mean number of candidate cell types from 3.96 to 2.67 (**Fig. S9**), thus refining disease-associated loci into more interpretable gene–cell-type relationships and establishing the predicted direction of their effects on disease risk. For example, the Crohn’s disease-associated locus chr16:28222090:29084436 initially showed colocalisation with 15 genes across 21 cell types. Our procedure refined these associations to prioritise *IL27* in CD14+ monocytes as the most likely causal gene–cell-type combination. *IL27* is predominantly produced by myeloid cells and can promote T-cell activation ^30^. A proinflammatory role has also been demonstrated in a mouse model of T-cell-mediated colitis, in which transfer of T cells lacking the IL27 receptor resulted in reduced colonic inflammation, supporting a direct contribution of IL27 signalling to intestinal inflammation in this model ^31^. Together, the myeloid origin of IL27 and its proinflammatory activity in this setting are consistent with our prioritisation of CD14+ monocytes and the inferred association between higher *IL27* expression and increased susceptibility to Crohn’s disease. This example illustrates how our procedure can refine broad colocalisation signals into biologically plausible gene–cell-type hypotheses.

Distinct classes of disease engaged different immune compartments. Across all disease groups, T cells accounted for the largest fraction of colocalisation events, followed by monocytes (**Table S11**). However, the relative contribution of immune lineages differed markedly between disease classes (**Table S12**). Immune-mediated and allergic diseases were significantly enriched for lymphoid regulatory effects (41% of colocalisation signals, OR=1.33 p-value=1.58x10^-3^, and 39%, OR=1.82 p-value=1.32x10^-5^, respectively), whereas cardiometabolic diseases preferentially colocalised with myeloid eQTLs (27.9%, OR=1.29, p-value=3.09x10^-7^; **Table S12)**, particularly within CD14⁺ and CD16⁺ monocytes (**Fig. 3d**, **Table S11**). At finer cellular resolution, allergic diseases predominantly involved CD4 memory T cells and regulatory T cells, together with other CD4⁺ subsets, whereas immune-mediated diseases additionally implicated CD8 T cells and memory B cells, indicating distinct adaptive immune mechanisms despite broad overlap in lymphoid involvement (**Table S11)**.

Consistent with these cellular enrichments, PBMC-restricted eGenes exhibited disease-specific functional organisation (**Fig. 3e**). Immune-mediated and allergic diseases shared a core programme of T-cell activation, lymphocyte differentiation, adhesion and inflammatory regulation, whereas immune-mediated diseases showed broader enrichment for cytokine signalling, interferon responses, lymphocyte proliferation and effector functions (**Table S13**, 242 enriched terms at FDR ≤5%). In contrast, cardiometabolic diseases and neoplasms showed no functional enrichment, suggesting that many of these associations reflect broader systemic regulatory mechanisms rather than immune-specific biology.

Finally, we sought to test whether eQTL-disease relationships followed a similarly structured hierarchical organisation across domains. Previous studies have shown that disease-associated variants preferentially colocalise with regulatory effects detected in disease-relevant tissues and cell types ^17^. Our hierarchical framework allowed us to ask a different question: at what biological scale do disease-associated regulatory effects operate? Rather than considering cellular specificity as a binary property, we assigned each disease-colocalising eQTL to successive levels of regulatory breadth, ranging from multi-tissue effects (shared across non-blood tissues) to *cis*-eQTLs active within the PBMC compartment, haematopoietic lineages, major immune cell classes or individual cell types (as in **Fig. 2d**). Compared with other disease groups, immune-mediated and allergic diseases were preferentially associated with regulatory effects acting at intermediate levels of the haematopoietic hierarchy, including PBMC-wide, lineage-restricted and major-cell-type eQTLs, and were significantly depleted for multi-tissue effects (OR=1.3-1.8 for intermediate classes; vs OR=0.63-0.72 for multi-tissue effects, **Fig. 3f**). By contrast, cardiometabolic disease loci were enriched for multi-tissue regulatory effects and depleted for lineage-restricted immune regulation (OR=1.2 and OR=0.6, **Fig. 3f**). Notably, strictly cell-type-specific eQTLs were not enriched in any disease class (**Fig. S10**). Across disease classes, these eQTLs were also less likely than multi-tissue eQTLs to colocalise with disease associations, even after accounting for differences in allele frequency, distance to the transcription start site and eQTL effect size and other possible confounders (OR=0.5 for cardiometabolic diseases to OR=0.7 for immune-mediated diseases; **Fig. S11**). These analyses indicate that disease predisposition is predominantly linked to regulatory effects shared across immune-cell populations, suggesting that inherited risk acts through core immune functions deployed across multiple cellular contexts rather than through mechanisms confined to individual cell types. Resolving these mechanisms therefore requires identifying not only the causal variant and effector gene, but also the hierarchical level —across tissues, immune lineages or individual cell types— at which regulatory activity operates. This framework extends genetic fine-mapping beyond variant-to-gene assignment to reveal how inherited variation perturbs the cellular organisation of core immune functions to influence human disease.

### Genetic architecture of cellular programmes links *cis*-regulatory variation to multicellular gene networks

In *cis*-eQTL frameworks, genetic variants are generally interpreted through their effects on nearby genes. However, the biological consequences of these local regulatory perturbations are likely to extend beyond individual transcripts to coordinated transcriptional programmes operating across multiple cell populations. How *cis*-regulatory effects propagate into higher-order gene networks, and how these contribute to complex disease beyond individual eQTLs at single cell resolution, is largely unexplored. To address this question, we integrated fine-mapped *cis*-eQTLs with multicellular gene programmes inferred from single-cell transcriptomes using Latent Interaction Variational Inference (LIVI) ^32^ (**Methods**). LIVI is an interpretable deep-learning framework that infers latent factors corresponding to transcriptional programmes that vary across individuals and cell states (LIVI programmes), facilitating *trans*-eQTL mapping at single-cell resolution. Specifically, donor-level LIVI programme values can be treated as quantitative traits and tested for association with genetic variants, enabling the discovery of *trans*-regulatory effects on co-regulated gene programmes rather than individual genes.

We trained LIVI on the expression profiles of 15,268 genes across 6.7 million cells from 4,428 UKB participants to infer 1,200 LIVI programmes (**Methods**), each recapitulating diverse aspects of immune biology (**Fig 4d**; **Fig. S12a**; **Table S14).** We then tested the 1,200 LIVI programmes for associations with 7,254 loci identified in the *cis*-eQTL analysis (see Methods), identifying 4,209 significant associations (*p* < 2x10^-5^), termed factor-QTLs (fQTLs), across 1,072 LIVI programmes **(Methods**). Fine-mapping identified 4,908 independent fQTL association signals, and subsequent colocalisation analysis (**Fig. 4a**) prioritised 6,219 shared *cis*–LIVI-programme regulatory events involving 2,415 *cis*-eSNPs and 1,056 LIVI programmes sharing the same causal variant (**Table S15**).

**Figure 4.**
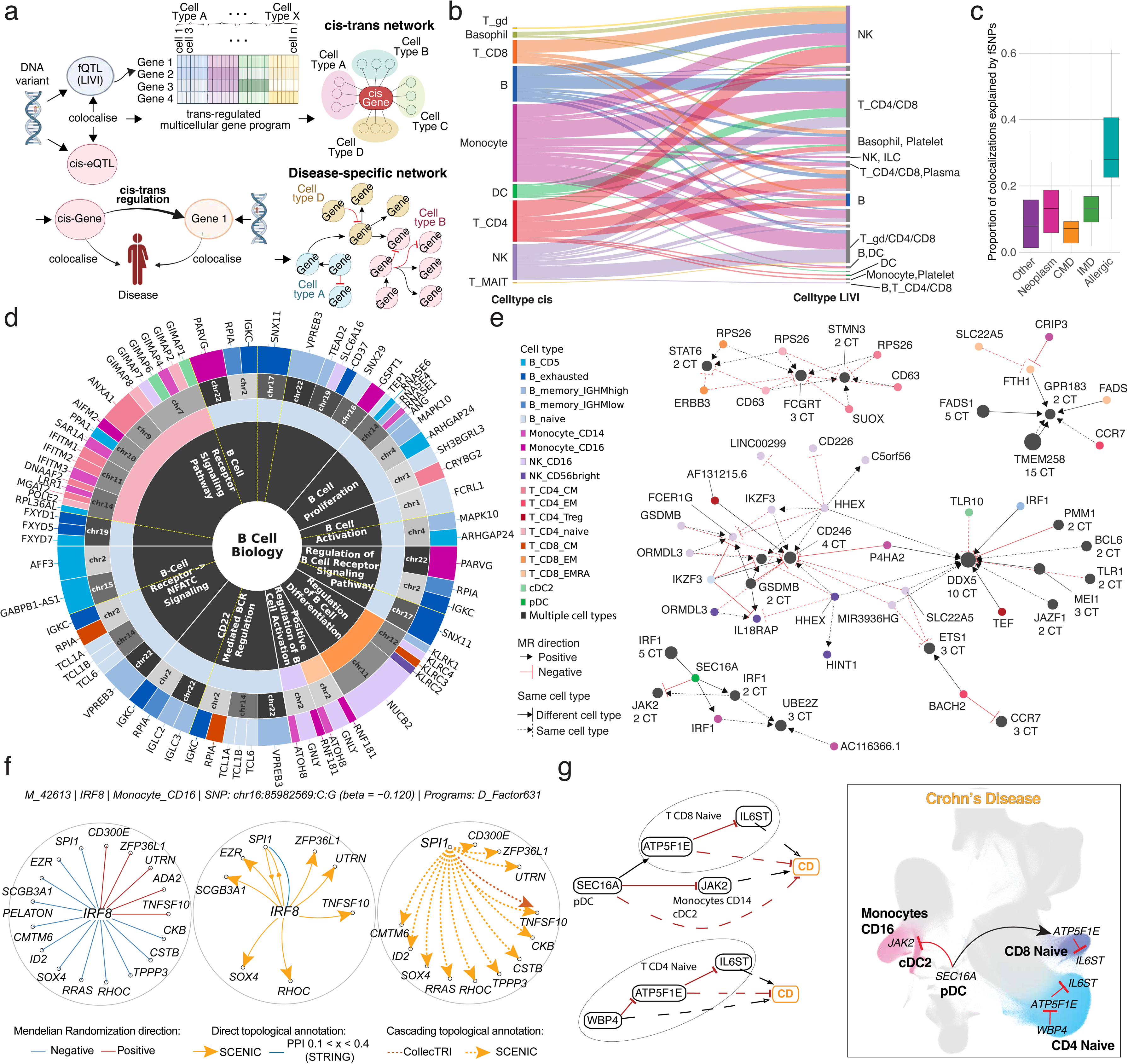
LIVI factor genetics enables the investigation of gene-gene regulatory relationships. a, Overview of the approach. LIVI factor-associated SNPs (fSNPs) are colocalised with cis-eSNPs to identify regulators of multicellular programmes and define cell-type-resolved *cis–trans* relationships (top). Disease-specific graphs comprise edges for which both the cis-eSNP and trans-eGene colocalise independently with the same disease; mediation analysis validates these edges (bottom). **b,** Sankey diagram of cross-cell-type relationships supported by colocalising cis-eQTL–fQTL pairs. Nodes indicate the cis-eQTL cell type (left) and cell types contributing to the LIVI programme (right). Link width represents the number of colocalisations and colour indicates the cis cell type; only transitions supported by at least two events are shown. **c,** Proportion of disease loci explained by fSNPs, stratified by disease category. fSNPs explain a substantial fraction of loci, reaching a median of 30% for allergic diseases. **d,** Sunburst plot of colocalised cis-eQTLs and fQTLs for B-cell-related LIVI programmes. Rings show, from inside out, programme category, biological annotation, LIVI cell type, eSNP chromosome and cis-eGene. Solid white lines separate annotations and dashed yellow lines separate programmes sharing an annotation. Differences between LIVI and cis-eGene cell-type colours indicate cross-cell-type regulation. **e,** Asthma-specific *cis–trans* regulatory network. Nodes represent genes within cell types. Edge colour and shape indicate the direction of effect relative to increased cis-eGene expression, while dashed and solid lines denote cell type autonomous and non cell type autonomous relationships, respectively. Identical relationships involving multiple cell types were collapsed. **f,** Internal topology of an *IRF8*-associated LIVI factor comprising 17 genes. *IRF8* interacts directly with a single programme member, which regulates the remaining genes through downstream interactions. **g,** Crohn’s disease regulatory relationships converging on *IL6ST*. *SEC16A* expression in pDCs is protective by reducing *JAK2* and *IL6ST* expression through *ATP5F1E*, whereas *WBP4* increases risk by inhibiting *ATP5F1E*, thereby increasing *IL6ST* expression (left). Single-cell estimates identify the cell types mediating each *cis–trans* relationship (right). Solid lines represent the inferred graph while dashed lines represent the observed MR effect estimated through *cis*-eQTL colocalisations.

The majority of *cis*–LIVI-programme relationships implicated closely related immune cell types (e.g., within B or T cells; 71%), while the remaining one-third propagated into programmes operating in different immune cell types (**Table S15**). These cross–cell-type relationships were not randomly distributed; *cis*-eQTLs identified in T and B cell populations frequently converged on NK-cell transcriptional programmes (and vice versa), whereas monocyte *cis*-eQTLs preferentially connected to programmes spanning multiple lymphoid populations (**Fig. 4b**). We hypothesised that *cis*-eQTL-programmes defining within-cell-type and cross-cell-type relationships may involve transcription factor cascades and cell-to-cell signalling.

To investigate this hypothesis, we first tested whether *cis*-eGenes colocalising with LIVI programmes were enriched for transcription factors (TFs), and whether the corresponding programmes contained known target genes of the respective TF. *Cis*-eGenes colocalising with LIVI programmes were indeed enriched for TFs (OR = 1.24, *p* = 0.021; **Table S16**; **Methods**), and nearly one-third of these TFs were connected to programmes encompassing at least one of their known targets, significantly more than expected by chance (OR=1.48, *p* < 10^-4^; **Table S16**; **Methods**). We observed a similar enrichment when considering signalling ligands involved in cross–cell-type interactions. Among *cis*-eGenes encoding such ligands, more than half colocalised with LIVI programmes enriched for the respective ligand’s downstream targets as predicted by NicheNet ^33^ (*p* < 0.05, **Table S17**; **Methods**). For instance, at *ICAM2*, *cis*-regulation in effector memory CD4^+^ T cells and CD14^+^ monocytes colocalised with a programme active in CD16^+^ NK and EMRA CD8^+^ T cells that contained its predicted targets, *DUSP1* and *FOS* (**Fig. S12e-g**). This is consistent with *ICAM2*’s reported role in NK cell cytotoxicity ^34^ and migration ^35^. Additionally, *ICAM2*’s receptor genes, *ITGB2* and *ITGAL*, are expressed in CD16^+^ NK cells in our dataset (**Fig. S12c-d**).

Next, we used the 6,219 colocalisation events to construct *cis–trans* regulatory networks, linking each *cis*-eSNP–eGene association in its cellular context to the genes and cell types comprising the colocalising LIVI programme. This effort yielded a network including 292,886 edges (83,918 unique for source module, source gene, LIVI programme and target gene combinations, **Table S18**, **Methods**). Edges were classified as cell-type-autonomous when the corresponding *cis*-eQTL and LIVI programmes mapped to the same cell types (63.3%), and as non-autonomous when their cellular contexts did not overlap (36.7%); these categories were mutually exclusive. We expanded on the prior annotation effort using different sources of evidence, including TF–target regulons using both CollecTRI and SCENIC+-inferred activator and repressor networks derived from our own dataset, ligand–receptor interactions from the Liana consensus resource, and RNA-binding protein (RBP)–target, peptidase–substrate (MEROPS), and protein–protein interaction (PPI, STRING) resources (testing directional annotations separately for the *cis*-eGene as regulator or as target; **Methods**). Cell-type-autonomous edges were significantly enriched for TF–target annotations (CollecTRI, SCENIC+ activator and repressor) both irrespective of direction and when the *cis*-eGene acted as regulator, and for RBP–target annotations specifically when the *cis*-eGene acted as regulator. Conversely, edges were enriched among non-autonomous pairs only when the *cis*-eGene was the target of a repressor TF. No significant enrichment was observed for ligand–receptor or peptidase–substrate annotations in either direction, or for PPI (tested irrespective of direction only, given its undirected nature) (**Fig. S13, Table S19**).

To validate our results with orthogonal information, we assessed replication of our *cis*-*trans* pairs in the largest *trans*-eQTL study to date ^36^. A total of 454 of 52,203 pairs were supported, significantly exceeding the 188 pairs expected by chance (odds ratio = 2.46, Permutation P <0.01). Among supported pairs, 77% showed concordant effect directions (binomial test, P = 3.94 × 10^-28^).

Additionally we systematically compared the identified cis-trans pairs with a Perturb-seq dataset for resting CD4^+^ T cells ^37^, focusing on pairs with a single effect direction (52,203 pairs). We found that 319 of 5,966 (5.34%) overlapping pairs were nominally significant (FDR<0.05) (**Table S18**), exceeding the permutation-based expectation (4.57%; permutation P = 0.03), with 62.69% showing concordant effect direction (binomial test, P = 6.73 × 10^-6^). Restricting the analysis to intra-cellular CD4^+^ T cell effects increased overlap to 8.46% (171/2,019, permutation P = 0.02), while directional concordance remained similar (62.57%; P = 1.25 × 10^-3^).

These analyses provide a genetically anchored framework for reconstructing putative regulatory paths linking genes within and across immune-cell types, resolving the potential propagation of *cis*-regulatory effects through multistep intracellular cascades - as illustrated for the *IRF8* monocyte programme (**Fig. 4f)**. Genetically regulated expression of *IRF8* was associated with a 17-gene LIVI programme that included the transcription factor *SPI1*. Only a minority of the 17 genes were direct targets of *IRF8*, and *SPI1* was the sole programme member supported by evidence of a direct protein–protein interaction with *IRF8*. SCENIC+ regulons, however, linked *SPI1* to 13 of the remaining 16 genes, suggesting that the effect of *IRF8* may propagate through *SPI1* to coordinate the broader programme. We identified analogous regulatory cascades among both cell-type-autonomous and non-autonomous edges (**Fig. S14-S15**), consistent with intermediate regulators relaying and amplifying local genetic perturbations into broader transcriptional responses. Collectively, these findings reveal a multilayered regulatory architecture through which *cis*-acting variation can influence coordinated gene programmes within and across immune-cell types.

### Multicellular gene programmes connect disease-associated genes into regulatory networks

Because fQTLs act on multicellular gene programmes rather than individual genes, we hypothesised that they could disproportionately contribute to complex disease. We therefore asked whether variants associated with these programmes were more likely than other *cis*-eQTLs to colocalise with disease-associated loci (**Methods**). Across all disease categories, fQTLs showed substantially greater enrichment for disease associations (**Fig. 4c**; OR=1.7, p-value=1×10^-19^, **Fig. S16, Table S20**), with particularly strong effects observed for allergic diseases (OR=4.5, p-value=2.3x10^-25^, **Fig. S16, Table S20)**. Although fQTLs represented only a subset of all regulatory variants, they accounted for approximately 10% of disease colocalisations overall and up to one-third of allergic disease loci (**Fig. 4c**), suggesting that regulatory variants acting through multicellular gene programmes have a disproportionate influence on human disease.

We next asked whether these *trans*-regulatory relationships could extend genetic interpretation beyond individual disease genes to reconstruct higher-order regulatory networks implicated in disease. Specifically, we connected cis-regulated genes to genes in their associated LIVI programmes when both genes independently colocalised with the same disease through their respective cis-eQTL signals (**Methods**). To avoid residual local effects, we excluded all programmes containing the *cis*-regulated gene, thereby restricting the analysis to likely *cis–trans* regulatory relationships.

This analysis revealed extensive regulatory connectivity across the human phenome (18,449 trait *cis–trans* relationships triplets; 10,740 *cis–trans* edges; 487 *cis* genes; 672 *trans* genes; 196 traits including 61 diseases, **Table S21**). Half of all *cis-trans* edges occurred within the same immune cell type (cell-type-autonomous), whereas the remainder linked distinct immune populations (non-autonomous). However, this balance differed markedly across disease classes. Cardiometabolic and allergic diseases were dominated by non-autonomous regulatory interactions (65.2% and 67.9% respectively), whereas immune-mediated diseases and neoplasms preferentially involved cell-type-autonomous regulatory networks (61.2% and 65.8% respectively) (**Fig. S17**), reflecting disease-specific engagement of immune programmes, with some predominantly mediated through intercellular communication and others through regulation within specialised immune compartments.

The resulting disease graphs revealed extensive convergence of independent genetic loci onto shared regulatory pathways. The complexity of these networks varied substantially across diseases, with asthma exhibiting the largest regulatory network (**Fig. 4e**), comprising 174 *cis*–*trans* regulatory interactions connecting 45 genes across 20 immune cell types. Highly connected hub genes emerged naturally from the network topology, identifying candidate regulatory bottlenecks through which multiple disease-associated variants converge. Unlike conventional colocalisation analyses, which typically assign variants to independent candidate genes, these multicellular networks reveal how apparently unrelated disease loci participate in common biological pathways.

To evaluate these inferred regulatory relationships, we performed mediation analyses testing whether the observed effect of each *cis-*gene on disease agreed in direction with the effect expected if it acted through its *trans*-regulated target. (**Methods**). Across all major disease classes, predicted and observed effects showed significant directional concordance (**Fig. S18,** 459 of 719 regulatory edges; 64% concordance, binomial test p=1.076x10^-13^), with particularly strong agreement for allergic diseases and several individual immune-mediated disorders, including Crohn’s disease, indicating that multicellular gene programmes accurately capture the propagation of regulatory effects through disease-associated networks.

The Crohn’s disease network illustrates the biological insight provided by this framework (**Fig. 4g, Fig. S19**). Multiple independent disease-associated loci converged on a common regulatory programme centred on *IL6ST* in naïve and memory CD4⁺ and CD8⁺ T cells. While conventional colocalisation could not distinguish among several candidate *cis*-regulated genes sharing the same causal variant, the downstream regulatory network highlighted *ATP5F1E* as the most likely intermediary linking upstream genetic associations to established inflammatory pathways. Independent regulatory inputs from *SEC16A* and *WBP4* converged on *ATP5F1E*, which in turn modulated *IL6ST* expression together with an interferon-γ-responsive transcriptional programme involving *JAK2*. Rather than identifying isolated disease genes, this analysis reconstructed convergent regulatory circuits linking multiple genetic loci to established inflammatory pathways implicated in Crohn’s disease.

Overall, multicellular gene programmes provide a mechanistic framework for connecting local genetic regulation to complex disease biology. Rather than acting through isolated *cis*-effects, many disease-associated variants propagate through coordinated transcriptional programmes that link genes, immune cell types and biological pathways into coherent regulatory networks. Our approach extends genetic fine-mapping from the identification of causal variants and effector genes to the reconstruction of the multicellular regulatory circuits through which inherited variation influences human disease.

### Rare variants independently validate regulatory mechanisms and therapeutic targets

Common-variant eQTLs capture regulatory effects that are sufficiently frequent to be observed at population scale, but are expected to miss highly constrained genes whose expression is primarily perturbed by rare deleterious variants. As whole-genome sequencing becomes increasingly available for large single-cell cohorts, rare variant association studies will provide an orthogonal opportunity to characterise these genes while independently validating regulatory mechanisms inferred from common variants. To explore this question, we therefore systematically tested the effects of rare (MAF < 0.01 in the CARDINAL subset) coding and regulatory variants on gene expression using burden tests across all major immune cell populations ^38^ (10 variant annotations related to expression; **Methods**).

Across 22 immune cell types, rare variant burden analyses identified widespread associations between predicted deleterious variants and gene expression (**Fig. 5a,b**; 6,777 rare-variant eQTL associations at 5% FDR, involving 2,875 genes out of the 13,225 tested, **Table S22**). Similar to common-variant eQTLs, discovery power depended on donor number and baseline gene expression, although associations involving highly deleterious classes of variants—including predicted loss-of-function, splice-disrupting and promoter-disrupting mutations—were disproportionately affected by limited carrier numbers (**Fig. S20, S21**). Variants predicted to produce severe functional consequences generally exhibited larger effects on gene expression than common *cis*-eQTLs and frequently acted across multiple immune cell types (**Fig 5f, S21d**).

**Figure 5.**
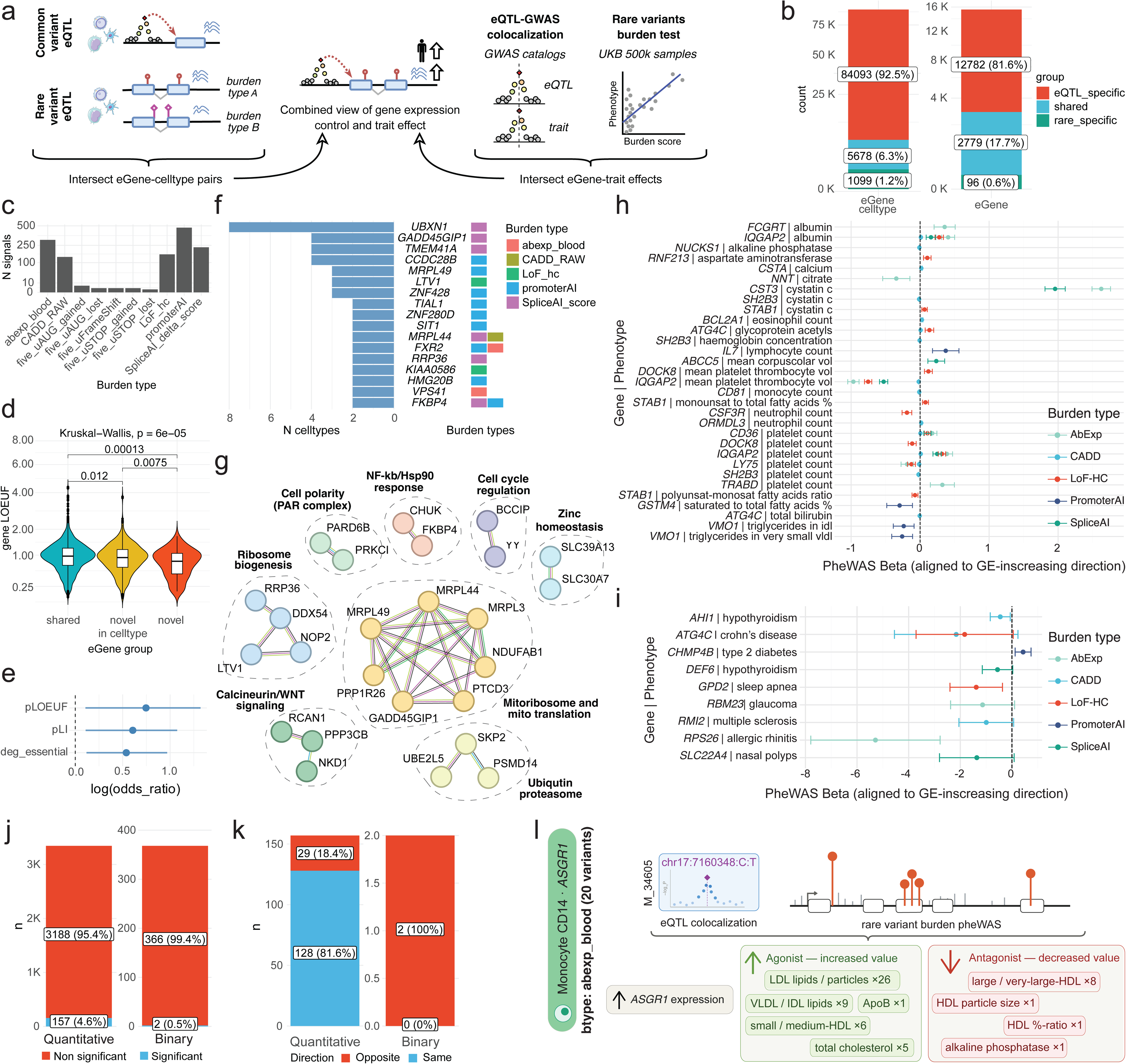
Allelic convergence supports regulatory mechanisms inferred from common variation. a, Framework integrating rare-variant burden tests, cis-eQTLs and phenome-wide associations to assess concordance across the allele-frequency spectrum. **b,** eGene–cell-type associations (left) and eGenes (right) detected by common variants, rare variants or both. Rare variants recover many common-variant eGenes while identifying additional constrained genes. **c,** Significant associations across functional burden masks, including AbExp, CADD, promoterAI, SpliceAI and predicted loss-of-function variants. **d,** LOEUF distributions across discovery classes. Rare-variant-specific eGenes show greater constraint. **e,** Enrichment of low-LOEUF, high-pLI and essential genes among rare-variant-specific associations. **f,** Representative genes across cell types (left) and contributing burden masks (right). **g,** Pathways enriched among rare-variant-specific genes, including ribosome biogenesis, mitochondrial translation, proteolysis, zinc homeostasis and cell polarity. **h, i,** Phenome-wide rare-variant effects for quantitative traits (**h**) and diseases (**i**) in 463,072 UK Biobank participants, aligned to the expression-increasing direction predicted by common variants. **j,** Proportions of common-variant predictions replicated by rare-variant burden tests. **k,** Directional concordance between common- and rare-variant associations. **l,** Allelic convergence at *ASGR1*: common-variant eQTL colocalisation and rare-variant burden associations independently support a shared lipid-regulatory mechanism and therapeutic direction.

Although most rare-variant associations were found in genes also identified by common-variant analyses, burden testing uncovered 1,099 additional regulatory signals across 96 genes detected exclusively through rare variants (**Fig. 5b,c**, **Table S23**). These 96 genes represent highly constrained eGenes (**Fig. 5d,e**), often derived from severe variant annotations (LoF_hc, PromoterAI and SpliceAI_score) with a recurrent effect across multiple cell types (**Fig. 5f**). These genes were implicated in essential cellular processes and core immune pathways (**Fig. 5g, Fig. S22**), suggesting that both deleterious coding variants and common regulatory variants in the genes are subject to strong purifying selection.

Finally, we asked whether rare variants support the disease or quantitative trait mechanisms inferred from common-variant genetics. For genes identified by both approaches, we compared phenotypic associations from rare variant burden analyses across the whole UK Biobank cohort with phenotypic associations predicted through common-variant eQTL colocalisation (**Methods**). Across all tested traits, we identified 159 instances in which common-variant eQTL colocalisation and a significant rare-variant burden association implicated the same gene and trait (**Fig. 5h–j**, **Table S24**). Of these, 80% (128/159) exhibited concordant genetically predicted directions of effect (**Fig. 5k, Table S24**). This high level of agreement indicates that common- and rare-variant associations frequently converge on shared biological mechanisms. Representative examples illustrate this allelic convergence. Both common and rare regulatory variation supported roles for *ASGR1* in lipid metabolism (**Fig. 5l**), *ORMDL3* in neutrophil count, *SH2B3* in renal function, *IL7* in lymphocyte homeostasis and *IQGAP2* in serum albumin, with remarkably consistent effects across variant classes (**Fig. S23**). These findings demonstrate that rare variants not only extend regulatory discovery into highly constrained genes, but also provide independent genetic validation for disease mechanisms and therapeutic targets inferred from common-variant single-cell genetics.

### Single-cell genetics guide therapeutic target prioritisation and validation

One of the principal current applications of human genetics is to identify therapeutic targets together with the direction in which they should be pharmacologically modulated. We therefore asked whether cell-type-resolved eQTLs could predict not only candidate target genes but also whether increasing or decreasing their expression would be expected to reduce disease risk. We classified colocalising gene–disease associations according to the genetically inferred direction of effect, defining candidate *predisposing* targets when increased expression was associated with an increase in disease risk and *protective* targets when increased expression was associated with a decrease in disease risk (**Methods**). Across all cell-type-resolved colocalisations, we identified 13,334 genetically supported gene–disease GWAS study relationships involving 3,071 genes and 432 disease endpoints (1,393 GWAS studies) (**Fig. 6a**; 6,732 predisposing, 6,369 protective and 233 both predisposing and protective associations, **Table S25**). Among genes implicated in multiple disease endpoints, more than half (53%, 836 out of 1591) exhibited consistent directionality across indications, identifying robust candidate agonist or antagonist targets, whereas the remaining (47%, 755 out of 1591) displayed disease-specific effects, indicating that the optimal therapeutic modality depends on disease context (**Table S25**). For example *IRF4* was consistently associated with dermatological conditions and inflammatory disease (**Fig. 6e**).

**Figure 6.**
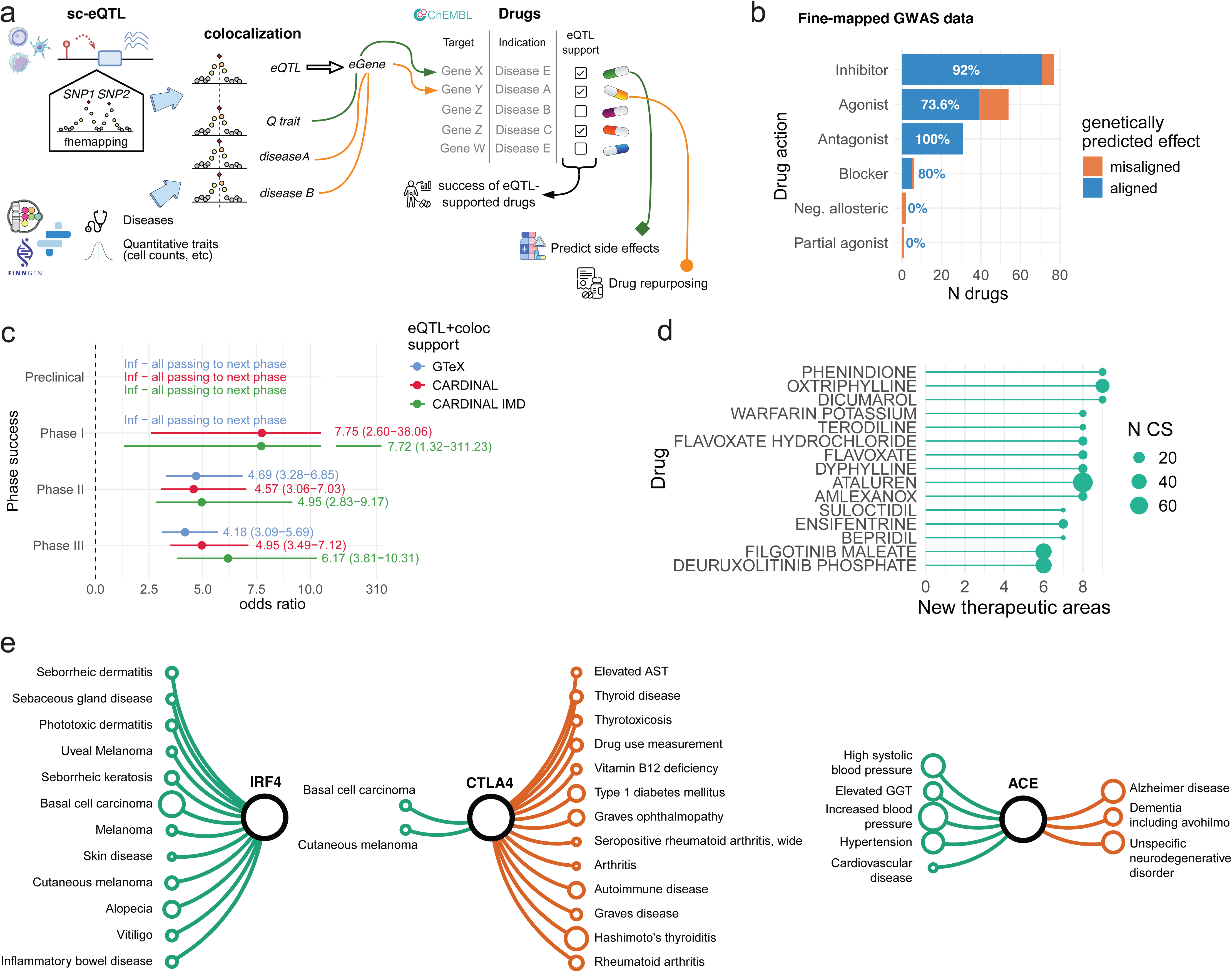
Human genetics guides therapeutic intervention through cell-type-resolved regulatory maps. a, Analytical framework integrating fine-mapped single-cell eQTLs and GWAS associations to prioritise disease genes, infer therapeutic direction and identify repurposing opportunities and potential adverse effects. **b,** Concordance between genetically inferred therapeutic direction and approved drug mechanisms. Bars show the proportion of concordant drug–target pairs; numbers indicate percentages. **c,** Enrichment of genetically supported targets across clinical-development phases. Odds ratios compare targets supported by disease-matched CARDINAL PBMC (red), immune-mediated disease-specific CARDINAL (green) or GTEx (blue) eQTL colocalisations. Disease-matched PBMC genetics shows the strongest enrichment. **d,** Approved drugs with the greatest number of additional genetically implicated therapeutic areas beyond their recorded current assignments. Point size indicates the number of supporting colocalisation signals (CS). Associations are shown before stratification by therapeutic direction. **e,** Examples of genetically supported repurposing and adverse effects. Networks connect targets to indications with therapeutic directions concordant (green) or discordant (orange) with the expected pharmacological effect.

To benchmark these predictions, we compared genetically inferred directions of effect with the mechanisms of action of approved drugs curated by the Open Targets Platform ^29^. Overall, genetically predicted therapeutic directions closely matched established pharmacology (**Fig. 6b**; 140 of 164 evaluable drug–target–disease combinations; 85.3% concordance, **Table S26**), with particularly strong agreement (92%) for inhibitory drugs. Furthermore, drug targets supported by disease-matched single-cell colocalisation were substantially enriched among successful clinical programmes compared with targets supported by eQTL evidence alone (Phase III OR=4.95 versus baseline OR=2.15, **Fig. 6c, Table S27**). Although multi-tissue GTEx identified regulatory evidence for a larger number of drug targets (1,882 vs 1421 at preclinical phase), disease-matched PBMC single-cell colocalisation achieved comparable predictive performance while providing substantially greater improvement over baseline regulatory evidence (**Fig. 6c, Table S27**). These findings indicate that immune-cell-resolved genetics provides highly discriminative evidence for therapeutic target validation and complements existing bulk-tissue resources.

We next asked whether these genetically supported regulatory effects could identify opportunities for drug repurposing across the human disease phenome. Integrating disease-colocalising genes with approved drug–target relationships from Open Targets identified hundreds of candidate drug–target–indication combinations supported by single-cell genetics (604 candidate repurposing links involving 224 approved drugs, 65 target genes and 69 disease indications, **Table S28**). Before accounting for therapeutic direction, these associations linked individual drug entries to as many as nine therapeutic areas beyond their recorded current assignments (**Fig. 6d**). Incorporating genetically inferred direction of effect allowed us to distinguish therapeutically concordant from potentially deleterious interventions. Of 604 drug–target–disease combinations, more than half (58%, 351) showed directional agreement between predicted genetic effects and the annotated drug mechanism, whereas approximately one-third (35%, 211) exhibited discordant effects suggestive of indication-specific contraindications or context-dependent biology; finally, a small percentage (7%, 42) had unresolved directionality (**Table S28**). To assess prior treatment precedent, we refined the annotations through an LLM-assisted review of existing evidence (**Methods**). Among the therapeutically concordant combinations, 131 had prior target- or pharmacological-class treatment precedent, demonstrating that genetic prioritisation recovered previously investigated therapeutic relationships. A further 27 had prior target-level biological evidence, providing mechanistic context for the proposed interventions. For 96 combinations involving 61 drug entries and 21 disease phenotypes, no relevant precedent was identified within the review scope, highlighting potential repurposing opportunities (**Table S28**).

Immune-mediated inflammatory diseases presented multiple opportunities for therapeutic repurposing. We identified 71 therapeutically concordant drug–target–disease combinations involving 42 drug entries. Excluding existing drug approvals and broad phenotypes overlapping approved indications left 61 candidate combinations involving 41 drug entries for additional indications within the IMD cluster (**Table S28**). These included predicted *JAK2* inhibition in ulcerative colitis and Crohn’s disease, *PDE4A* inhibition in rheumatoid arthritis, and *NR3C1* activation in Hashimoto’s thyroiditis. Evidence refinement identified prior target- or pharmacological-class treatment precedent for 44 combinations, supporting the biological relevance of the genetic prioritisation. Eight combinations had no relevant precedent identified within the review scope, including five rheumatoid arthritis combinations without recorded review concerns.

The cellular architecture of these repurposing opportunities was equally informative. CD14⁺ and CD16⁺ monocytes, together with CD16⁺ NK cells, accounted for the largest populations among therapeutically concordant drug–target–disease combinations. Supporting colocalisation evidence spanned multiple immune populations for 137 of 351 combinations (39%), corresponding to 23 of 65 distinct target–disease pairs (35%, **Table S28)**. Rather than identifying isolated disease-driving cell types, these results suggest that successful immune therapeutics frequently act on regulatory programmes shared across coordinated cellular compartments, consistent with the hierarchical organisation of immune regulation described above.

Finally, genetically inferred directionality also highlighted situations in which therapeutic perturbation may produce unintended consequences. Several targets exhibited disease-dependent or cell-type-dependent effects, implying that identical pharmacological interventions could be beneficial in one indication while detrimental in another. For example, stimulation of *CTLA4*, although therapeutically useful in selected cancer, was genetically predicted to increase the risk of several autoimmune disorders such as Type I Diabetes and Graves disease (**Fig. 6e**). This is in line with known side effects of this class of drugs, highlighting the power of these approaches. Similarly, inhibition of *ACE* was correctly associated with a reduction in blood pressure, however the genetic analysis suggests that it may lead to increased risk of Alzheimer’s Disease. Although these results clearly need validation, they suggest that integrating cell-type-specific regulatory genetics with therapeutic mechanism provides a systematic framework for anticipating both repurposing opportunities and potential adverse effects before clinical testing. Overall however, the effects on the main indication were generally stronger compared to the side effects (**Fig. S24**) highlighting that the drug was brought to market with a clear benefit.

Together, these analyses show that single-cell genetics extends human genetics from therapeutic target identification to therapeutic target prioritisation. By combining disease colocalisation with genetically inferred direction of effect and cellular context, single-cell regulatory maps can identify candidate therapeutic targets, predict the direction of pharmacological modulation, prioritise opportunities for drug repurposing and anticipate context-dependent adverse effects, thereby providing a comprehensive genetic framework for therapeutic development.

## Discussion

Inherited genetic variation shapes organismal phenotypes through coordinated activity across cells and tissues, yet regulatory effects are commonly interpreted as interactions between individual variants and nearby genes. By integrating population-scale single-cell transcriptomics with robust inference of sharing of genetic effects through the FLANDERS framework, we show that immune regulatory variation is instead organised across a hierarchy of biological scales and propagates through multicellular transcriptional networks. This organisation provides a scaffold connecting local genetic perturbations to disease mechanisms and therapeutic opportunities.

Our findings argue against a simple distinction between shared and cell-type-specific regulation. Regulatory effects span successive levels of organisation, from organism-wide activity to haematopoietic lineage-, cell-type- and cell-state-restricted effects, with distinct underlying genomic architectures. Broadly shared variants preferentially localise to promoter regions and constitutively active regulatory elements, whereas increasingly restricted effects arise from more distal and context-dependent chromatin. Importantly, restricted variants remain enriched in active regulatory elements within their cognate cellular compartments, indicating that specificity reflects the biological context in which regulatory elements are deployed rather than weaker regulatory activity.

This hierarchy changes how cellular specificity should be interpreted in disease genetics. Rather than preferentially acting through the most narrowly defined cell states, disease-associated variants were concentrated at intermediate levels of regulatory organisation. Immune-mediated and allergic diseases preferentially involved PBMC-, lineage- and broad cell-type programmes, whereas cardiometabolic disease showed greater contribution from organism-wide effects, consistent with more systemic regulatory mechanisms. Strictly cell-type-specific effects were depleted among disease colocalisations despite representing a large fraction of regulatory variation. This does not diminish their importance at individual loci; instead, it suggests that disease risk is often mediated by effects that are sufficiently restricted to perturb a relevant biological compartment, yet sufficiently broad to influence a substantial cellular programme. This distinction may reconcile our findings with previous reports of disease enrichment among cell-type-specific eQTLs, because specificity depends critically on the cellular and tissue hierarchy against which it is defined. Another point to consider is that our definition of tissue sharing is based on comparisons with bulk eQTLs, which have been shown to contain high levels of contaminating cells from other tissues ^39^. This is likely to overestimate the level of sharing for multi-tissue eQTLs, which future single cell studies in human reference tissues will address.

The consequences of regulatory variation also extend substantially beyond proximal targets. Multicellular gene programmes inferred by LIVI revealed how local perturbations propagate into coordinated transcriptional responses within and between immune populations. Direct transcription-factor cascades occurred preferentially within cell types, whereas cross-cell-type effects were more consistent with ligand-mediated signalling and indirect molecular interactions, providing complementary routes through which genetic effects can spread through the immune system. Thus, phenotypic pleiotropy need not require a variant to act independently in many cellular contexts. A comparatively local perturbation can acquire broad consequences through the topology of the regulatory network in which it is embedded.

This propagation provides an additional dimension for interpreting complex disease loci. Variants influencing multicellular programmes were disproportionately associated with disease, and *cis*–*trans* relationships connected independently associated loci into coherent disease-specific networks. These networks revealed convergence points receiving input from multiple loci and upstream regulators influencing multiple downstream genes, identifying candidate regulatory bottlenecks that are invisible when loci are considered independently. The ability to test whether genetically predicted and observed effects were directionally concordant across disease networks using the disease as anchor, allowed us to validate the biological structure captured by these programmes. These observations provide a population-genetic route from distributed disease associations towards the core regulatory processes through which their effects may converge.

This systems-level view also has direct implications for therapeutic genetics. Single-cell colocalisation refined both effector genes and their relevant immune contexts, while genetically inferred directions of effect recapitulated the mechanisms of established therapies and identified potential opportunities for repurposing. Disease-matched single-cell genetic support was strongly enriched among targets reaching late-stage clinical development, despite bulk tissue resources identifying a larger absolute number of targets, illustrating the complementary value of cellular resolution for target validation. Extending this framework across the phenome identified candidate repurposing opportunities as well as genetically discordant drug–disease relationships that may indicate adverse or context-dependent effects. Human genetics may therefore inform not only which gene to perturb, but in which cellular context, in which direction and with what potential consequences elsewhere in the phenome.

Rare variants provided an orthogonal test of these regulatory inferences. Burden analyses extended discovery to constrained genes that are poorly interrogated by common regulatory variation, while genes implicated independently by common and rare variants frequently showed overlapping phenotypic effects and predicted directions of action. This convergence across the allele-frequency spectrum supports the underlying causal mechanisms and illustrates the value of constructing allelic series that integrate common regulatory variation with rare, often more disruptive genetic perturbations. Such convergence should provide particularly strong evidence for therapeutic target prioritisation.

Relevant to the interpretation of this study, our analyses were restricted to circulating, predominantly resting immune cells and therefore are likely to capture only one component of the regulatory architecture underlying complex disease. Regulatory effects emerging in non-haematopoietic tissues, transient activation states, development or disease progression may be captured from population data (e.g., in contextual eQTLs ^26^), but their validation will require complementary sampling and targeted explorations. The restriction to individuals of European ancestry excludes inference of population-specific and highly cohort-dependent effects. Finally, multicellular programmes, ligand-mediated interactions and network edges remain genetically informed predictions rather than direct demonstrations of molecular causality. Resolving their internal topology will require larger and more diverse cohorts together with perturbational, spatial and temporal measurements that distinguish initiating regulators, intermediate nodes and downstream responses.

Overall, however, our findings underscore the importance of progressing from a variant-to-gene model towards a more explicit assessment of regulatory hierarchy, multicellular networks and disease contribution. In this view, our results suggest that inherited variation is deployed at different biological scales and its effects propagate through interconnected regulatory programmes rather than remaining confined to individual genes or cell types. Extending these maps across tissues, developmental stages and environmental contexts, and coupling them to perturbation experiments in controlled settings, should enable increasingly predictive models of how genetic perturbations reshape multicellular systems. Such models provide the necessary next step to derive a general framework for translating human genetic variation into disease mechanisms and therapeutic hypotheses.

## Methods

### Ethics

Recruitment descriptions: UK Biobank obtained approval from the North West Multi-centre Research Ethics Committee (MREC) UK NHS REC approval reference number: 21/NW/0157. Genes & Health obtained ethical approval from London South East NRES Committee of the Health Research Authority approval reference number: 14/LO/1240.

### CARDINAL single cell cis-eQTL dataset

We used *cis*-eQTL summary statistics generated from the CARDINAL PBMC single-cell RNA-sequencing dataset, as described in Giacopuzzi *et al.* ^26^. Briefly, the analysis included 4,452 UKB participants of European ancestry and 23 immune cell types meeting predefined sample and cell-count thresholds.. Cell-type-specific expression was normalised and aggregated by individual. *Cis*-eQTL mapping, performed using TensorQTL software ^40^. Variants within ±1 Mb of each gene’s transcription start site were tested, and significant gene–cell-type associations were identified at a Storey’s q-value on Aggregated Cauchy Association Test (ACAT) ^41^ gene-level p-value of ≤ 0.05. Full details of cohort selection, data processing, covariate adjustment and association testing are provided in Giacopuzzi *et al.* ^26^.

### TileDB QTL processing

To enable efficient storage and querying of QTL summary statistics, we used TileDB-sumstat (https://github.com/HTGenomeAnalysisUnit/TileDB-sumstat), a dedicated pipeline. This pipeline leverages TileDB technology (https://github.com/single-cell-data/TileDB-SOMA) to store the data as an indexed, multidimensional TileDB array, with chromosome, cell type, gene and genomic position defining the array coordinates, association statistics, including effect estimates, standard errors and allele frequencies, stored as attributes. Values that were constant across all variants for a given cell type–gene pair, such as sample size and phenotypic variance, were stored as array metadata, substantially reducing storage requirements. During ingestion into TileDB, variants were harmonised by alphabetically ordering the two alleles, designating the first allele as the effect allele. Where allele order was reversed, the sign of the effect estimate was inverted and the effect-allele frequency was updated accordingly. Following ingestion, a set of functions implemented in the Python Polars library were used to efficiently extract the regions defined by LocusBreaker for downstream analysis with the FLANDERS pipeline.

### The FLANDERS framework

The FLANDERS toolset (https://github.com/Biostatistics-Unit-HT/Flanders) is composed of a set of computational implementations aimed at simplifying analyses downstream of genetic association analysis, with a particular focus on fine-mapping and colocalisation as a means to study pleiotropy. It first fine-maps loci associated with each phenotype and then combines pairwise colocalisation with a graph representation of the resulting relationships to identify groups of credible sets that share an underlying causal SNP across phenotypes. The whole process is highly optimised in terms of computational and storage efficiency, and supports the incremental integration of previously fine-mapped datasets. FLANDERS primary output is “*cis*-regulated modules”, each comprising the set of phenotypes influenced by a shared causal variant at a given locus. Joint fine-mapping is then performed within each module, to improve fine-mapping resolution and generate refined association signals that can be directly tested for colocalisation with other traits and diseases. Details of the whole workflow and benchmarking of the new methods implemented can be found in the Supplementary Notes.

#### Locus identification with Locusbreaker

To identify the boundaries of genomic regions significantly associated with each trait, we implemented LocusBreaker, an algorithm that defines loci based on the shape of the association signal, as previously described by May-Wilson *et al*. ^42^. LocusBreaker requires two p-value thresholds: a significance threshold and a base threshold used to define the boundaries of the association peak. We applied LocusBreakers to all cell type–eGene pairs with an ACAT q-value ≤ 0.05. For pairs having no SNPs showing a nominal p-value < 1x10^-6^, the base threshold was set to one order of magnitude higher than the minimum p-value within the window. Otherwise, a fixed threshold of 1x10^-5^ was applied.

#### Fine-mapping and colocalisation

Following locus identification, fine-mapping and colocalisation analyses were performed within the FLANDERS framework. Each locus was fine-mapped using the Sum of Single Effects (SuSiE) ^43^ model with in-sample LD, allowing for up to 10 independent causal signals. When SuSiE failed to identify reliable credible sets or when the iterative Bayesian stepwise selection (IBSS) algorithm did not converge, Wakefield’s approximate Bayes factor (ABF) method ^44^ was used as an alternative, assuming a single causal variant within the locus. SuSiE-derived credible sets were subsequently subjected to quality control on statistical support and linkage disequilibrium–based purity metrics, and credible sets failing any of these criteria were excluded from downstream analyses. For each retained signal, FLANDERS then constructed a “coloc-sufficient” credible set comprising the conventional 99% credible set together with additional variants supported by high log approximate Bayes factor (lABF) values. This approach preserves the principal fine-mapping signal while retaining variants that may provide informative evidence during colocalisation. For each coloc-sufficient credible set, lABFs, conditional effect-size estimates, and corresponding standard errors were stored in a cs-AnnData object. Full implementation details and parameter settings are provided in the Supplementary Notes. Fine-mapping identified 114,063 credible sets for 88,513 eGene–cell-type pairs.

Pairwise colocalisation was subsequently performed within FLANDERS using iColoc, an extension of the coloc framework ^9^ which imputes the lABF of variants outside coloc-sufficient credible sets using the minimum lABF value observed in the corresponding region. Colocalisation was assessed between signals from different cell types and/or genes within the same population, restricting analyses to pairs of credible sets sharing at least one SNP. Evidence for a shared causal variant was defined as a posterior probability of PP.H4 ≥ 0.75 corresponding to PP.H4/(1-PP.H4) ≥ 3.

#### Cis-regulated modules identification and joint fine-mapping

We applied the second part of the FLANDERS workflow to identify which of the colocalised signals could be driven by the same underlying eSNP, thereby characterising their pleiotropic effects across cell types and genes. Briefly, FLANDERS combines pairwise colocalisation tests graph-based representation and SuSiE-HyPrColoc ^45^ (see Supplementary Notes) to group credible sets into modules representing signals consistent with a shared causal variant. For each module, joint fine-mapping was then used to identify a credible set estimating the posterior probability that each SNP is the causal variant underlying all participating traits. Again, we considered PPH4 ≥ 0.75 as significant evidence of colocalisation. For each module, we defined the SNP with the highest PIP as the “most-likely SNP” and used it to compare effect sizes and direction across traits, and as representative for the actual casual eSNP. eSNPs were classified in 4 different categories based on increasing pleiotropy: “Specific affecting a single gene in a single cell type; “Multi-gene affecting multiple genes within a single cell type; “Muti-context” affecting the same gene across multiple cell types; and “Highly pleiotropic affecting multiple genes across multiple cell types.

### Multivariate association testing with mashr

To assess how statistical power influenced the specificity of the observed associations, we used mashr ^27^. Mashr jointly analyses effect estimates across multiple conditions to identify recurring patterns of effect sharing. This approach improves effect-size estimates and statistical power, while distinguishing shared from condition-specific effects and accounting for estimation uncertainty. We treated cell types as conditions and gene-expression levels as traits. Mashr requires the estimation of two covariance matrices from the summary statistics: one using the strongest association effects for each trait across all conditions, and another using a set of randomly selected SNPs. Because this procedure requires the same SNPs to have been tested across all conditions, we excluded rarer cell types (B CD5, exhausted B, pDC, Basophils and CD16-bright NK). We consequently identified a set of 12,109 genes which were tested across all 18 common cell types and selected three random SNPs from the *cis* window of each gene. Following mashr analysis, we performed fine-mapping. We colocalised each credible set classified as cell-type ‘specific’ with the mashr credible sets from the same gene across all other cell types. This allowed us to identify ‘apparently specific’ credible sets, which appeared specific because the corresponding associations in other cell types had insufficient statistical power.

### Overrepresentation analysis of eGenes exclusive of each pleiotropy category

Cumulative partition-exclusive eGene identification. A cumulative definition of exclusivity pooled subsets of broad.cat levels into 9 hierarchical lineage/cell-class groups, plus a pan-immune PBMC_cumulative group spanning all 32 levels, retained as a reference despite being degenerate by construction. Redundant single-category partitions subsumed by a cumulative group were dropped and fine-grained subtypes with no higher-level grouping retained unchanged, yielding 33 partitions in total. For each partition, exclusive eGenes were those detected among its member broad.cat levels and absent from every broad.cat level not already included in that partition (its “rest” set).

Over-representation analysis (ORA) of cumulative-partition-exclusive eGenes. ORA was performed with clusterProfiler::enricher() on each partition’s exclusive eGene set, against two term resources: GOBP immune-pathway terms ^26^ and four in-house gene collections (immune-database genes, curated immune genes, splicing factors, transcription factors). The background/universe was fixed to all unique eGenes in the UKB eQTL dataset, with minGSSize = 10, maxGSSize = 500. Within each partition, p-values from both resources were pooled and corrected for multiple testing using the Benjamini–Hochberg procedure, separately per partition (padj ≤ 0.05 considered significant). PBMC_cumulative, being degenerate (query set equals the background universe), returned fold-enrichment = 1 in the ORA and no significant terms, serving as an internal negative control and was removed from the table of results. Partition membership, rest definitions and ORA results for all 32 partitions are given in **Table S4**.

### Functional annotation characterisation and enrichments

We selected four public datasets spanning distinct layers of regulatory annotation, each providing partial cell type matching with CARDINAL PBMC populations: Promoter Capture Hi-C (PCHi-C) (PCHiC_Feature_matrix_cutoff5.tsv) ^46^, chromatin state predictions from BLUEPRINT (BP ChromHMM) ^7^, ATAC-seq profiles (29August2017_EJCsamples_allReads_500bp) ^47^, and Activity-by-Contact (ABC) scores (<u>AllPredictions.AvgHiC.ABC0.015.minus150.ForABCPaperV3.txt.gz</u>) ^48^, all generated from bulk immune cell populations. In all cases we lifted over the genomic coordinates from hg19 to GRCh38. For the PCHi-C data, enhancer–promoter interactions with a Chicago score ≥ 5 were retained within each cell type. ATAC-seq counts were normalised to log2 counts per million (CPM), and signals were partitioned into deciles within each cell type. Similarly, ABC scores were partitioned into deciles within cell type based on the ABC_Score metric. The correspondence between the source dataset cell types and CARDINAL PBMC populations is shown in **Table S29.** All the BP ChromHMM samples were from venous blood unless indicated otherwise.

#### Cell-type-specific eQTLs enrichment in particular classes of regulatory elements

To investigate the relationship between eQTL classification and functional genomic annotations, we extracted the most likely causal variant from each credible set at both fine-mapping resolutions (L0 and CT2) and intersected these variants with a curated functional annotation reference using matched cell-type annotations. For promoter capture Hi-C (PCHi-C) and Activity-by-Contact (ABC) datasets, overlaps were further classified as gene-matched when the annotated target gene corresponded to the eQTL eGene. Variants were stratified into high-confidence (maxPIP ≥ 0.9) and unfiltered (“No_Filter”) sets, which were analysed separately. For ABC and ATAC-seq datasets, decile-based annotations were additionally collapsed into composite high-signal categories (deciles 8–10) and, for ATAC-seq, low-signal categories (deciles 1–3) to improve statistical power and interpretability.

Overlap results were aggregated at the eQTL-module level and converted into a binary feature-presence matrix indicating whether a given annotation was observed in at least one matching cell type. Module counts were then summarised by eQTL specificity class, and enrichment of feature-positive modules among cell-type-specific eQTLs relative to all other eQTL classes was assessed using two-sided Fisher’s exact tests. P values were adjusted for multiple testing using the Benjamini–Hochberg false discovery rate (FDR) procedure within each PIP-filter group, with FDR < 0.05 considered statistically significant.

#### Enrichment of lineage-restricted eQTLs in matched lineage-restricted regulatory elements

To investigate the relationship between eQTL pleiotropy and regulatory element specificity, functional genomic annotations were collapsed into broad hematopoietic lineage categories according to the mapping shown in **Table S30**. Functional annotations were processed as described above, including gene-matched classifications for PCHi-C and ABC annotations and composite signal categories for ATAC-seq and ABC datasets. Fine-mapped eQTLs were represented by the most likely causal variant from each credible set and assigned to a hematopoietic lineage category based on their pleiotropy classification. Variant–annotation overlaps were identified while retaining the lineage classifications of both the eQTL and the overlapping functional element.

For each lineage category (PBMC, myeloid, lymphoid, and related subclasses), enrichment was assessed using 2 × 2 contingency tables comparing eQTL lineage specificity with the specificity of overlapping regulatory annotations. This framework tested whether eQTLs shared across lineages preferentially overlapped regulatory elements that were also shared across lineages, and whether lineage-restricted eQTLs preferentially localised to lineage-specific regulatory elements. Associations were evaluated using two-sided Fisher’s exact tests, and odds ratios with 95% confidence intervals were estimated. Confidence intervals were calculated using the Haldane–Anscombe correction to accommodate contingency tables containing zero counts. p-values were adjusted using the Benjamini–Hochberg FDR procedure within each annotation dataset, with FDR < 0.05 considered statistically significant.

#### TF Motif analysis of eSNPs

We used motifbreakR (2.24) ^49^ to annotate eSNPs predicted to alter TF binding activity. A list of 35,622 SNVs was created containing all eSNPs from joint fine-mapping, excluding indels, and analysed in chunks. The HOCOMOCO ^50^ (426 motifs)) database was chosen among available motifbreakR TF collections to avoid excessive redundancy of motifs. A p-value threshold of 0.001 was used to retain only motifs that had significant matches with PWMs, using the filterp=TRUE option. To define eSNPs with differential allelic effect on predicted motifs we selected those with effect defined as “strong”, resulting in 18,000 out of 35,622 variants with at least one predicted altered motif.

To identify TF motifs more likely to be causally linked to eQTLs we used Fisher exact test, comparing, for each motif the fraction of eSNPs with predicted alteration between eSNPs with PIP >0.8 and PIP ≤0.8. Significant motifs were defined at a FDR <0.1 (n=35). Individual motifs were assigned to TF subfamilies using the classification provided in the HOCOMOVO v14, motif clusters (https://hocomoco14.autosome.org/clusters). Enrichment results were further summarised by the TF subfamily (n=17) by keeping the motif in each subfamily with largest enrichment (lowest p-value and higher odds ratio).

To calculate enrichment for discordant pairs of motifs in “switching” eSNPs (eSNPs with discordant direction between cell types) we used Fisher exact test, counting instances where motif alteration involved at least two individual motifs in opposite direction, stratified by different annotations including PIP >0.8, overlap with ATAC peak and PIP >0.8 + overlap with ATAC peak. We performed the same analysis using as universe of variants either the 35,622 tested eSNPs or the 18,000 eSNPs predicted to alter at least one motif, returning similar results.

#### Enrichment in discordant effect cell types

To measure if the divergence of effect direction was preferentially happening between specific groups of cells more than chance, cell types were first mapped to their corresponding broad cell type. We then measured the correlation as the difference between the observed probability of co-occurrence of 2 cell types vs its expected probability by chance. This was then normalised by the variance of the two independent probabilities. This is analogous to the correlation measure between two alleles of different SNPs.

### Phenome-wide colocalisation analysis with PBMC sc-eQTLs

We downloaded SuSiE-inf fine-mapped credible sets from the Open Targets Platform ^29^ for GWAS Catalog studies (release 25.12) and FinnGen (release 25.09) ^51^ traits and transformed them into FLANDERS-ready AnnData objects to enable efficient large-scale colocalisation analysis. AnnData was annotated with disease locus coordinates, lead variants, colocalised genes, phenotype EFO mappings and therapeutic-area annotations. We then performed genome-and phenome-wide colocalisation analyses between PBMC single-cell *cis*-eQTLs and disease or quantitative-trait GWAS signals. We retained colocalisation events with evidence of a shared causal variant (PP.H4 ≥0.75). This resource served as the basis for downstream driver-cell-type inference, clinical trial support analyses, target safety assessment and drug repurposing analyses.

### Driver cell type identification

Disease-associated variants frequently colocalised with eQTL signals affecting the same gene across multiple PBMC cell types and, in some loci, across multiple genes simultaneously. To prioritise the most likely causal cellular context underlying each association, we developed a Mendelian-randomisation-inspired framework that quantified directional agreement between genetically inferred gene expression and disease risk across independent shared signals. For each colocalised gene–trait pair, we compared the direction and relative magnitude of disease and eQTL effects across all shared signals supporting colocalisation. We selected representative variants from each credible set and computed signed z-scores (β/SE) for both disease and eQTL effects. For loci containing multiple candidate genes, we performed the analysis independently for each gene-by-cell-type combination. To prioritise driver cell types, we transformed z-scores into inverse-rank weighted signed values, giving greater weight to high-confidence signals while preserving effect directionality. For each cell type, we evaluated two alternative regulatory models relative to the disease signal: an agonist model, in which increased gene expression aligned with increased disease risk, and an antagonist model, in which increased gene expression aligned with reduced disease risk. We calculated directional distances for both models as:

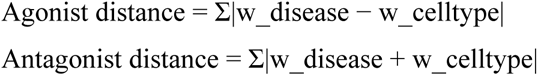

where w denotes inverse-rank weighted signed z-scores across shared signals.

We assigned regulatory direction according to the model with the smaller directional distance and designated the driver cell type as the PBMC population with the minimum directional distance across all candidate cell types. In loci containing multiple candidate genes, we applied the same criterion across all gene-by-cell-type combinations. This framework prioritises the cellular context in which genetically regulated gene expression most consistently parallels the disease-associated genetic effect. We additionally estimated the effect of gene expression in implicated cell types on disease risk using a Mendelian-randomisation-style framework. For each shared signal, we calculated a Wald-ratio estimate relating disease and eQTL effects and combined estimates across signals using inverse-variance weighting. These analyses yielded directional effect-size estimates for each gene–trait–cell-type combination, allowing us to quantify whether increased expression of a gene within a specific immune cell type was predicted to increase or decrease disease risk.

### Enrichment of eSNP categories with disease categories

Enrichment analysis of each eSNP hierarchical category and specific cell type with disease categories was performed using the fisher test of each cell type/category vs all others (coded as 1 = category/cell type, 0= other) and disease category vs all other disease categories. For biological categories and for fSNPs we also performed a different analysis estimating the probability of an eSNP to colocalise with a given disease category (coded as 1/0) given its biological category or it being an fSNP. For this we fit a logistic model:

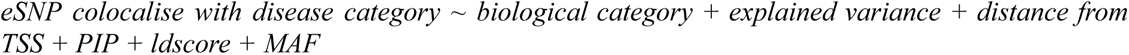

and

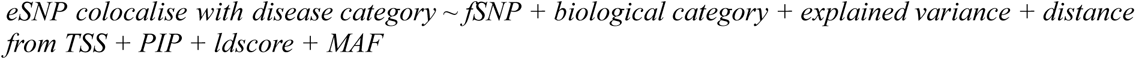

### Functional enrichment theme summarisation

Gene enrichment analysis was performed with enrichR ^52^ using the go terms biological processes. As input we used all eGenes which explained the loci colocalisation of each disease category that were explained only by CARDINAL compared to any other non-blood eQTL. This analysis identified enrichments only in the IMDs and Allergic categories. We then sought to highlight differences in enrichment terms themes in the two groups. Given the high number of terms in the results we used GPT-5.6 Sol to summarise the enrichment results into broader biological themes to reduce redundancy among closely related GO terms and facilitate comparison between disease clusters (**Table S31**). Enriched terms were assigned to one of seven predefined meta-themes using keyword-based annotation of the GO term description. The meta-themes comprised: shared T-cell activation and immune regulation; immune development and T-cell lineage specification; cytokine, interferon and JAK–STAT signalling; proliferation and immune effector function; negative regulation of T-cell responses; immune-cell survival and homeostasis; and costimulation and adhesion control. More specific categories were assigned before broader categories to minimise ambiguous classification of terms matching multiple keyword sets. Terms that did not match any predefined category were excluded from the theme-level comparison.

For each cluster and meta-theme, genes contributing to the corresponding enriched GO terms were extracted from the enrichment results. Duplicate genes occurring in multiple terms within the same meta-theme were counted only once. Theme representation was quantified as the proportion of unique enriched genes assigned to that meta-theme relative to the total number of unique genes represented across all enriched terms in the corresponding cluster:

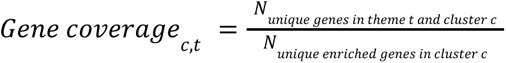

Differences in theme representation between clusters were calculated by subtracting the Allergic gene coverage from the IMD gene coverage. Positive values therefore indicated greater representation in the IMD cluster, whereas negative values indicated greater representation in the Allergic cluster. Themes with an absolute difference in gene coverage of no more than 0.03 were classified as broadly shared, while larger positive or negative differences were classified as IMD-biased or Allergic-biased, respectively.

### LIVI gene programme inference and *cis*–*trans* regulatory analyses

#### LIVI training

We trained LIVI ^32^ on the expression of 15,268 highly variable and/or highly expressed genes measured in 6,694,819 cells from 4,428 UKB donors of European ancestry. All cells were derived from fresh samples, but we performed correction for sequencing time and pool, as well as for donor sex, during LIVI training. Log-normalised gene counts were used to select 10,000 highly variable genes (HVGs) with scanpy (sc.pp.highly variable genes(adata, n top genes=10000, flavor=“seurat”)) and the 10,000 highest expressed genes. We used the union of those genes (n=15,268) to train LIVI. To facilitate genome-wide *trans*-eQTL discovery, we corrected for *cis*-eQTL effects at the single-cell level prior to training LIVI. Specifically, for all *cis*-eQTLs discovered by single-gene testing at the cell-type level, we re-estimated their effect sizes by averaging the expression of a given gene within the corresponding cell type. We then used these mean estimated effect sizes to correct the observed expression of each gene in each single-cell for the total *cis*-genetic effect of all SNPs associated with that gene in cells of the same type as follows: 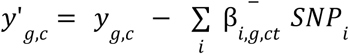 , where *y_g_*_,*c*_ is the original log-normalised expression of gene *g* in cell *c* , 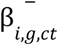 is the estimated effect size of SNP *i* on gene *g* in cell type *ct* (*c* ∈ *ct*) , and *y’_g,c_* is the *cis*-corrected log-normalised expression of gene *g* in cell *c* . We trained LIVI on the *cis*-corrected log-normalised expression by replacing the Negative Binomial likelihood of the original model with a Normal likelihood. We used 25 *C*, 1200 *D*, and 5 *V* factors.

#### Annotation of LIVI gene programmes

To annotate LIVI gene programmes (eSNP-associated *D* factors), we selected the top genes for each factor (i.e. genes with loadings more than 100 times above or below the interquartile range) and performed gene set enrichment analysis (GSEA) using Enrichr ^52^ as implemented in the `gseapy` package (version 1.1.9). We used all 15,268 genes that LIVI was trained on as the background set. The analysis was run separately for six databases: 1. Kyoto Encyclopedia of Genes and Genomes (KEGG) (as of 2026), 2. Reactome (as of 2024), 3. Gene Ontology (GO) Biological Process (as of 2025) and 4. GO Molecular Function (as of 2025), 5. Elsevier Pathway Collection, and 6. Hallmark gene sets from the Molecular Signatures Database (MSigDB) (as of 2020). For each database, we retained terms with an adjusted *p*-value < 0.05. When multiple terms were significantly enriched within a database, we selected the term involving the largest number of genes; if several terms involved an equal number of genes, we chose the one with the smallest adjusted *p*-value and highest odds ratio. We then manually curated the selected annotations into hierarchical groups. Specifically, we created 37 non-overlapping groups based on shared biological function: 22 immune signaling categories (e.g., *T Cell Biology, NK Cell Biology, Interferon Signaling, Pattern Recognition Receptors, Antigen Presentation & MHC)* and 15 disease-associated categories (e.g., *Autoimmune Disease, Viral Infection & Host Defense, Haematological Malignancies*). Assignment was performed manually, guided by canonical immunological classification. For instance, pathways involving Toll-like and NOD-like receptors were grouped under *Pattern Recognition Receptors*, while interleukin-specific signaling cascades (e.g., IL-6/JAK/STAT3, IL-17) were grouped under *Interleukin Signaling*, separately from broader cytokine signaling pathways. Disease-contextualised immune terms (e.g., *Th1 Cell Activation in Crohn’s Disease*) were assigned to their respective disease category rather than the underlying immune mechanism. For visualization purposes, pathway display names were harmonised by removing GO accession suffixes and converting fully capitalised terms to title case while preserving standard abbreviations (e.g., MHC, IgA, TNF, NF-κB). Full GSEA results, as well as the complete mapping of factor annotations to biological categories are provided respectively in **Tables S32 and S14**.

#### Assignment of LIVI gene programmes to discrete cell types

LIVI factors have continuous values across cells; however, they can also be assigned to known discrete cell types using a two-step approach. First, we use LIVI’s assignment matrix, **A** ^32^, to select cell-state (*C*) factors relevant to each *D* factor (**A** ≥ 0.6). Second, we map cell-state factors to known cell types by averaging the cell-state factor values across cells of each type, and subsequently selecting the cell type with the highest mean value for a given cell-state factor. This yields the cell type(s) in which a given factor is primarily active. Nevertheless, for the disease-specific *cis*-trans graphs, we also took into account secondary cell types in which the factor is active. To include these, we relaxed the maximum mean value criterion and instead selected all cell types with a mean value greater than one standard deviation above the mean for the given cell-state factor. This is a meaningful relaxation given that a single cell-state factor may represent closely related cell types for which the difference between the mean values is negligible.

#### Association testing of LIVI programmes

GWAS of 1,200 LIVI programmes across 4,428 donors were conducted using REGENIE (v4.0) implemented via the nf-pipeline-regenie pipeline (https://github.com/HTGenomeAnalysisUnit/nf-pipeline-regenie). Single-variant association testing was performed using REGENIE Step 2, with Step 1 model fitting omitted. Association models included sex and 20 genetic principal components (PCs) to account for population stratification. Prior to GWAS, LIVI factors were inverse rank–normal transformed using REGENIE’s --apply-rint option to improve normality and stabilise variance. Variants were filtered using a minor allele count (MAC) threshold of 10 and a minor allele frequency (MAF) threshold of 1%. GWAS summary statistics were harmonised and ingested into a TileDB database and subsequently fine-mapped using the FLANDERS workflow, applying the loci boundaries already identified in the *cis*-eQTL analysis.

To minimise the number of multiple tests and computational burden we focused only on the eQTL loci whose top associated SNP showed a significant association (FDR<0.05) with at least one of the LIVI-Factors (7,254 loci). Credible sets having at least one SNP with conditional p-value < 2x10^-5^ were then colocalised with *cis*-regulated gene expression modules.

#### Enrichment of cross-cell type *cis*–*trans* colocalisations for intercellular communication

We tested whether colocalised *cis*-regulated modules and LIVI gene programme pairs acting across different cell types could be explained by cell-cell communication. For this analysis, we focused on colocalisation events involving *cis*-eGenes encoding for known ligands. Because ligand-receptor databases describe surface interactions but not the downstream genes a signal regulates, we used the NicheNet ^53^ prior model, which integrates ligand-receptor, signal-transduction, and gene-regulatory networks into a ligand-target regulatory-potential matrix (∼1,200 ligands × ∼25,000 target genes). For each association we treated the *cis*-eGene as the candidate ligand and the LIVI programme genes as the gene set of interest, yielding 686 unique ligand–LIVI programme pairs regardless of cell type. For these pairs, we used permutation tests to assess whether the LIVI programme was more enriched for the colocalising ligand’s targets than expected by chance compared with a random gene set of the same size. Specifically, first we took the intersection of the NicheNet background and the genes we used to train LIVI to define a common gene universe. Within this universe, we computed the ligand’s average precision of its NicheNet regulatory-potential vector against binary LIVI programme membership. We compared this to a null distribution of average precisions obtained from 10,000 random gene sets of the same size drawn from the same universe 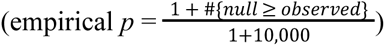.

Because the null average precision depends only on gene-set size and not on the ligand, it was computed once for gene-sets of the same size. *P*-values were adjusted (Benjamini-Hochberg) across unique ligand-programme pairs appearing in at least one cross-cell-type association.

#### Enrichment of LIVI gene programmes for TF target genes

To assess whether colocalisations between *cis*-eQTLs and LIVI gene programmes reflect TF-mediated trans regulation, we tested (i) whether cis-eGenes colocalising with a LIVI programme are enriched for TFs relative to the full set of discovered *cis*-eGenes, and (ii) whether, among colocalisations whose *cis*-eGene is a TF, the LIVI programme contains the TF’s known downstream targets more often than expected by chance. TF regulons were obtained from CollecTRI ^33^, a literature-curated resource covering 1,189 TFs with 43,536 unique TF–target edges.

To assess overall TF over-representation among colocalising *cis*-eGenes, we used a one-sided Fisher’s exact test (alternative = “greater”) to compare TF representation in *cis*-eGenes colocalising with LIVI programmes vs. *cis*-eGenes not colocalising with LIVI programmes. The analysis was restricted to *cis*-eGenes with known HGNC symbols (13,603 out of the 15,419). Of the 2,033 *cis*-eGenes colocalising with LIVI programmes, 124 (6.1%) were TFs and 1,909 were not, whereas of the 11,570 *cis*-eGenes not colocalising with any LIVI programme, 577 (5.0%) were TFs and 10,993 were not.

To assess enrichment of TF-target genes in colocalising LIVI programmes, we intersected CollecTRI regulon genes with the 15,268 genes we used to train LIVI. We also reduced the colocalisation events to 640 unique TF-LIVI programme pairs, each representing a distinct TF-LIVI programme colocalisation regardless of the underlying eSNPs and cell types. We computed the number of pairs for which the colocalising LIVI programme contained at least one of the known TF target genes and compared this to the number we would expect by chance. To obtain the null distribution of LIVI programme-regulon overlap, we shuffled the TFs in the TF-LIVI programme pairs 10,000 times. This preserves the marginal distributions of regulon size, LIVI programme size, and gene frequency across LIVI programmes, and breaks only the specific TF-LIVI programme pairing being tested. One-sided p-values were computed using the conservative approximation of the Phipson & Smyth exact permutation p-value: 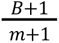, where *B* is the number of permutations yielding a test statistic at least as extreme as the observed value and *m* is the total number of permutations (here 10,000).

#### Comparison of cell-type-autonomous and cell-type non-autonomous edges

Unique *cis*-*trans* eQTL edges were defined as distinct Source_module–Source_gene–program–Target_gene–edge.type combinations (n = 83,918 edges) and classified as cell-type autonomous (Within-only: both eGenes active within the same cell type) or cell-type non-autonomous (Across-only: eGenes active in different cell types); no edges were observed in both categories (Mixed). Edges were annotated against eight gene-regulatory and protein-interaction resources: CollecTRI ^33^ (transcription factor–target pairs supported by ≥2 corroborating sub-resources), SCENIC+ ^54^ regulon activator and repressor networks derived from CARDINAL, POSTAR3 ^55^ (RNA-binding protein–target pairs passing a method filter of ≥2 supporting methods with ≥1 high-confidence method among CLIPper/PureCLIP/PARalyzer), the Liana consensus resource ^56^ (ligand–receptor pairs), MEROPS ^57^ (peptidase–substrate pairs restricted to physiological cleavage), and STRING ^58^ noisy-OR rescaled to isolate the protein protein interaction experimentally validated channel at two confidence bands (0.1 < score ≤ 0.4 and score ≥ 0.4).

For each resource, enrichment of annotated edges among cell-type autonomous relative to non-autonomous edges was tested with Fisher’s exact test (odds ratio > 1 indicates enrichment among autonomous edges), under three complementary definitions of a “hit”: (i) the edge’s gene pair is present in the resource irrespective of orientation; (ii) the *cis*-eQTL source gene occupies the regulator role (TF, RBP, ligand, or peptidase); (iii) the *cis*-eQTL source gene occupies the target role (TF-target, RBP-target, receptor, or substrate). STRING, being undirected, was tested only under (i). P-values from all tests across all resources and directions were corrected jointly using the Benjamini–Hochberg procedure.

#### Visualisation of cell-type autonomous and cell-type non autonomous *cis*/*trans* networks

*Trans*-eQTL direction/magnitude (MR effect) was merged onto each edge; a single LIVI programme-level effect magnitude is shared across a hub’s targets, with sign varying per target. Edges are colored by MR sign (negative/positive), and spoke ordering follows MR sign block with cascading TFs clustered within each block. Direct topological annotation marks edges with direct support from SCENIC/CollecTRI/STRING/Liana; cascading annotation marks edges reached through an intermediate regulator, using the resource-agnostic pairwise cascade detector applied to a factor’s *trans*-eQTL targets (excluding the hub gene) against CollecTRI, SCENIC regulons and Liana Ligand-receptor annotations.

#### Definition of disease specific *cis*-*trans* graphs

To define disease specific *cis*-trans regulation networks we transformed the *cis*-eGene to LIVI programme relationships into a *Source gene in cell type* to *Target gene in cell type* network. First, we examined whether the cell types involved in the colocalising *cis*-eSNP overlapped with the programme cell types; if this was the case we assumed that the *cis*-*trans* regulation was happening within these cell types and edges were created between all *Source gene in cell type* (according to the *cis*-eSNP) and all genes x cell types in the programme. If no overlap was observed between the cell types of the *cis*-eSNP and those of the programme a similar procedure was applied, but using all cell type combinations involved. This created a graph made of 292,886 edges including 2,885 source traits (*cis* eGenes in cell types) and 24,087 target traits For each edge, we estimated the effect of the source gene on the target gene in two steps. First, we estimated the causal effect of the *cis*-eGene on the LIVI programme using the Wald ratio, with the lead *cis*-eSNP as the instrument. Second, we multiplied this estimate by the sign of the loading of the gene onto the program weighted by the correlation between the target *trans*-regulated gene’s expression and the programme value. This step assigns each target *trans*-regulated gene the appropriate direction of effect while preserving the relative magnitude of effects across genes within a programme differently across cell types. For each disease, we retained edges in which both the source and the target gene colocalised with the disease (PP.H4 ≥ 0.75). For the source *cis*-eGene, colocalisation was required at the same *cis*-eSNP used to estimate the *cis*–*trans* effect, while for the target *trans*-regulated gene, colocalisation was assessed using its own *cis*-eQTLs from the previous analysis (section **Phenome-wide colocalisation analysis with PBMC sc-eQTLs**). Edges meeting these criteria were identified for 60 diseases (Fig. S18). For each retained edge, we computed the expected effect of the source *cis*-eGene on disease mediated through the target trans-regulated gene as the product of the predicted effect of the *cis*-eGene on the *trans*-regulated gene and the observed (MR) effect of the target *trans*-regulated gene on disease (analysis described in section **Driver cell type identification**). We compared this expected mediated effect with the observed effect of the source *cis*-eGene on disease. Because *cis*–*trans* effect sizes are not on a common scale with disease effects, we assessed only the concordance of effect directions, testing whether the proportion of concordant edges differed from the 50% expected by chance using a binomial test.

### Rare variant analysis

#### WGS data processing

Genotype-level quality control was performed on the 154,430 project variant call format files (pVCFs) provided by UK Biobank using bcftools v1.15, applying criteria adapted from Hawkes et al. ^59^. Genotypes were set to missing if any of the following conditions were met: (i) local allele depth (LAD) <10 per sample genotype, (ii) genotype quality (GQ) <10, or (iii) heterozygous calls with binomial test on allelic depth *p*-value ≤0.001 (binom(FMT/LAD) ≤0.001). Variants were subsequently excluded if they had >10% missingness or deviated from Hardy–Weinberg equilibrium (HWE *p*-value ≤1×10^-15^).

#### Rare variant burden computation

Rare variant gene burden analyses were performed using multiple annotation tools for variant weighting. We included variants with minor allele frequency (MAF) <0.01 based on the CARDINAL subset of UK Biobank. Variants within the gene body and ±5 kb flanking regions were defined using GENCODE v44 ^60^ and annotated with Ensembl Variant Effect Predictor (VEP) v113 ^61^. Functional annotations for burden computation were obtained from CADD v1.7 ^62^, SpliceAI ^63^, LOFTEE ^64^, 5′ UTRAnnotator ^65^, (Tissue = Whole Blood ^66^), and PromoterAI ^67^. For each gene, individual, and annotation, the burden score was defined as the sum of the annotation scores of all rare variants in the gene body and flanking regions. For binary annotations (LOFTEE and 5′ UTR consequences), burden scores was the count of variants carrying the corresponding consequence.

#### Rare variant eQTL testing

Gene-level burdens were tested for association with cell type–specific expression using the same framework as the common-variant *cis*-eQTL analysis, differing only in the tested predictor. Specifically, we used identical expression phenotypes — the per–cell type pseudo-mean expression values, normalised (PFlog1pPF), aggregated across cells and inverse-rank-normal transformed exactly as described for the common-variant analysis — and the identical covariate set (sex, 20 genetic PCs and 20 expression PCs), replacing the SNP genotype/dosage with the per-individual gene-level burden as the tested predictor (adapted TensorQTL implementation). Each gene’s burden was tested only against the expression of that same gene. Only (gene, annotation) pairs with a cumulative allele count > 5 that also passed the common-eQTL expression filter (expressed in ≥ 50 individuals and in ≥ 20% of individuals) were included. Multiple-testing correction was performed with the Benjamini–Hochberg method separately within each cell type and annotation, and associations with FDR < 5% were considered significant. Analyses were restricted to third-degree unrelated individuals of European ancestry (n = 3,984; UKB Field 22006). To confirm calibration, we permuted the sample labels of the expression matrix once and re-ran the full association procedure, yielding one null test per gene for each burden mask and cell type. Calibration was assessed by comparing the resulting p-value distribution to the uniform expectation. All analyses were performed using the cellink package (https://cellink-docs.readthedocs.io/en/latest/).

#### Characterisation of rare variant specific signals

Intersecting eQTL results from common variant and rare variant analysis, we identified a set of eGenes discovered only by rare variant analysis in a specific cell type (novel in cell type) and a set of eGenes exclusively identified by rare variants tests, but never observed in any cell type in common variant analysis (novel genes). To characterise these gene sets, we first annotated LOEUF and pLI constraint values for each gene using gnomAD v4 ^68^ annotations and we used Kruskal-Wallis test to compare the LOUEF value distribution across shared, novel in cell type, novel genes. We then used Fisher’s exact test to assess over-representation of essential genes in the novel genes set. Essential genes were defined according to 3 different criteria, namely pLI > 0.9, LOEUF < 0.362 (10th percentile) or present in the DEG database annotations ^69^. Finally, we annotated existing relationships between rare variant novel genes/proteins using STRING DB v12.0 ^70^. From the resulting network, we extracted connected sub-components and annotated them by manual curation of the genes function to identify key cellular processes involved by rare variant novel eGenes.

#### Rare variant phenotype association testing in the full UK Biobank cohort

Genes with a burden association passing FDR < 5% in the discovery analysis were intersected, on the gene, with the common-variant eQTL–GWAS Catalog colocalisations (described above) to define the (burden mask, gene, phenotype) triples carried forward for testing in the full UK Biobank cohort (N = 463,072, restricted to European ancestry). This yielded 2,104 triples for quantitative traits (381 unique genes, 248 phenotypes) and 160 triples for disease traits (98 unique genes, 26 phecodes). GWAS Catalog phenotypes were mapped to their corresponding UKB equivalents using PhecodeX ^71^ for clinical disease traits and UKB Showcase fields for quantitative traits. Binary case–control indicators for the 26 disease traits were derived from UKB ICD-10 codes using the PhecodeX mapping framework.

For each gene and burden mask, gene burdens were computed for all 463,072 UKB participants of European ancestry as described above, here restricting to variants with MAF < 0.1% (0.001), and association testing was performed using REGENIE ^72^. In REGENIE Step 1, we used 483,446 imputed SNPs (UKB Field 22828) filtered for MAF > 0.01, MAC > 100, genotyping rate > 0.99 and HWE p ≥ 1×10^-15^, and pruned for linkage disequilibrium (r² < 0.9; 1,000-unit window, 100-unit step). In Step 2, each colocalised (gene, burden mask, phenotype) triple was tested: the 248 quantitative phenotypes were rank-inverse-normal transformed, and binary traits were tested using the Firth likelihood-ratio test. Triples with fewer than five cases (binary) or fewer than five measured individuals (quantitative) carrying a variant contributing to the burden mask were discarded. All models included age, age², genetic sex, age × sex, and the first 20 genetic PCs as covariates. Benjamini–Hochberg FDR correction was applied separately within each phenotype category (quantitative and binary), and associations with FDR < 5% were considered significant.

#### Identification of allelic series

The results from the PheWas rare variant analysis were intersected with the colocalisation results between CARDINAL modules and Open Targets GWAS traits from GWAS catalog giving us 3,345 variants. A direction of effect was built for rare variants similarly to the colocalisation results, by taking the ratio between the beta from the GWAS and the beta from the PheWas analysis. We later filtered for variants that had an adjusted p-value ≤ 0.05 from PheWas analysis. Finally, we checked for variants that shared the same direction of effect between PheWas and colocalisation analyses with respect to quantitative and binary traits.

### Therapeutic target prioritisation and validation

#### scRNA-seq eQTL support

We evaluated whether genetically supported targets identified from PBMC eQTL colocalisation analyses were enriched among successful drug-development programmes using historical drug-development annotations from the Open Targets Platform (release 25.09). Drug–target relationships, indication EFO identifiers and maximum clinical trial phase annotations were obtained from Open Targets drug records, and disease ontology annotations were obtained from the corresponding disease dataset.

A drug was considered genetically supported if at least one of its target genes overlapped a colocalised eGene. Indication-matched support additionally required exact matching between the drug indication EFO identifier and the colocalised trait EFO identifier. We excluded oncology drugs using disease ontology annotations containing cancer-related terms.

We summarised drug-development progression using maximumClinicalTrialPhase and defined progression endpoints as entry into clinical testing (≥0.5), progression beyond phase I (≥2), progression beyond phase II (≥3) and regulatory approval (≥4). For each endpoint, we evaluated associations between genetic support and drug-development progression using two-sided Fisher’s exact tests applied to 2 × 2 contingency tables and reported odds ratios with 95% confidence intervals.

#### Target safety assessment and classification

We assessed the phenome-wide safety profile of PBMC sc-eQTL supported targets by evaluating the consistency of inferred gene-expression effects across all colocalised traits. This analysis included both disease phenotypes and ten quantitative biomarker traits: fasting glucose, LDL cholesterol, systolic blood pressure, body mass index, estimated glomerular filtration rate (eGFR), alanine aminotransferase, aspartate aminotransferase, alkaline phosphatase, gamma-glutamyl transferase, and C-reactive protein. For each colocalised gene–trait pair, we classified increased gene expression as predisposing when it aligned with increased disease risk or adverse biomarker profile, and as protective when it aligned with reduced risk. Biomarker polarity was curated according to established clinical interpretation. For all biomarkers except eGFR, higher trait values were considered adverse. We then classified genes according to the consistency of their inferred effects across traits. Directionally concordant targets showed uniformly risk-increasing or uniformly risk-decreasing effects across all colocalised traits, indicating no genetic evidence of opposing on-target effects. Directionally discordant targets showed both risk-increasing and risk-decreasing associations, suggesting potential pleiotropic trade-offs associated with therapeutic perturbation. We excluded genes colocalising with only a single trait because they lacked sufficient information for phenome-wide consistency assessment.

#### Drug repurposing

We identified potential drug repurposing opportunities by integrating disease–gene colocalisation results with approved drug–target relationships from the Open Targets Platform (release 25.09), restricting the analysis to non-oncology approved drugs. Drug–target relationships, mechanism-of-action annotations and registered disease indications were obtained from Open Targets drug records.

We prioritised repurposing candidates based on evidence of disease–gene colocalisation and the presence of approved drugs targeting the encoded protein. We then integrated driver-cell-type assignments and inferred regulatory direction to get a predicted therapeutic effect of upregulation of target’s gene expression.

To evaluate therapeutic alignment, we integrated mechanism-of-action annotations with genetically inferred regulatory direction. We classified colocalised gene–trait pairs as risk-increasing when increased genetically inferred gene expression was associated with increased disease risk, and as risk-decreasing when increased expression was associated with reduced disease risk. Downregulating mechanisms, including inhibitors, antagonists, blockers, inverse agonists and degraders, were considered therapeutically aligned for risk-increasing targets and discordant for risk-decreasing targets. Conversely, upregulating mechanisms, including agonists, activators and positive modulators, were considered aligned for risk-decreasing targets and discordant for risk-increasing targets. Combinations with insufficient mechanistic or directional information were classified as unresolved.

We assigned GWAS traits and drug indications to therapeutic areas using Open Targets ontology annotations and prioritised repurposing opportunities where the colocalised disease differed from the approved indication of the drug. We then integrated driver-cell-type assignments, therapeutic alignment and target safety classification.

We performed an LLM-assisted evidence review using recorded drug indications, selected regulatory sources and web searches to identify existing indication overlaps and prior drug-, target- or pharmacological-class evidence. Combinations were classified according to existing approval, direct or related human investigation, preclinical drug evidence, target/class treatment precedent, target-level biological evidence, or absence of identified precedent. We assessed whether the evidence concerned the proposed disease, accounting for synonyms, subtypes and differences in treatment setting or endpoint. Adverse-direction evidence and target or modality mismatches were recorded as review concerns. Supporting findings, evidence scope, sources and review dates were retained in **Table S28**. Prior-evidence classification was assessed independently of therapeutic alignment. Additional indication candidates comprised therapeutically concordant combinations after excluding existing drug approvals and broad phenotypes overlapping approved indications. Combinations for which no prior evidence was identified were classified as potential repurposing opportunities.

## Supporting information

Supplementary Note FLANDERS methods and benchmarking

## Acknowledgements

This research was funded in part by the Wellcome Trust [grant number 220540/Z/20/A], the Chan Zuckerberg Foundation [grant number 2020-224316] and by the British Heart Foundation (BHF) Cambridge Centre for Research Excellence RE/18/1/34212. Fondazione Human Technopole is supported by public funds awarded by the Italian Ministry of Economy and Finance, the Ministry of University and Research, and the Ministry of Health.

This work is supported by the project IMPACT (G.A.N.) 101201772 Funded by the European Union. Views and opinions expressed are however those of the author(s) only and do not necessarily reflect those of the European Union or grant authority (ERCEA). Neither the European Union nor the granting authority can be held responsible for them.

This research has been conducted using the UK Biobank Resource under application number [69938].

We thank Wellcome Sanger Institute scientific operations team members; Stefanie Lensing, Scott Thurston, Lesley Shirley, Sameena Iqbal, Katy Taylor, Cordelia Langford and Ian Johnston for automation, sample management and sequencing operations support. Wellcome Sanger Institute Human Genetics Department team members; Sally Bygraves, Nicola Corton, Kalia Dede, Guillaume Noel and Hannes Postingl.

Genes & Health is/has recently been core-funded by Wellcome (WT102627, WT210561), the Medical Research Council (UK) (M009017, MR/X009777/1, MR/X009920/1), Higher Education Funding Council for England Catalyst, Barts Charity (845/1796), Health Data Research UK (for London substantive site), and research delivery support from the NHS National Institute for Health Research Clinical Research Network (North Thames). We acknowledge the support of the National Institute for Health and Care Research Barts Biomedical Research Centre (NIHR203330); a delivery partnership of Barts Health NHS Trust, Queen Mary University of London, St George’s University Hospitals NHS Foundation Trust and St George’s University of London. Genes & Health is/has recently been funded by Alnylam Pharmaceuticals, Genomics PLC; and a Life Sciences Industry Consortium of AstraZeneca PLC, Bristol-Myers Squibb Company, GlaxoSmithKline Research and Development Limited, Maze Therapeutics Inc, Merck Sharp & Dohme LLC, Novo Nordisk A/S, Pfizer Inc, Takeda Development Centre Americas Inc. We thank Social Action for Health, Centre of The Cell, members of our Community Advisory Group, and staff who have recruited and collected data from volunteers.

We thank: Barts Health NHS Trust, NHS Clinical Commissioning Groups (City and Hackney, Waltham Forest, Tower Hamlets, Newham, Redbridge, Havering, Barking and Dagenham), East London NHS Foundation Trust, Bradford Teaching Hospitals NHS Foundation Trust, Public Health England, Discovery Data Service/Endeavour Health Charitable Trust (especially David Stables), Voror Health Technologies Ltd, NHS England - for GDPR-compliant data sharing backed by individual written informed consent. Most of all we thank all of the volunteers participating in UK Biobank and Genes & Health.

## Author Contributions

A.L., M.O., S.S., A.R., B.A., M.J.B. and N.P. performed eQTL analyses, with M.J.B. and N.P. as co-leads.

N.P., S.S., M.T.S., P.B., E.G. performed the eQTL functional and biological annotations.

A.L., S.S, B.A, E.G. and N.P. built the FLANDERS pipeline.

S.S. and N.P. developed the methods used in the FLANDERS pipeline.

A.L. performed testing and benchmarking for methods implemented in the FLANDERS pipeline.

B.A. developed TileDB-sumstat for storage and querying of association summary statistics.

D.V., N.P. and M.T.S. performed the LIVI analysis and consequent biological interpretation.

NP and M.T.S. performed the disease specific network analysis and biological annotation.

S.S. performed therapeutic target prioritisation and validation.

E.H., E.G. and B.A. performed rare variants analyses.

C.C., M.I.C., I.G., C.D.C., M.C., A.G., V.S., F.S., D.Z., S.W., O.D, J.P.E., G.C., D.B., L.A., B.C., S.L.,

A.C., E.R, R.E., A.S, D.V.S. E.C., M.H., G.I, F.T., N.Subasinghe., L.D, B.S., C.A.G., N.Staudt, J.W.,

R.T, J.G., S.F, P.F.C., F.C., D.A.v.H assisted with interpretation of results.

S.W., N.Staudt., V.I., S.A.T., M.J.B., O.S., D.A.v.H., N.P., and N.Soranzo designed and coordinated the project.

J.W., R.T., and S.F. provided research resources.

V.I., F.J.T., S.A.T., O.S., D.A.v.H., and N.Soranzo supervised the study.

N.P.*#, S.S.*, D.V.*, E.H.*, A.L.*, M.O.*, E.G.*, M.T.S.*, P.B.*, F.T., B.C.#, O.S#., V.I.#, M.J.B.#,

S.A.T.^#^, and N.Soranzo^#^ wrote the manuscript.

N.P.*#, S.S.*, D.V.*, E.H.*, A.L.*, M.O.*, E.G.*, M.T.S.*, P.B.* contributed equally to this work. All authors reviewed and approved the final manuscript.

## Additional Information

Supplementary Information is available for this paper. Correspondence and requests for materials should be addressed to Nicole Soranzo), Marc Jan Bonder, Nicola Pirastu), Sarah A. Teichmann, Oliver Stegle.

## Data availability

- The genotype and phenotype for the UKB cohort described in this study were obtained from the UK Biobank resource under approved application number 69938. These are available, together with raw and processed single-cell data, to bona fide researchers upon approved application to the UK Biobank Access Management System.
- Fine-mapped credible sets for FinnGEN, GTEx and bulk eQTLs datasets were downloaded from OpenTarget Platform v25.09; Fine-mapped credible sets for GWAS Catalog were downloaded from OpenTarget Platform v25.12 (https://platform.opentargets.org/).
- Functional genomic annotations used for enrichment analyses, Promoter Capture Hi-C, BLUEPRINT ChromHMM, ATAC-seq and Activity-by-Contact (ABC) scores are available as cited in Methods and the original publications.

## Code availability

Code for the TileDB workflow used to store sc-eQTLs summary statistics and run locusbreaker algorithm is available via GitHub at https://github.com/HTGenomeAnalysisUnit/TileDB-sumstat/tree/nextflow_branch. Code for the FLANDERS workflow used to perform fine-mapping, to create cis-genetic modules and perform colocalisation is available via GitHub at https://github.com/Biostatistics-Unit-HT/Flanders. Code for generating fine-mapping data from Open Target Platform credible set data is available via GitHub at https://github.com/Biostatistics-Unit-HT/Flanders/blob/main/bin/parse_credible_sets_refined.py. Code for the inference of multicellular gene programmes and *trans*-eQTL associations using LIVI is available via Github at https://github.com/PMBio/LIVI. Code for the nf-pipeline-regenie pipeline used for GWAS of LIVI gene programmes is available via GitHub at https://github.com/HTGenomeAnalysisUnit/nf-pipeline-regenie. The cellink package is available via GitHub at https://github.com/theislab/cellink. Custom code utilised in this study is available via GitHub at https://github.com/Biostatistics-Unit-HT/Pirastu-et-al-sc-eQTLs-immune-regulatory-architecture.

## Consortia

### Genes & Health Research Team

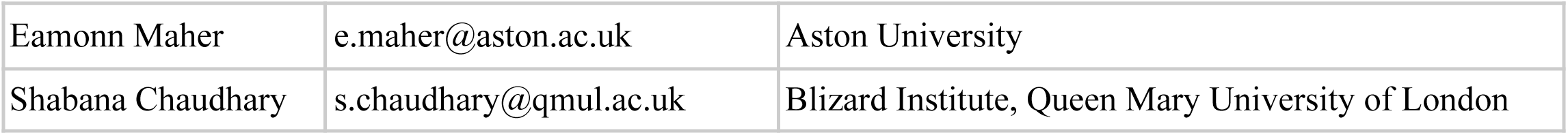
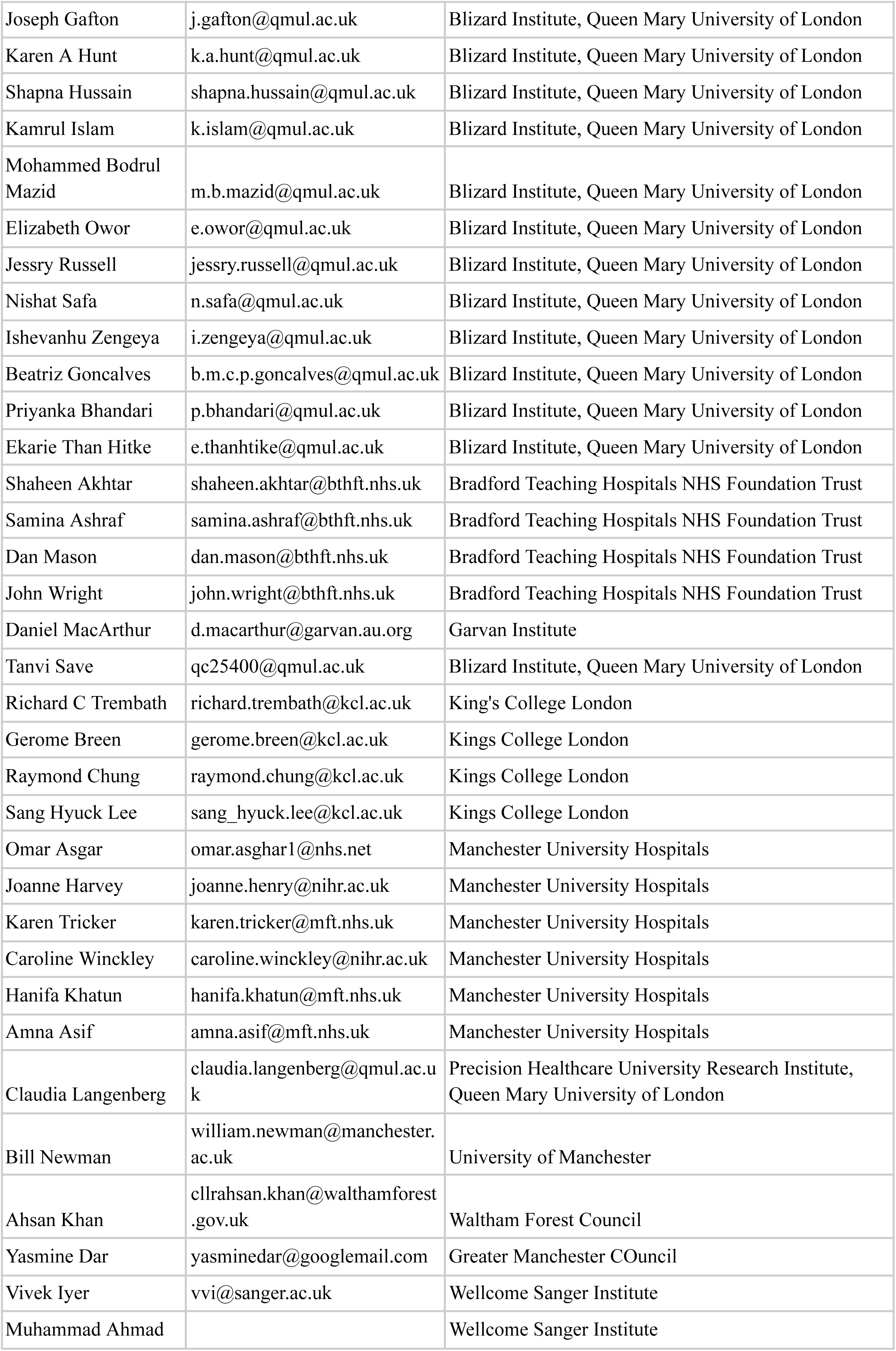
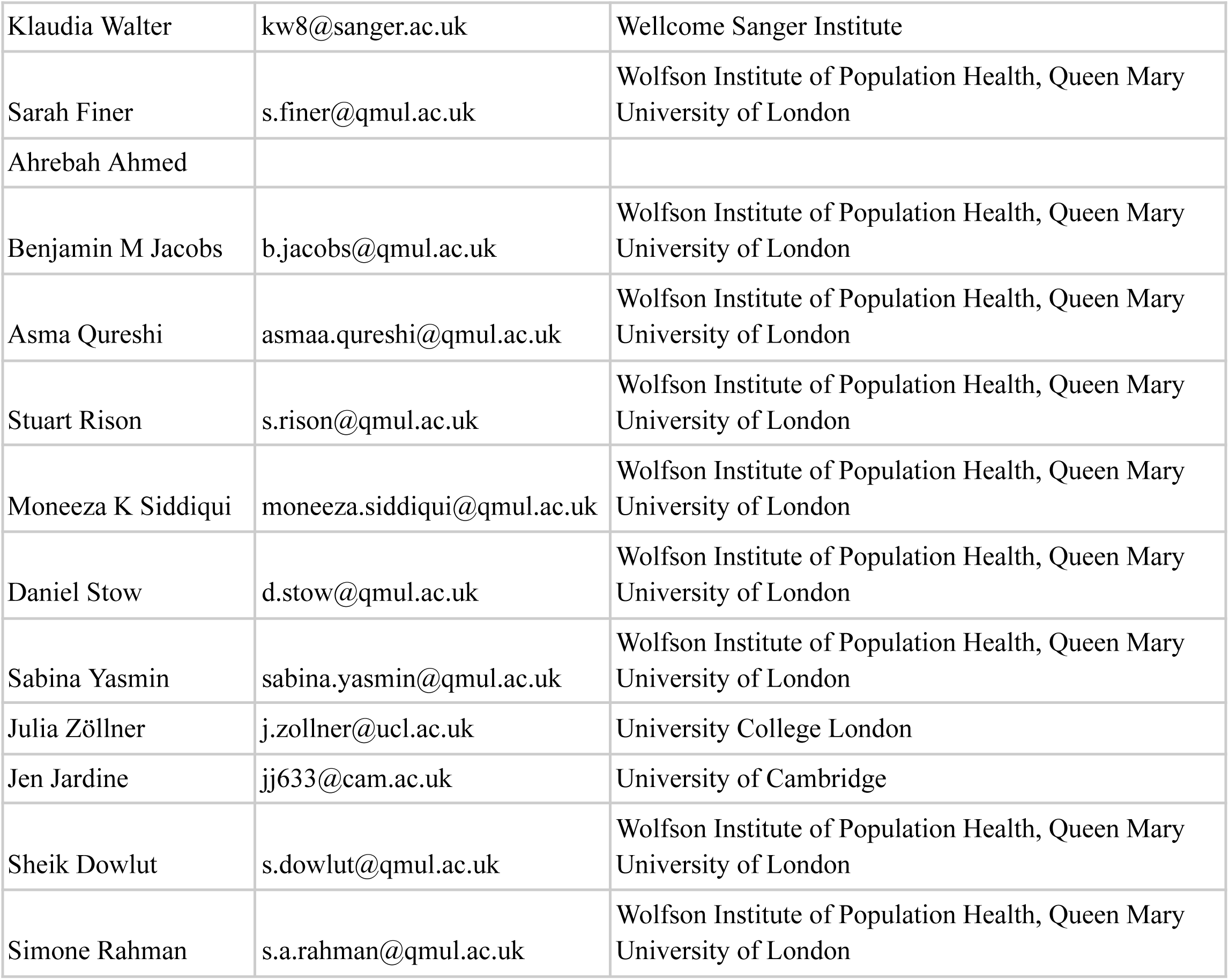

