## Supplementary figures for "Hierarchical genetic regulatory architecture of the human immune system informs complex disease mechanisms"

### Supplementary figures and legends

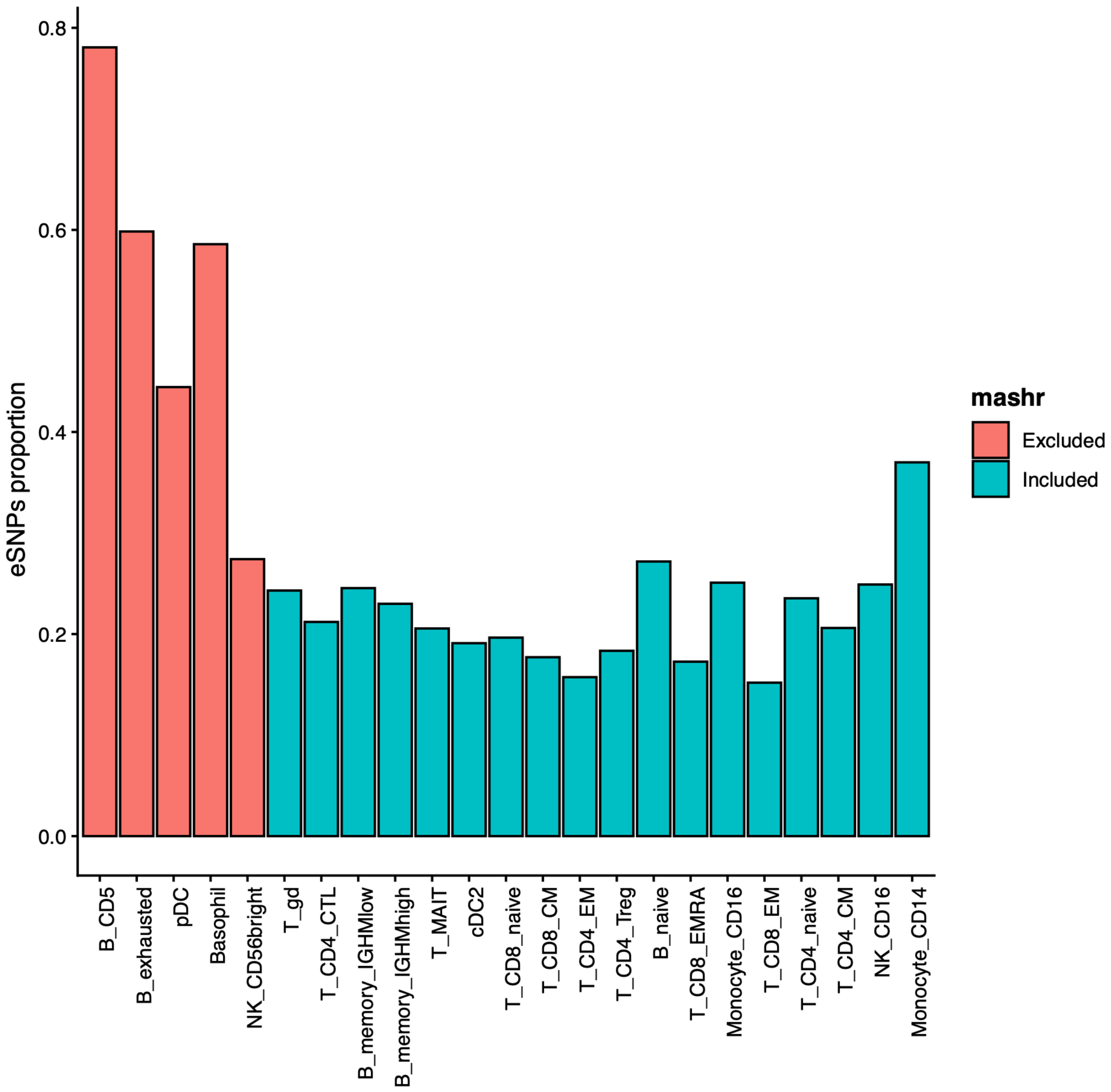

**Supplementary Figure S1 | Cell-type specificity of eSNPs following mashr-based assessment of effect sharing.**

Proportion of eSNP classified as cell-type-specific in each cell type after excluding those for which mashr identified evidence of shared effects across cell types ("apparently specific eSNP"), Colours indicate cell types included in the mashr analysis (turquoise) or excluded from it (red).

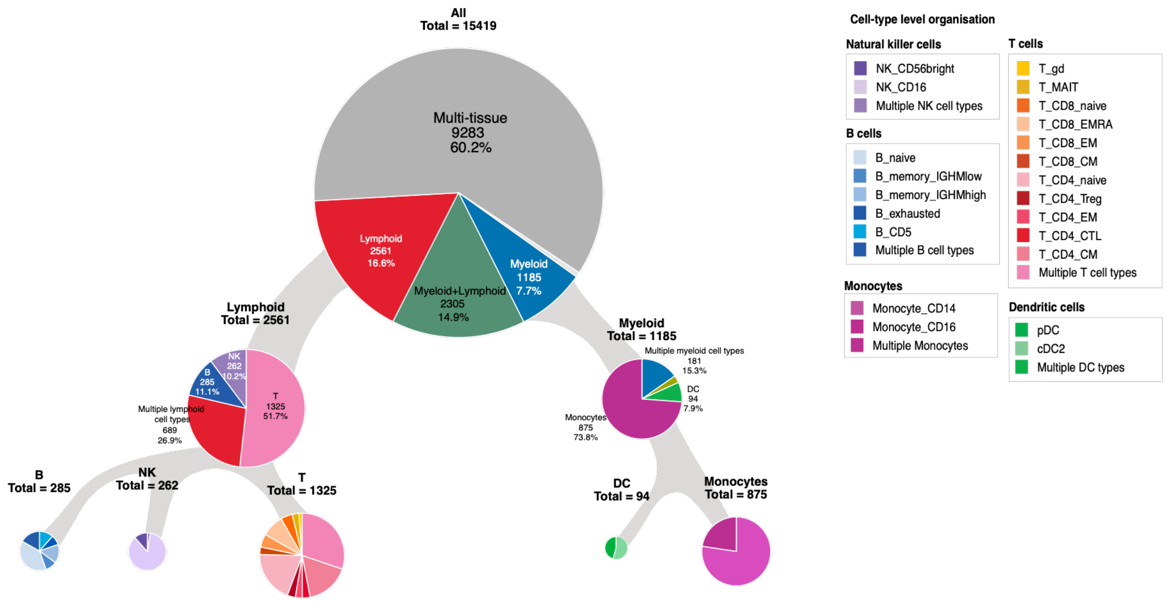

**Supplementary Figure S2 | Proportion of specific eGenes per cell type category.**

Proportion of eGenes across progressively finer categories of cell-type specificity, from broad lineage groups to individual cell types. Counts are cumulative within each branch: for example, the T-cell category at the second level includes both eGenes shared across multiple T-cell types and also those specific at the lower T-cell levels. Sectors labelled ‘multiple’ represent eGenes shared across cell types within the indicated group. Numbers and percentages refer to the total at the corresponding level.

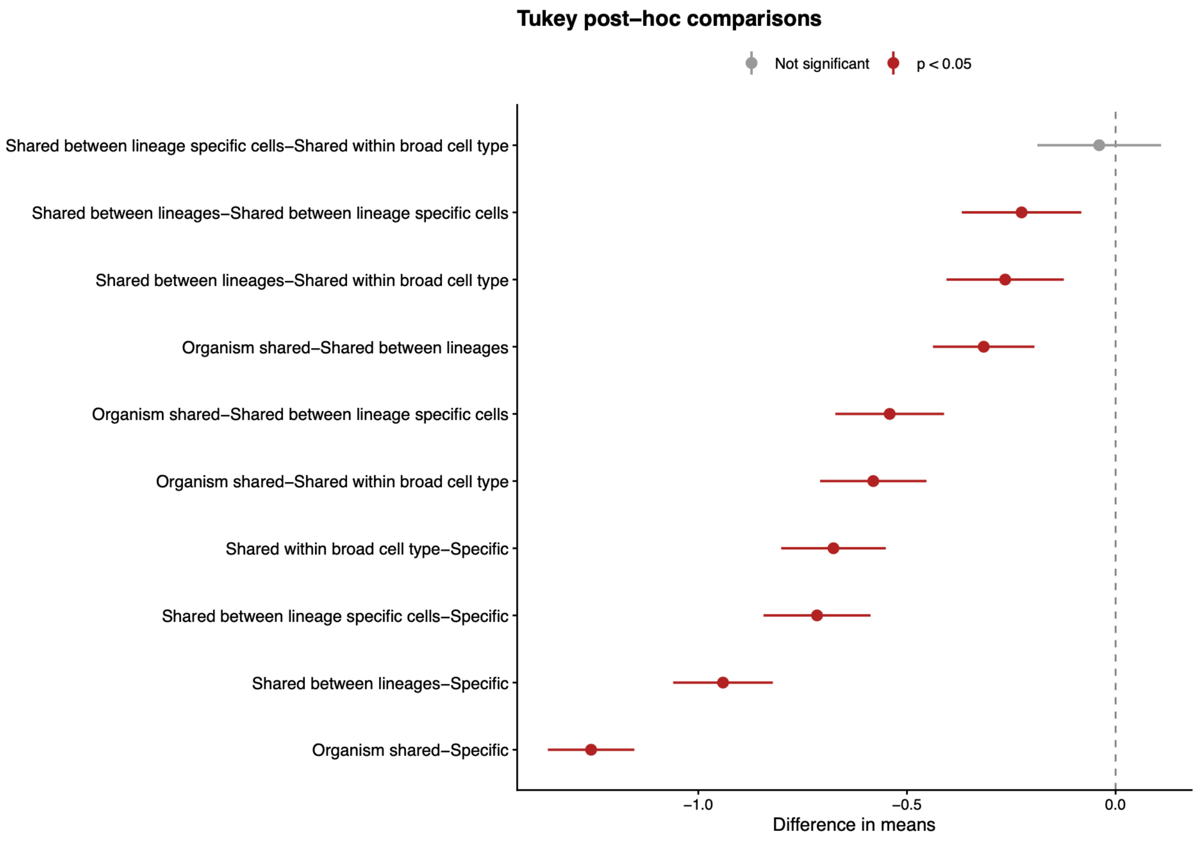

**Supplementary Figure S3 | TukeyHSD analysis result.**

The plot reports all pairwise differences in log10(distance from TSS) between different biological categories. Red indicates FDR <0.05. All categories were significantly different with the exception of the shared between lineage specific cells and shared between broad categories.

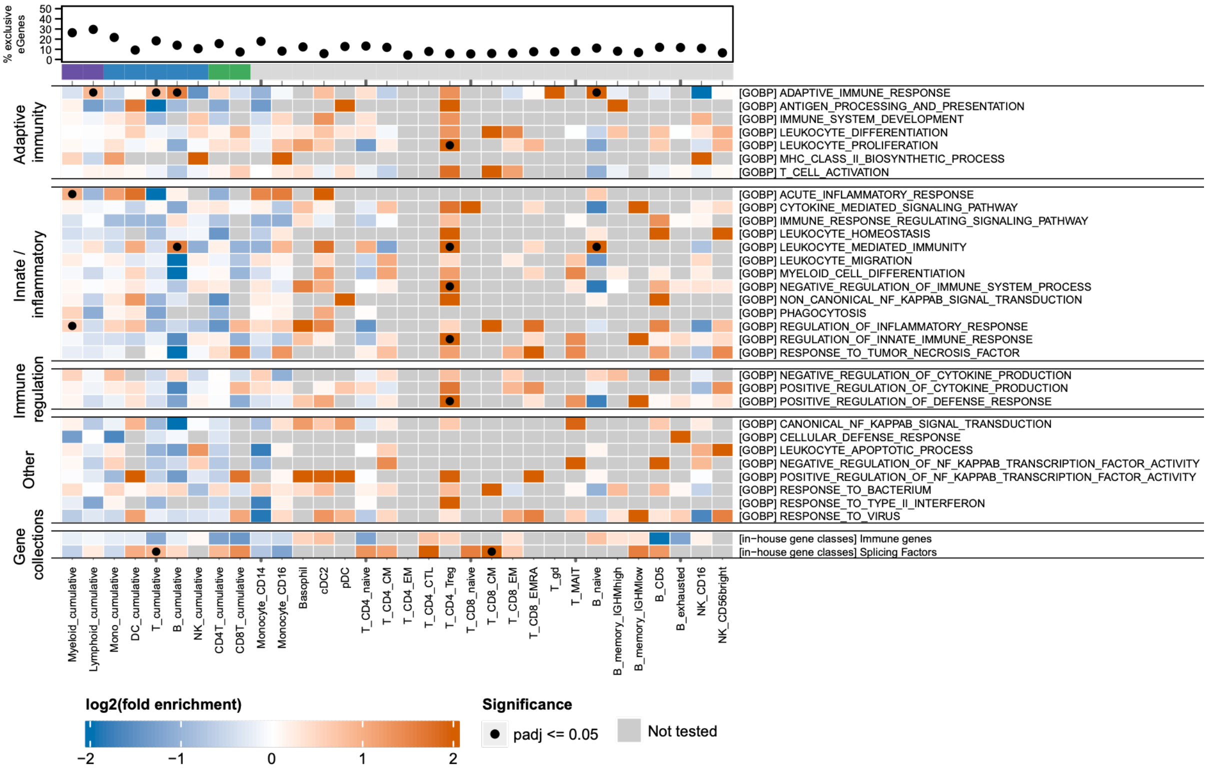

**Supplementary Figure S4 | Over-representation of immune gene-function annotations among cumulative-partition-exclusive eGenes.**

GOBP biological-process terms, grouped by theme (Adaptive immunity, Innate/inflammatory regulation, Immune regulation, Other), and in-house gene collections. Columns: 32 of the 33 cumulative pleiotropy partitions (**Methods and Supplementary Table S3**); Columns are ordered hierarchically in terms of cell types included and not clustered. Tile color: log2(fold enrichment); grey: not tested. Black dots: BH-adjusted p ≤ 0.05. Top dot plot: percentage of exclusive eGenes out of all eGenes detected per partition.

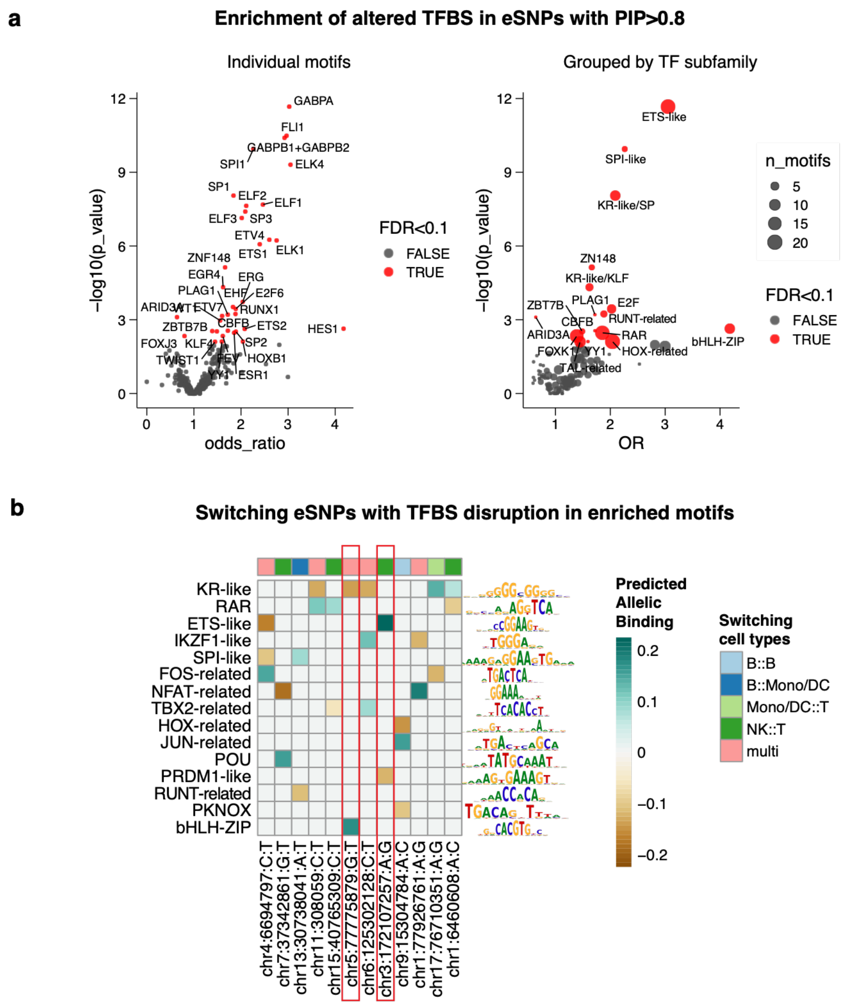

**Supplementary Figure S5 | Transcription factor-binding motif analysis.**

**(a)** Enrichment of predicted TF-binding motif alterations among eSNPs with PIP ≥ 0.8 relative to eSNPs with PIP < 0.8, highlighting the overrepresentation of canonical immune-lineage transcription factors. The left panel shows enrichment results for individual motifs, whereas the right panel shows only the most strongly enriched TF within each motif subfamily. Point size indicates the number of motifs represented. **(b)** Heatmap of predicted differential allelic binding at switching eSNPs (columns), which exhibit opposite eQTL effect directions across cell types. Values indicate the predicted change in binding for the effect allele, defined as the second allele in each pair (e.g., C in A:C). Only switching eSNPs with PIP ≥ 0.8 that overlapped snATAC-seq peaks and altered motif families observed at two or more variants are shown, representing the strongest candidate mechanisms underlying switching effects. TF motifs were collapsed by family, retaining the motif with the strongest predicted allelic effect.

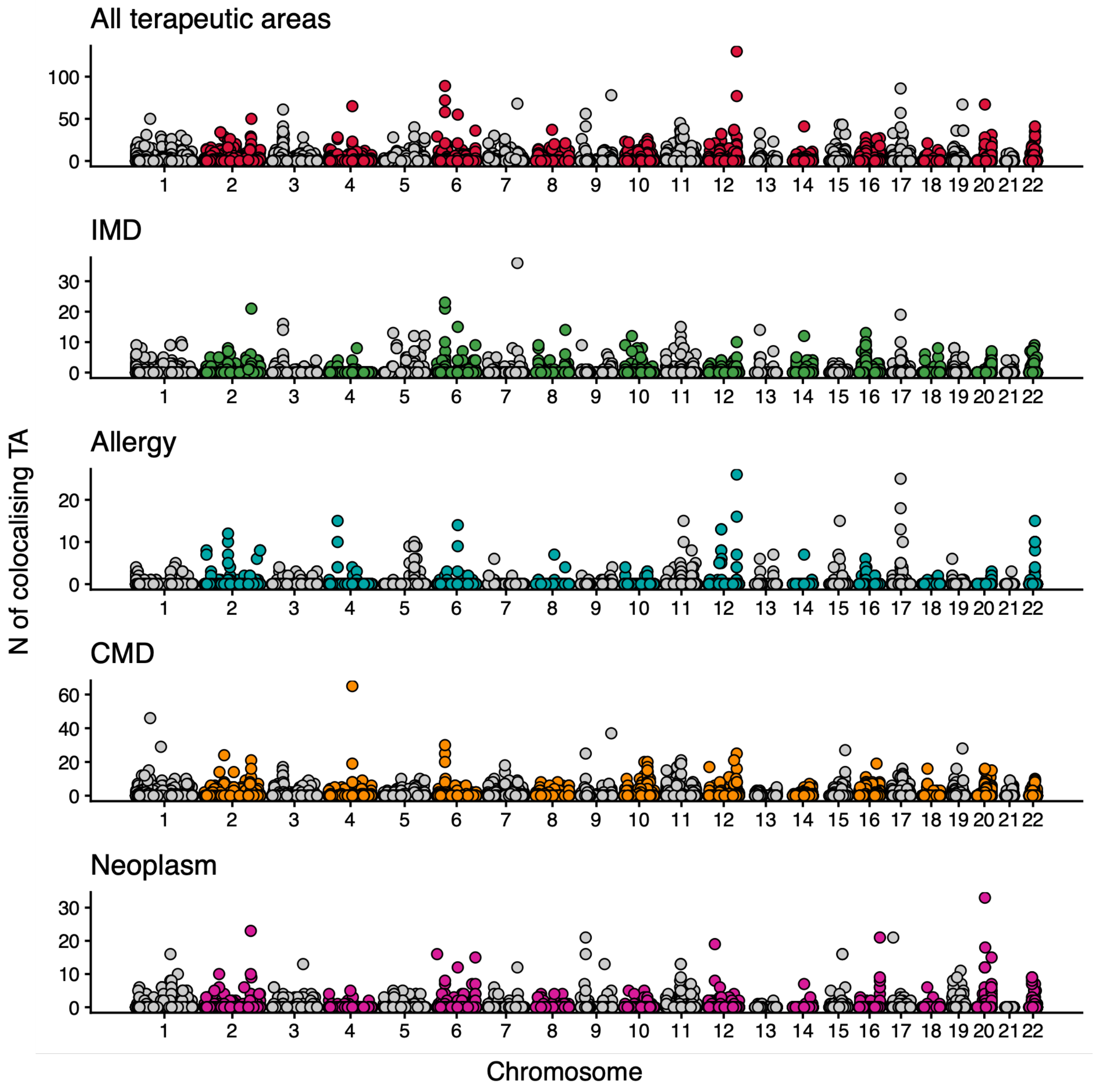

**Supplementary Figure S6 | Number of colocalising traits per eGene stratified by disease category.** Each point represents an eGene, positioned according to its genomic location. The y axis shows the number of traits with evidence of colocalisation with an eQTL for that gene, across all therapeutic areas (top panel) or within that disease category (remaining panels).

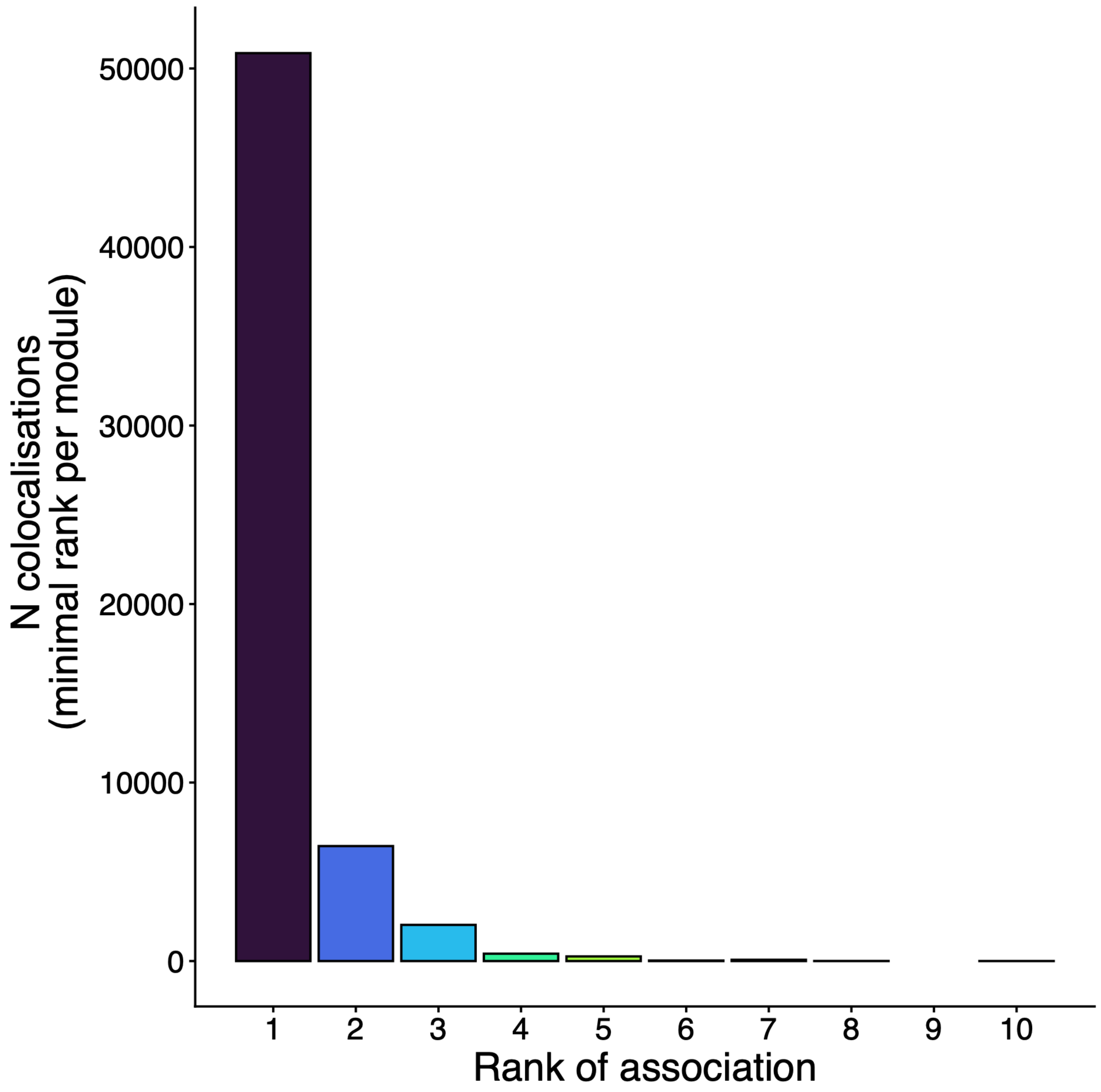

**Supplementary Figure S7 | Distribution of minimum signal rank of association per phenotype association.**

Bars show the number of colocalising module-phenotype pairs by the minimum rank of the colocalising association signals within each module. A minimum rank of 1 indicates colocalisation involving the primary signal, whereas a minimum rank ≥2 indicates colocalisation supported exclusively by secondary signals. Overall, 15.4% of colocalisations are explained uniquely by secondary signals.

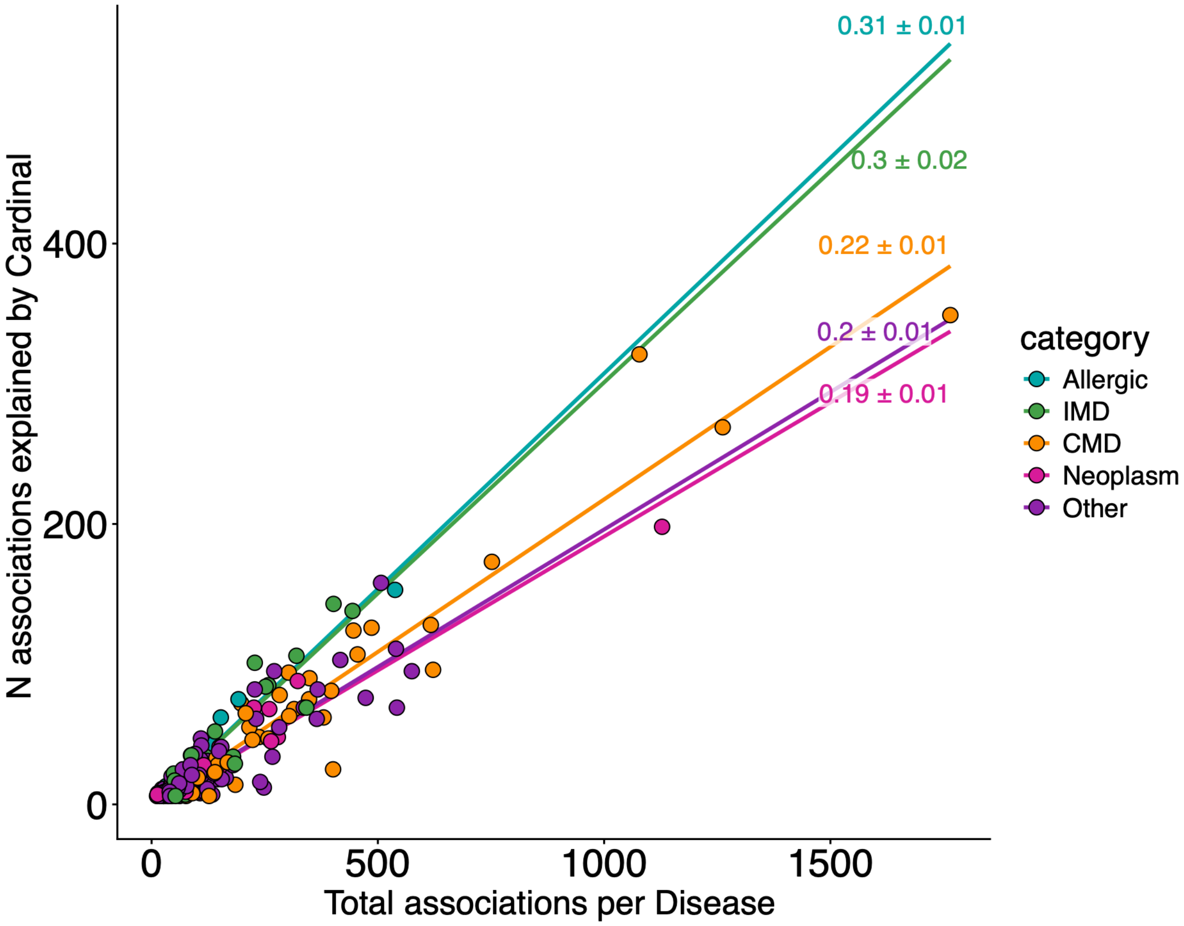

**Supplementary Figure S8 | Proportion of associations explained by sc-RNA eQTLs.**

Number of disease GWAS associations which colocalise with a sc-RNA eQTL (y axis), and total number of disease GWAS associations per disease (x axis). Colors reflect disease categories, the slope of the regression line represents the average proportion of loci explained by disease category.

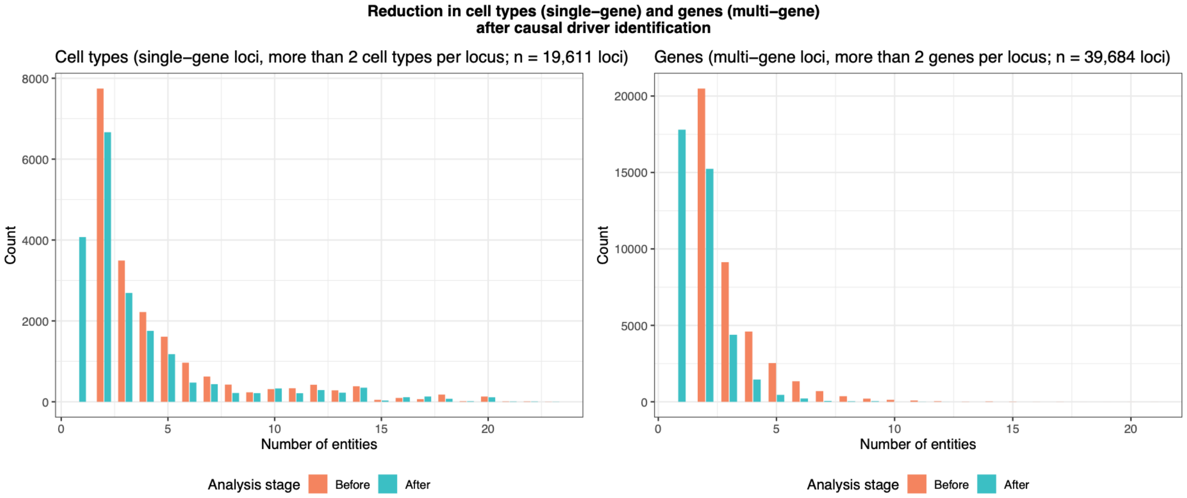

**Supplementary Figure S9 | Improvement in the number of cell types and genes given by prioritisation.**

Distribution of the number of cell types (left panel) and the number of genes (right) controlled by each eSNP before and after the prioritisation procedure. The results are shown only for pleiotropic eSNPs.

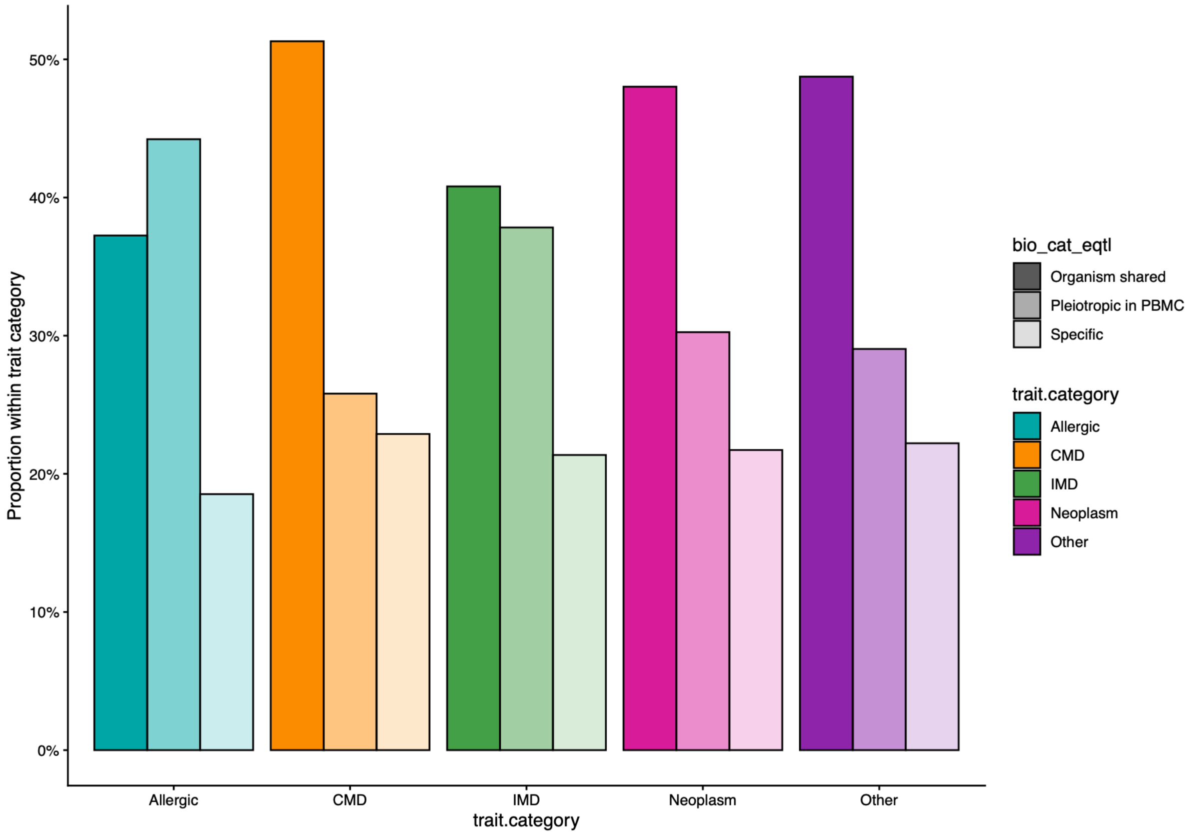

**Supplementary Figure S10 | Proportion of colocalisations for each biological category per disease class.**

Proportion of eSNPs in each biological category across disease classes. For ease of interpretation, the PBMC-restricted pleiotropic classes were collapsed into a single category. Immune-mediated diseases (IMDs) and allergic diseases show a substantially higher proportion of PBMC-restricted eSNPs, whereas in cardiometabolic diseases (CMDs), organism-wide shared eSNPs account for more than 50% of the total.

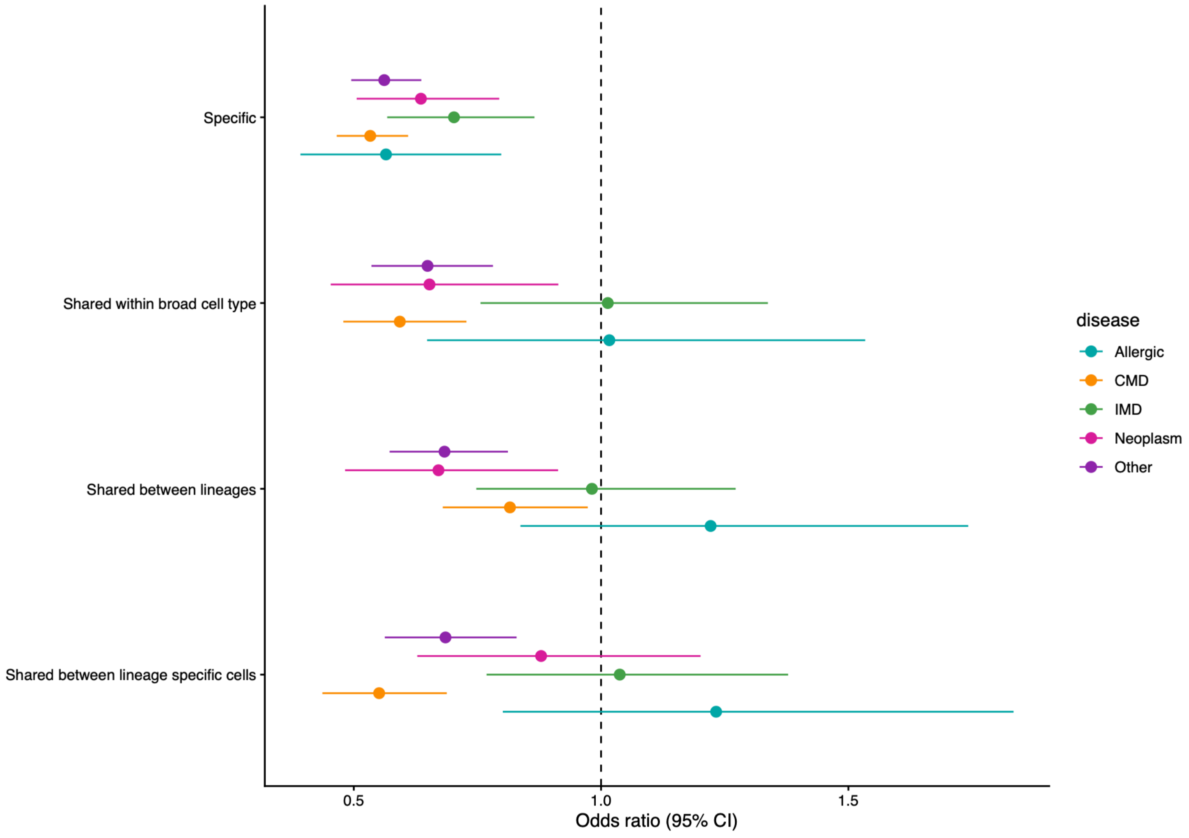

**Supplementary Figure S11 | Association between eSNP category and disease colocalisation by disease class.**

Odds ratios (ORs) for the probability that eSNPs in each biological category colocalise with a specific disease category, estimated using multivariable logistic regression and expressed relative to multi-tissue eSNPs. PBMC-restricted pleiotropic eSNPs are depleted for colocalisations with cardiometabolic diseases (CMDs), neoplasms and other diseases, but are as likely as multi-tissue eSNPs to colocalise with immune-mediated diseases (IMDs) and allergic diseases, despite being less frequent. Cell-type-specific eSNPs are less likely to colocalise with all disease categories.

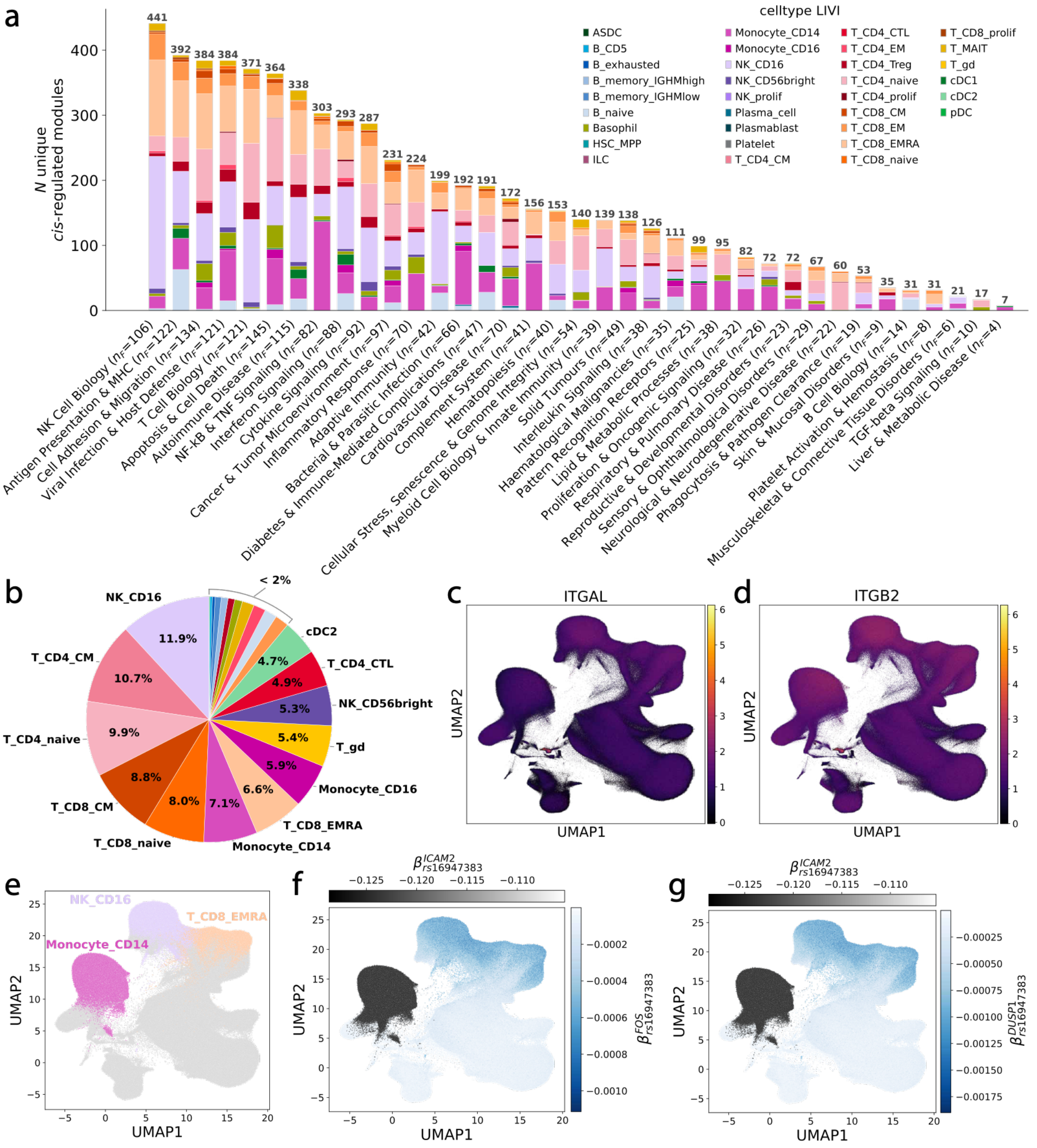

**Supplementary Figure S12 | Overview of LIVI programmes.**

**(a)** Stacked barplot showing the total number of unique cis-regulated modules (y-axis) that colocalise with LIVI gene programmes. LIVI programmes were annotated and then grouped into 37 categories (**Methods**; Table S17), which are shown on the x-axis. Within each category (bar) colors indicate the cell type in which the LIVI programmes belonging to the given category were primarily active (see **Methods** for details on LIVI programme to cell type assignment). The total number of programmes ($n_F$) comprising each category is shown in parentheses next to the category name. As shown, programmes capture diverse aspects of immune biology, spanning innate immune activation, antigen presentation, adaptive immunity, cytokine signalling and host defence, while also resolving disease-related biological pathways associated with viral infection, autoimmunity and cancer. **(b)** Piechart showing the proportion of colocalisation events between cis-regulated modules and LIVI programmes in each cell type. Only colocalisation events occurring in the same cell type are considered. Cell type labels are shown only for cell types contributing more than 2% of total colocalisation events. Colors same as in (a) . **(c-g)** UMAP on cell-state representation obtained by XX. Color denotes (c) the expression of *ITGAL*, (d) the expression of *ITGB2*, (e) cells annotated as $CD16^{+}$ NK, $CD14^{+}$ monocytes, or EMRA $CD8^{+}$ T , (f) the estimated effect of rs16947383 on the expression of *ICAM2* (cis-eGene; grey color) and on the expression of *FOS* (*trans*-eGene; blue color) in each cell, (g) the estimated effect of rs16947383 on the expression of *ICAM2* (cis-eGene; grey color) and on the expression of *DUSP1* (*trans*-eGene; blue color) in each cell.

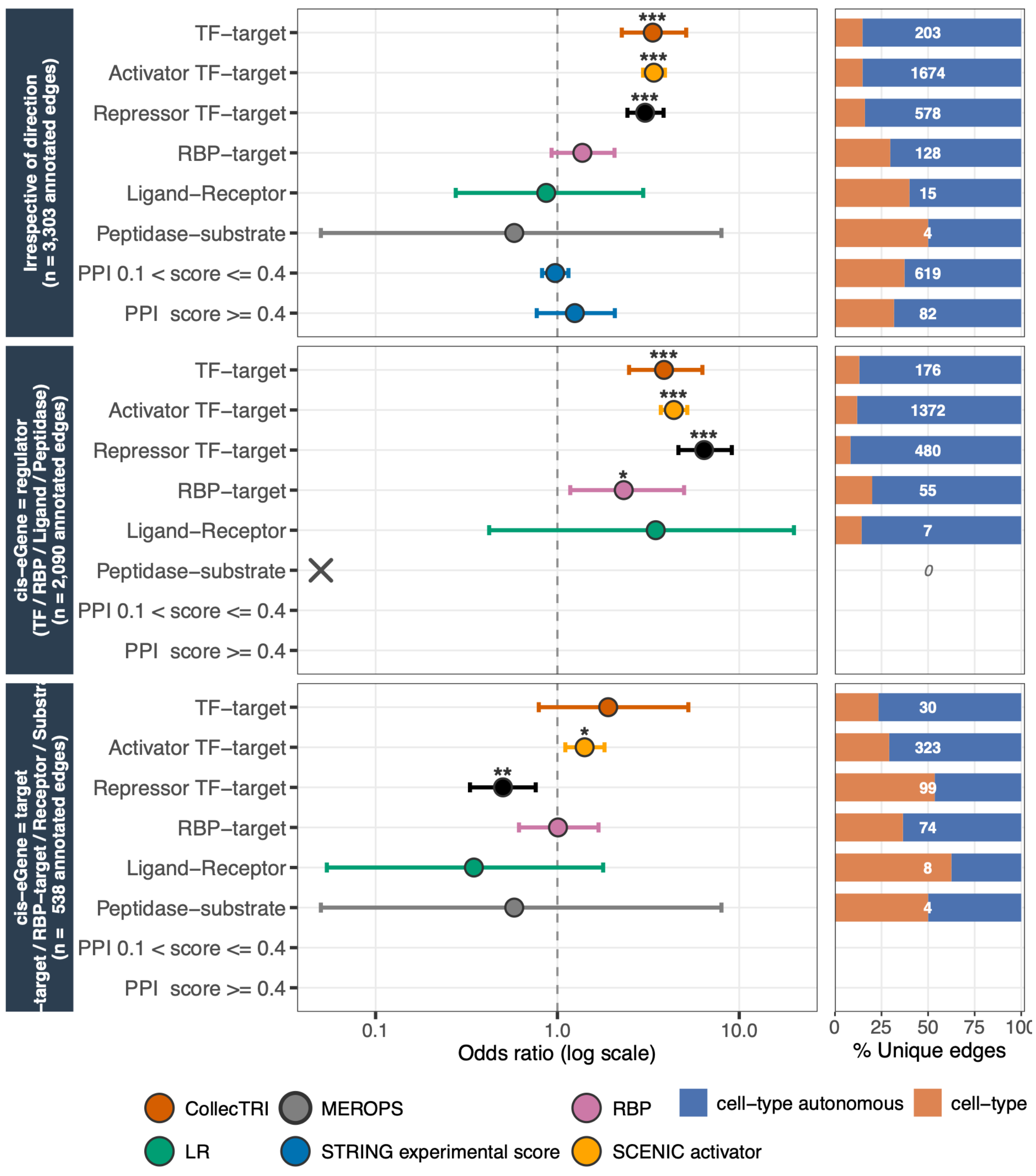

**Supplementary Figure S13 | Enrichment of gene regulatory annotations among cell-type autonomous versus non-autonomous eQTL edges.**

Given the directionality of some of the resources, we tested three complementary definitions of a "hit": (i) the gene pair corresponding to an edge is present in the resource, irrespective of orientation; (ii) the cis-eQTL source gene occupies the regulatory role (TF, RBP, ligand or peptidase); or (iii) the cis-eQTL source gene occupies the target role (TF target, RBP target, receptor or substrate). Points and error bars show odds ratios and 95% confidence intervals from Fisher's exact tests. Asterisks denote Benjamini–Hochberg-corrected significance (*$q < 0.05$, **$q < 0.01$, ***$q < 0.001$); crosses (×) indicate degenerate 2 × 2 contingency tables, with no annotated edges in either group; and blank rows indicate tests that were not applicable to the corresponding resource (STRING for tests ii and iii). Bars on the right show, for each resource and test, the proportions of annotated edges that were cell-type autonomous or non-autonomous, with the number of annotated edges indicated within each bar.

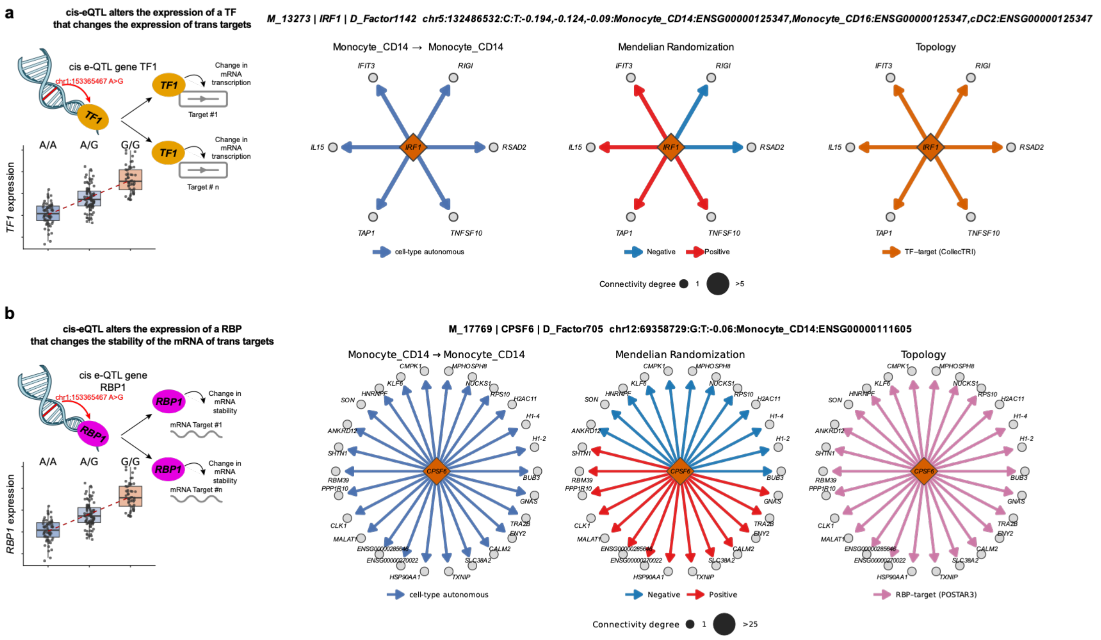

**Supplementary Figure S14 | Topological annotation and mechanistic hypotheses for cis/trans modules.**

In each row, the leftmost panel illustrates a mechanistic hypothesis for how a cis-eQTL may alter the expression of its trans target(s). The subsequent three panels show (left) the cell-type-autonomy classification of each edge (left), (middle) the direction of the Mendelian-randomisation effect (Methods section Definition of disease specific *cis-trans* graphs), and (right) the annotation resource and the topological relations. (a) A cis-eQTL acting on a transcription factor (TF) propagates to that TF's target genes *in trans*. We illustrate this with genetic signal M_13273, which alters expression of the TF *IRF1* across myeloid cell types (Monocyte_CD14, Monocyte_CD16 and cDC2; lead variant chr5:132486532 C>T; cis effect size −0.194 in Monocyte_CD14, −0.124 in Monocyte_CD16 and −0.09 in cDC2). This cis-eQTL has trans effects on a series of genes included in the LIVI programme D_Factor142. *IRF1* is the principal driver of the interferon-stimulated gene and antigen-presentation programme in human monocytes and macrophages, where it binds target loci and increases their chromatin accessibility [1]. Consistent with this role, the module recovers CollecTRI-annotated *IRF1* targets *in trans*, notably the antigen-processing transporter *TAP1*, a direct *IRF1* target with demonstrated *IRF-1*-dependent regulation [2]. (b) A cis-eQTL acting on an RNA Binding protein (RBP) propagates to that RBP target genes *in trans*. We illustrate this with the genetic signal M_17769 which alters the expression of the RBP *CPSF6* in Monocyte_CD14 cells (chr12:69358729 G>T; cis effect size -0.06). This cis-eQTL has trans effects on a series of genes from the LIVI programme D_Factor705. *CPSF6* is a core 3′-end / alternative-polyadenylation (APA) factor that controls macrophage antiviral responses [3].

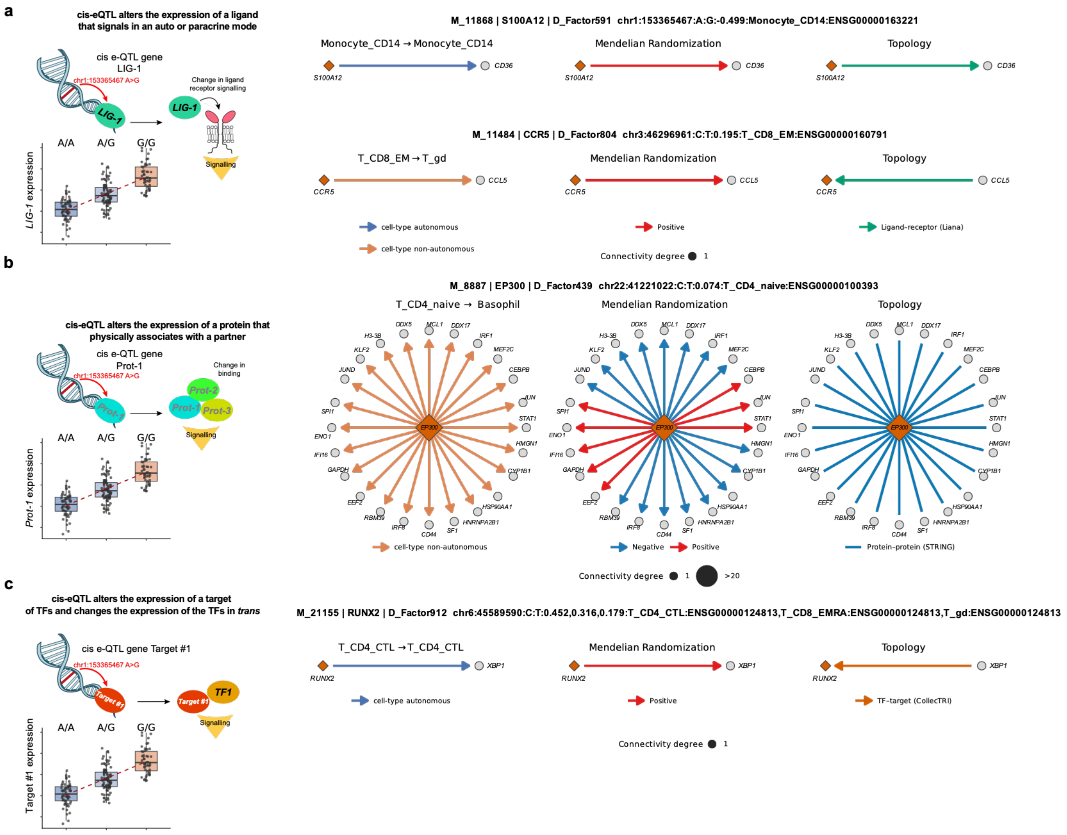

**Supplementary Figure S15 | Topological annotation and mechanistic hypotheses for cis/trans modules.**
**(a Top panel)** A cis-eQTL acting on a ligand propagates to the expression of the receptor gene *in trans*. We illustrate this with genetic signal M_11868, which alters expression of the ligand *S100A12* in Monocyte_CD14 cells (chr1:153365467 A>G, cis effect size -0.499). This cis-eQTL has trans effects on a series of genes from the LIVI programme D_Factor591. The class-B scavenger receptor *CD36* binds the monocyte alarmin *S100A12* with high affinity [4], an interaction proposed to contribute to atherosclerosis [5]. **(a lower panel)** A cis-eQTL acting on a receptor propagates to the expression of the ligand gene *in trans*. We illustrate this with genetic signal M_11484, which alters expression of the receptor *CCR5* in T_CD8_EM cells (chr3:46296961 C>T, cis effect size 0.195) and affects the expression of its ligand *CCL5* in TgD cells (belonging to the LIVI programme D_Factor804) Functional *CCR5* on effector-memory CD8 T cells and *CCR5–CCL5* control of T-cell responses are well established [6]. **(b)** A cis-eQTL acting on a protein propagates to the expression of its physical interaction partners *in trans*. We illustrate this with genetic signal M_8887, which alters expression of the transcriptional co-activator *EP300* (p300) in T_CD4_naive cells (chr22:41221022 C>T; cis effect size +0.074). This cis-eQTL has trans effects on a series of genes from the LIVI programme D_Factor8887. p300 is a promiscuous, limiting co-activator that physically docks onto sequence-specific transcription factors; consistent with this, the STRING-annotated partners recovered *in trans* include documented p300 interactors such as C/EBPβ (*CEBPB*) [7] and *STAT1* [8]. **(c)** A cis-eQTL acting on a gene that is itself the *target* of a transcription factor can feed back to alter that regulator's expression *in trans*. Genetic signal M_21155 alters expression of *RUNX2* in T-cell subsets (chr6:45589590 C>T; cis effect size +0.452 in T_CD4_CTL, +0.316 in T_CD8_EMRA, +0.179 in T_gd). This cis-eQTL has trans effects on a series of genes from the LIVI programme D_Factor912. *RUNX2* is annotated as a target of the TF *XBP1* (CollecTRI). The module recovers a trans edge from *RUNX2* onto *XBP1*, consistent with feedback from a target gene onto its upstream regulator. *RUNX2* itself has an established role in CD8$^{+}$ memory T-cell persistence [9]. The recovered edge onto XBP1 is consistent with feedback from a target gene onto its upstream regulator. In all panels, the cartoon (left) depicts the hypothesised mechanism; the panel immediately to its right shows the cell-type-autonomy classification of each edge, the middle panel the direction of the Mendelian-randomisation effect, and the rightmost panel the annotation resource.

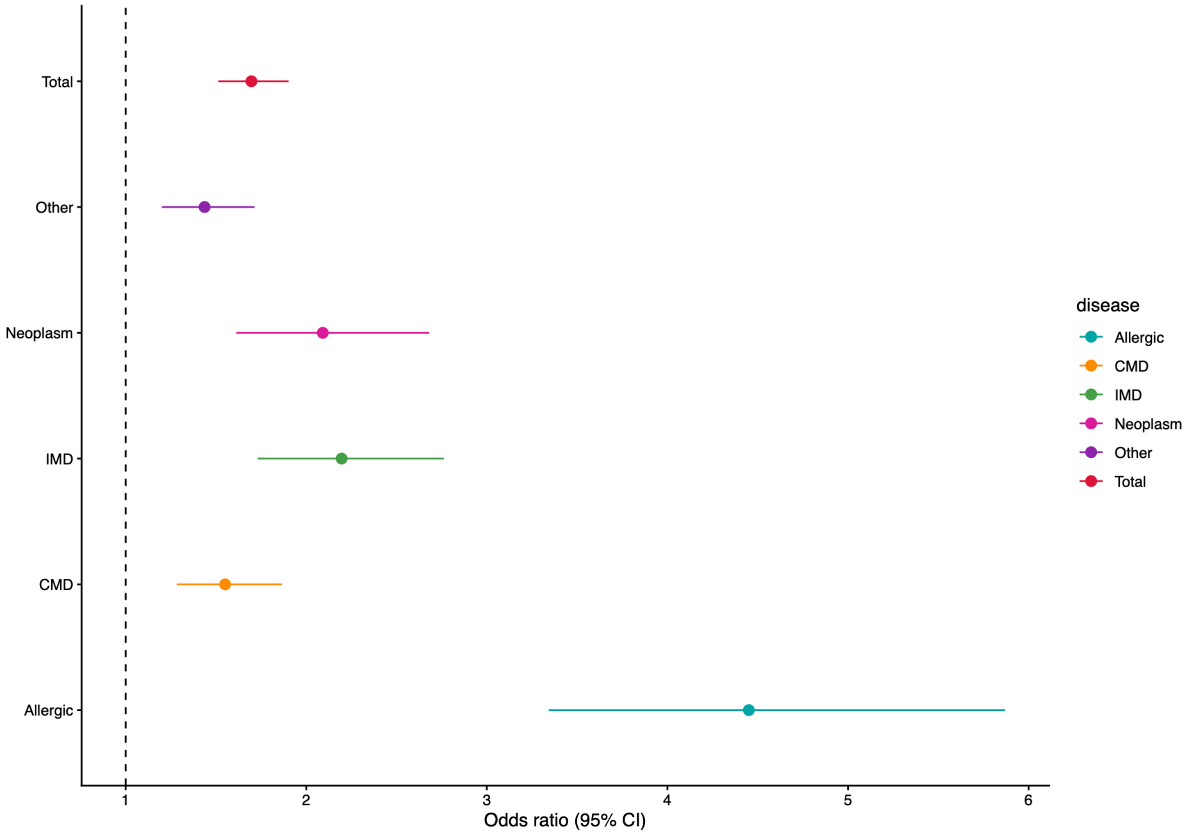

**Supplementary Figure S16 | Enrichment in colocalisation probabilities for factor SNPs (fSNPs) and different disease classes.**

Results from multivariable logistic regression assessing the probability that fSNPs colocalise with specific disease classes. Although fSNPs are enriched for colocalisation across all disease classes, the enrichment is particularly strong for allergic diseases, for which the odds ratio reaches 4.5.

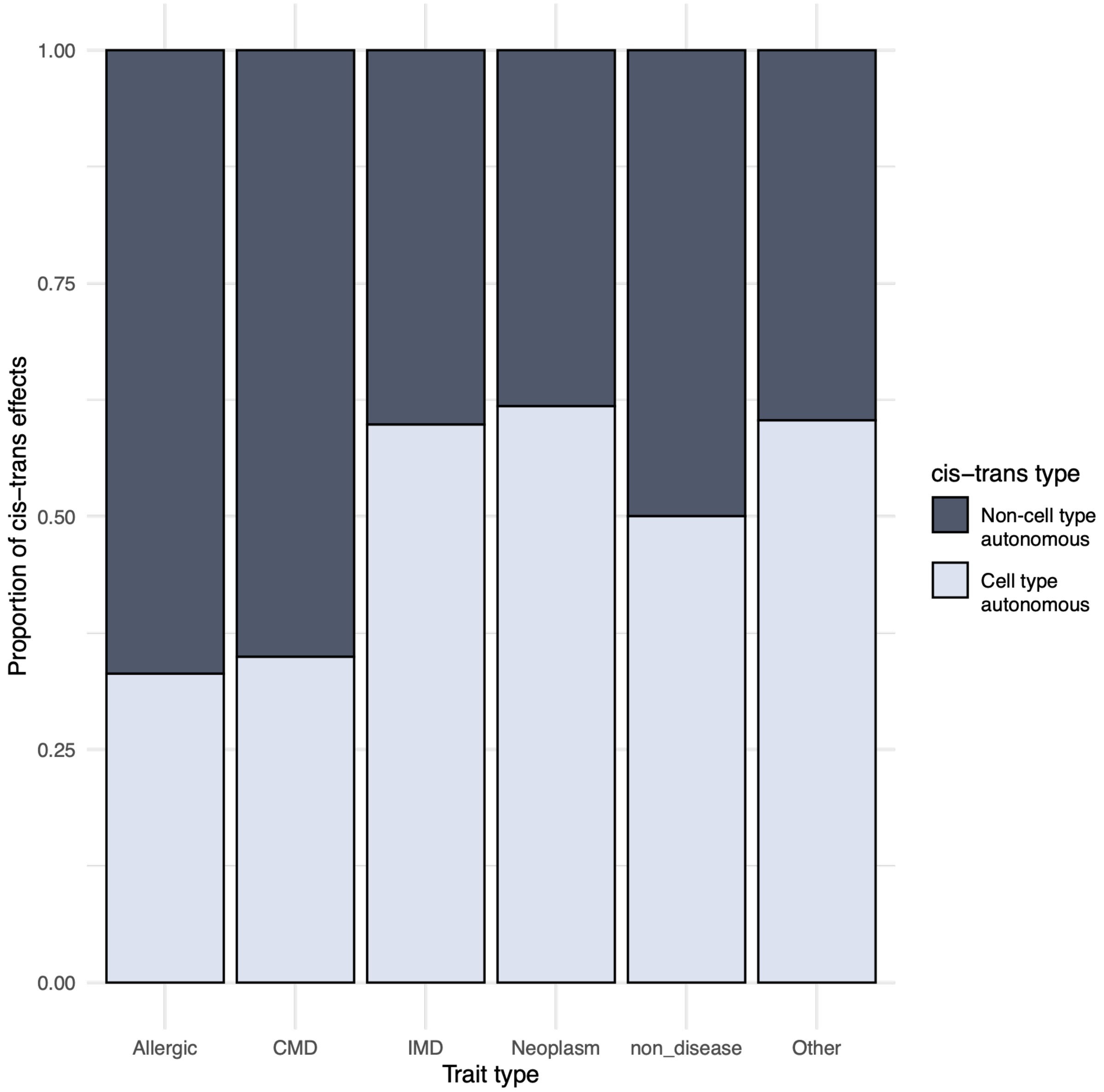

**Supplementary Figure S17 | Proportions of cell autonomous and non-cell autonomous edges in disease-specific networks.**

Allergic diseases and cardiometabolic diseases (CMDs) show a higher proportion of non-cell autonomous cis–trans regulatory edges.

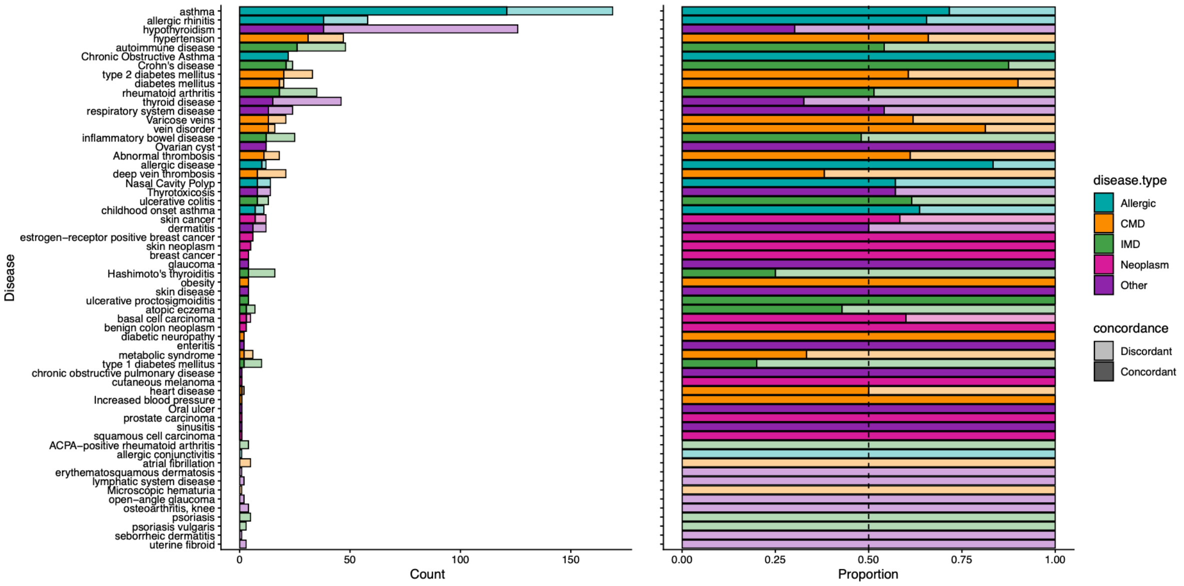

**Supplementary Figure S18 | Number and direction of graph edges across diseases.**

The left panel shows the number of concordant (darker shade) and discordant (lighter shade) edges for each disease, with colours indicating disease categories. The right panel shows the corresponding proportions of concordant and discordant edges.

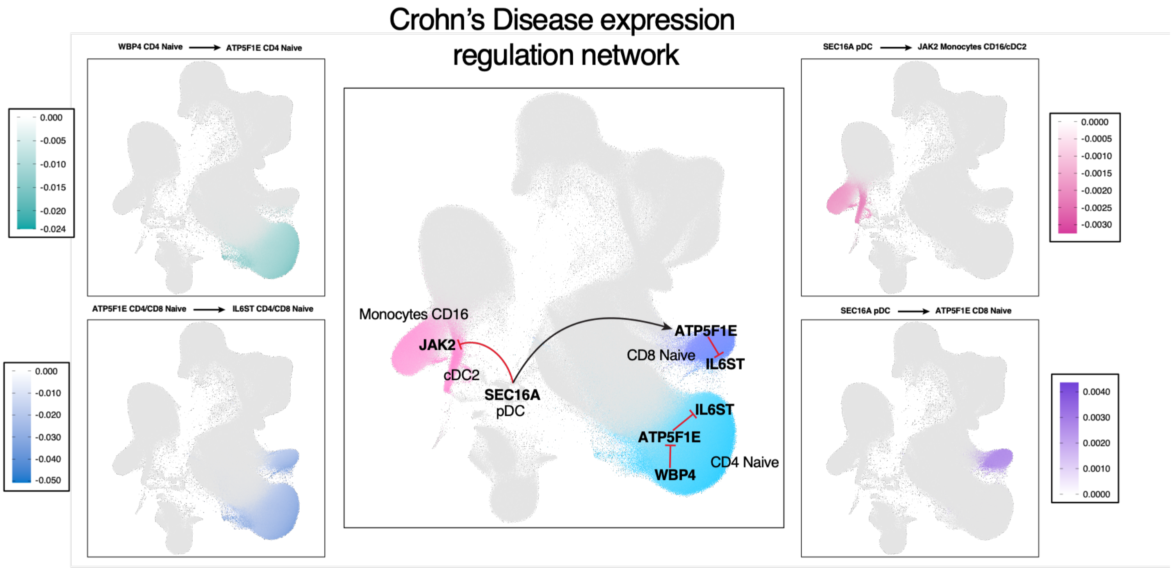

**Supplementary Figure S19 | Inferred regulatory relationships linking Crohn's disease risk genes across immune cell types.**

The central panel shows a putative regulatory network linking *SEC16A*, *JAK2*, *WBP4*, *ATP5F1E* and *IL6ST*. Increased *SEC16A* expression in pDCs is protective against Crohn's disease by reducing *JAK2* expression and increasing *ATP5F1E* expression, which in turn reduces *IL6ST* expression. Conversely, increased *WBP4* expression increases disease risk by inhibiting *ATP5F1E* expression, thereby increasing *IL6ST* expression. Arrows and blunt-ended lines indicate positive and negative inferred regulatory effects, respectively. Surrounding UMAP panels show single-cell estimates of the effects corresponding to individual *cis–trans* relationships, with colour scales indicating effect size.

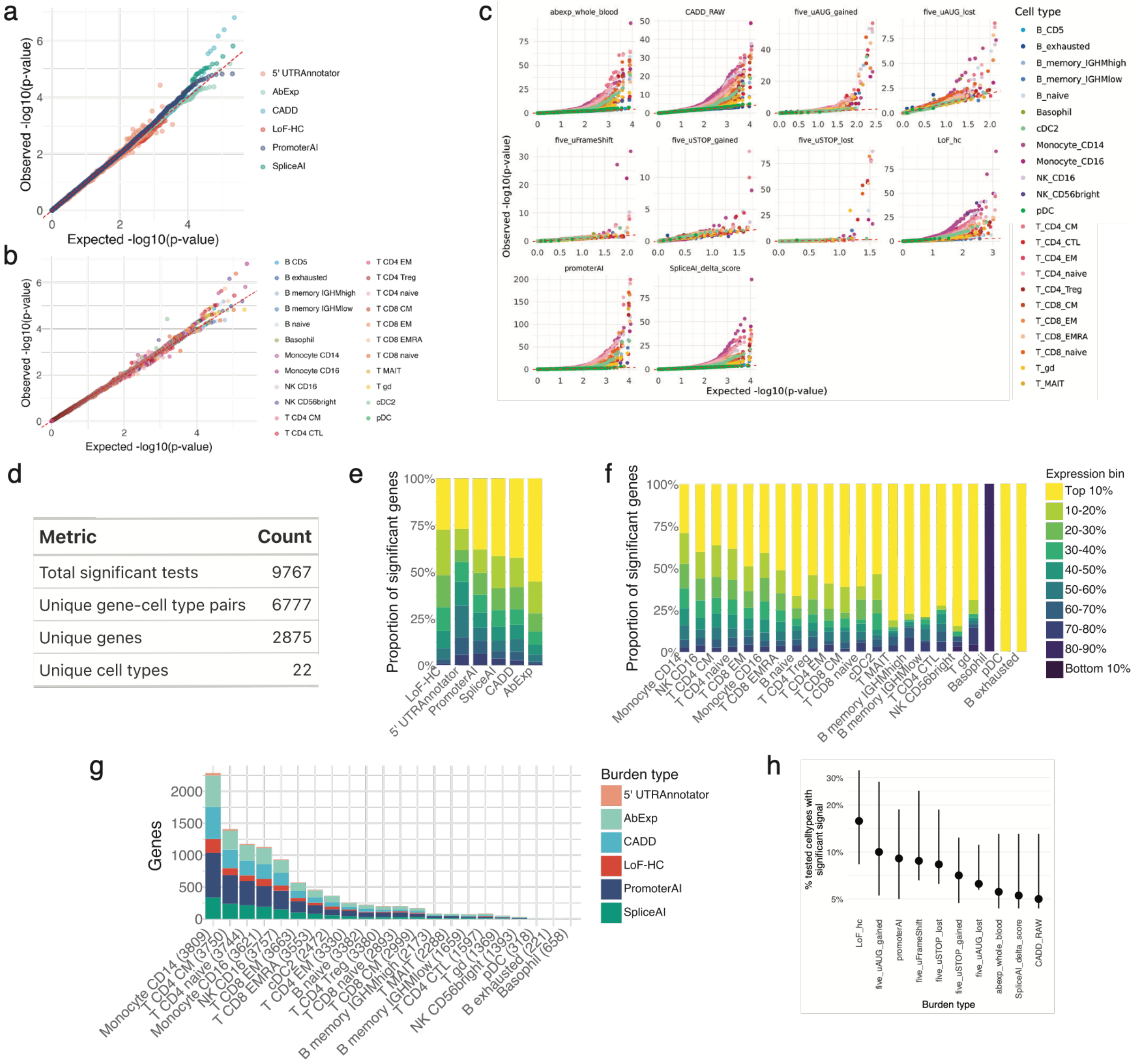

**Supplementary Figure S20 | Summary of RVAT discovery.**

**(a-c)** QQ plots for the rare variant test across considered burden types **(a)**, cell types **(b)** and burden type - cell type combinations **(c)**. **(d)** Summary counts of significant eGene signals discovered by rare variant analysis. e-f, The barplots report the fraction of significant eGenes discovered across gene expression quantile bins, stratified by burden type (d) or cell type (e). g, Number of significant eGenes discovered by cell type, stratified by burden type. h, The plot reports for each burden type the distribution (median, 1st/3rd quartile range) of the fraction of tested cell types where a given eGene was found significant in the rare variant analysis.

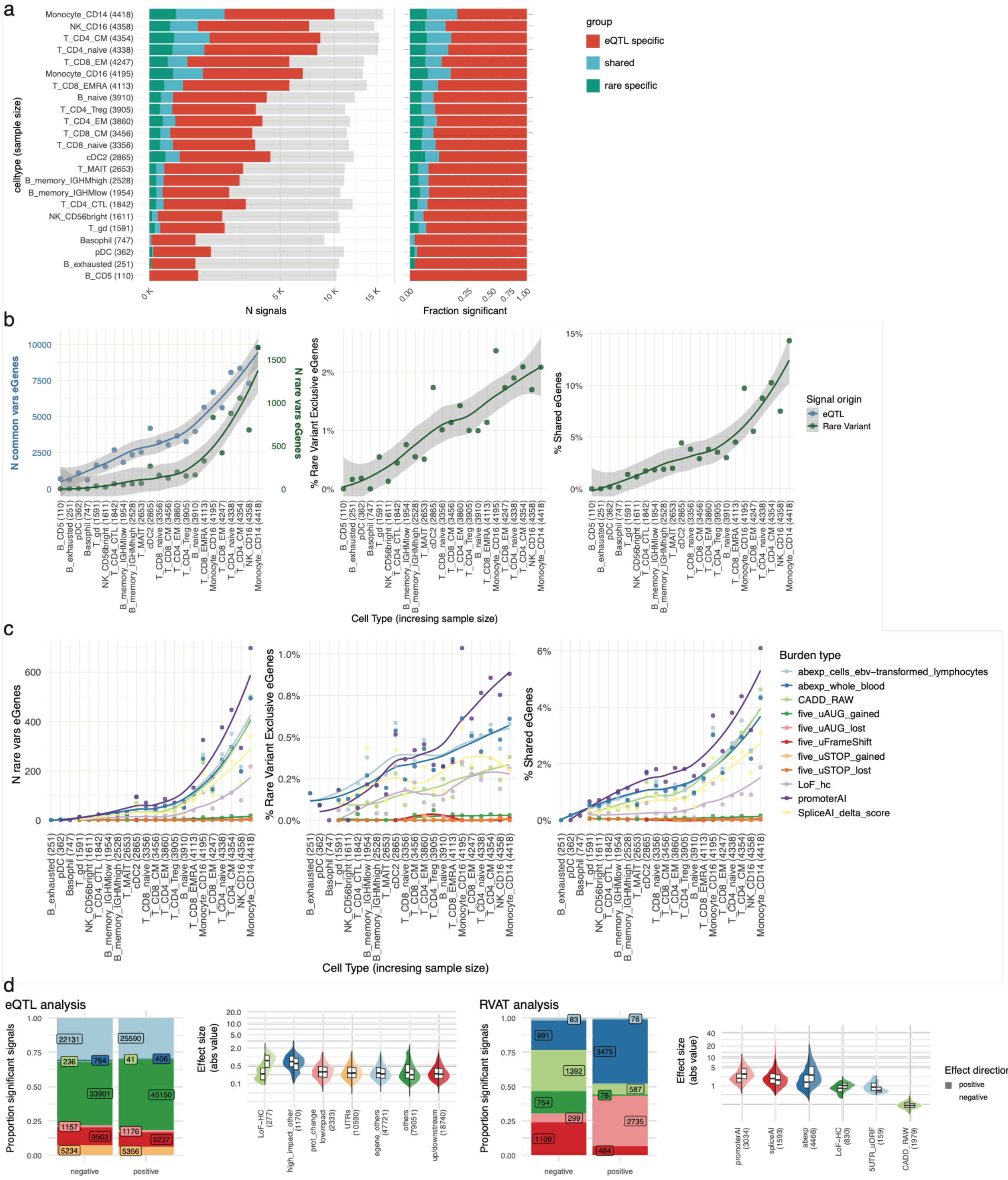

**Supplementary Figure S21 | eQTL-RVAT comparison.**

**a,** Overall comparison of detected eGenes when comparing results across cell types between standard eQTL analysis and the rare variant test results. The grey bar represents the total amount of tested genes. **b,** Effect of celltype sample size on eGene discovery for eQTL and rare variant analysis. The central and right panels represent the fraction of eGenes discovered only by rare variant analysis and the fraction of shared eGenes, respectively. **c,** Effect of celltype sample size on eGene discovery for the single burden types considered in the rare variant analysis. **d,** We report the fraction of significant signals having a negative / positive direction of effect for the eQTL leading variant stratified by variant consequence and for rare variant analysis stratified by burden type. The violin plots report the distribution of absolute effect sizes stratified in the same way. The eQTL variants impact are classified as: LoF-HC, high confidence loss of function; high_impact_other, other variant with potential classified as deleterious by spliceAI, promoterAI, or CADD scores; prot_change_lowimpact, missense variants with no score support, low confidence LoF, other protein changing labels; egene_others: other type of variants in the eGene like intronic or synonymous; other: all other variants outside the eGene.

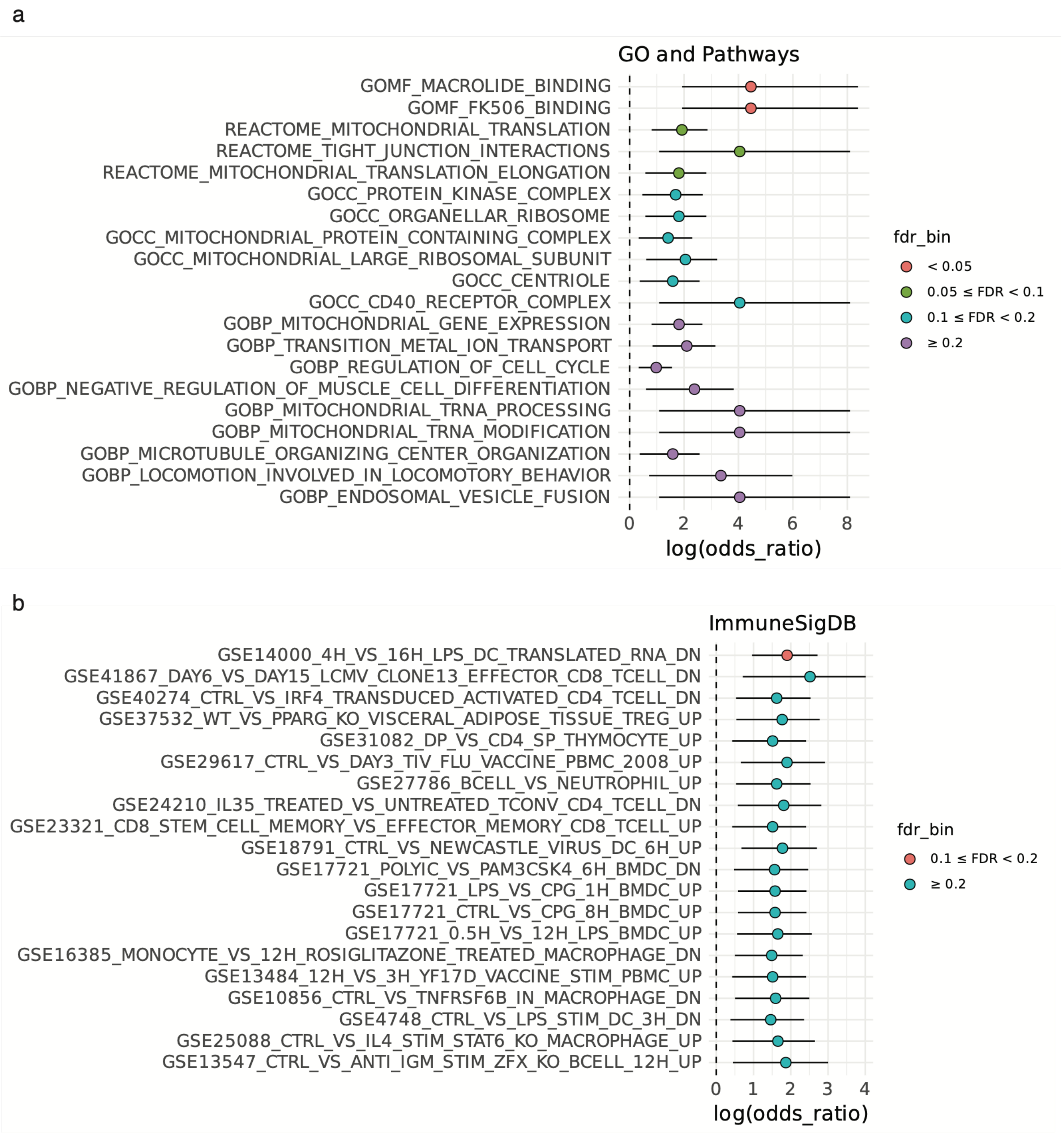

**Supplementary Figure S22 | RVAT exclusive genes enrichments.**

Over-representation analysis of eGenes detected only in rare variant analysis. The plot shows the top 20 genes ranked by FDR from enrichment results on Gene Ontology categories and REACTOME pathways (**a**) or ImmuneSigDB signatures (**b**).

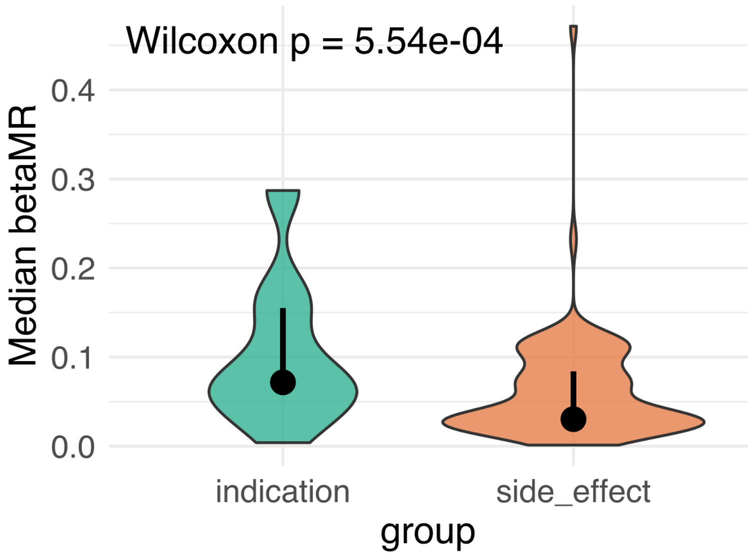

**Supplementary Figure S24 | Effect of drug target on indication vs on predicted side effects.**
Mendelian-randomisation effect estimates (betaMR) for approved indications and reported adverse effects. Indications show stronger genetic effects than adverse-effect associations (two-sided Wilcoxon test).
