## Supplementary Note FLANDERS methods and benchmarking for "Hierarchical genetic regulatory architecture of the human immune system informs complex disease mechanisms"

### Supplementary Notes

#### FLANDERS (Fine-mapping coLocalisation AND plEiotRopy Solver) workflow

The FLANDERS workflow aims at identifying groups of any kind of traits which are likely driven by the same underlying causal SNP, here called cis-regulated modules, while improving fine-mapping of the genomic region and enabling comparison of SNP effect sizes and directions.

The workflow is divided in several steps:

1) Harmonisation of GWAS summary statistics
2) Identification of association regions with 'Locus breaker'
3) Fine-mapping of each locus
4) Identification of 'coloc-sufficient' credible sets (CS)
5) Storage of coloc-sufficient CS into cs-AnnData object
6) Identification of overlapping CS (sharing at least one SNP)
7) Pairwise colocalisation with iColoc
8) Representation of pairwise positive colocalisations (PPH4>=0.75) as a graph and identification of groups of connected nodes
9) Resolving non-fully connected groups using SuSiE-HyPrColoc
10) Performing multi-trait joint fine-mapping and identifying the 'most likely causal SNP'
11) Storage of cis-regulated module CS in cs-annData for future colocalisations.

#### GWAS summary statistics harmonisation

GWAS summary statistics were harmonised by changing the genetic variants id to chromosome:GRCh38 position:A1:A2, where A1 is the first allele in alphabetical order while A2 is the second one. All association statistics are referred to A1 as the effect allele. This allows to eliminate any ambiguity in coding downstream as the alphabetical order can always be reconstructed without further information.

#### Locus identification with Locusbreaker

To identify the genomic regions significantly associated with a specific trait, we implemented an algorithm that is able to define genomic region boundaries based on the shape of the association peak. Locusbreaker has been previously described elsewhere[1]. Briefly, we define the boundaries of each significant genomic region by identifying all the SNPs with a p-value lower than a specific threshold, which is more liberal than the desired significance p-value. For cell type-eGene pairs having at least no SNP showing a nominal p-value < $1x10^{-6}$, the limit threshold was set to one order of magnitude higher than the minimum p-value within the window. Otherwise, a fixed threshold of $1x10^{-5}$ was applied. We then calculate the distance between each consecutive SNP below this threshold in the same chromosome; if two SNPs are further than a predefined distance, then they are considered as belonging to two different genomic regions. We used a default value of 250kb, as it is the average distance at which LD decays [2]. We then retain all regions where at least one SNP has a nominal p-value below the desired significance p-value threshold, which for eQTLs was set to twice the minimum p-value observed within the window. To capture the whole locus, the boundaries are then enlarged to capture the 'tails' of the association peak. To avoid overlapping loci this distance should be less than half of the distance that defines the minimum distance between two adjacent loci as default we have used 100kb. This algorithm allows to define the boundaries of each associated genomic region based on the local LD characteristics rather than a predefined distance between top SNPs, allowing for capturing very large loci while limiting the boundaries of very short association peaks. **Fig. S25**. shows the distribution of loci size after applying locus breaker to the Cardinal eQTLs. We found that the median locus size was 400kb, ranging between 138kb and 2.2 Mb. In comparison with the widely used approach of defining the locus as +/- 1MB from the lead SNP, we find that ~89% of the loci are smaller while 11% are larger, if we consider 100kb (10%) tolerance around the 1MB window only 4% of loci would fall within it.

#### Fine-mapping

Following locus identification, fine-mapping was performed within each locus using the Sum of Single Effects (SuSiE) model, as implemented in the susieR package [3] , with in-sample LD. SuSiE was run allowing up to 10 causal independent signals per locus (L=10), a target credible set coverage of 0.99 and a maximum of 400 iterations of the iterative Bayesian stepwise selection (IBSS) algorithm. When no credible set was identified at the initial requested coverage, fine-mapping was repeated by iteratively relaxing the coverage in decrements of 0.01, down to a minimum coverage of 0.7. If SuSiE still failed to identify any credible set under these relaxed criteria, or if the IBSS algorithm did not converge within the set number of iterations (400 by default) or yielded an implausibly large prior variance estimate, we instead applied Wakefield's approximate Bayes factor (ABF) method [4]. In these cases, we assumed a single causal variant within the locus (L=1). To eliminate independent signals which are statistically unreliable, we applied basic quality control on the 99% credible sets defined by SuSiE based on **(i)** minimum credible set Bayes factor threshold (*cs_bf_thr = 3*), **(ii)** statistical significance of the top variant in each credible set, assessed using a P-value threshold (*signal_pval_threshold = 1*), and **(iii)** linkage disequilibrium–based purity metrics, requiring both the mean and minimum pairwise $r^2$ within each credible set to exceed predefined thresholds (*purity_mean_r2_threshold = 0*; *purity_min_r2_threshold = 0*). Credible sets failing any of these criteria were excluded from downstream analyses.

##### Coloc sufficient credible sets

Fine-mapping aims at identifying the SNPs most likely to drive an observed association by estimating their posterior probability of causality from Bayesian evidence. These probabilities are typically summarised in credible sets containing, for example, 95% or 99% of the total posterior probability [5,6]. SNPs outside these sets collectively account for only a small fraction of the remaining probability and therefore generally have similarly low lABF values. However, a conventional 99% credible set may not retain all the information needed for downstream colocalisation. At very large sample sizes, posterior probabilities can become highly concentrated, such that a single SNP carries more than 99% of the total probability even when several neighbouring SNPs retain substantial lABF support. Restricting storage to the 99% credible set in these cases would therefore discard potentially informative variants. To address this, FLANDERS identifies variants with high lABF values and defines a "coloc-sufficient" credible set as the larger of the conventional 99% credible set and the set of variants selected on the basis of high lABF support. This preserves the information required for fine-mapping while retaining sufficient resolution for downstream colocalisation. To efficiently store all the information coming from the fine-mapping of a dataset we extended the AnnData [7] to represent fine-mapping results by organising credible sets (CSs) as columns and SNPs as rows. The primary data matrix (X) stores log approximate Bayes factors (lABFs, natural log scale) in a sparse format, where each non-zero element represents an SNP being included in a particular colocalisation-sufficient credible set. Metadata for each CS - including study ID, phenotype, genomic locus, and quality control metrics - is stored in the obs field (column annotations), while SNP-level information such as chromosome, position, and alleles is stored in the var field (row annotations). Additional layers store effect sizes (beta) and standard errors (se) derived from the SuSiE joint model, which are conceptually similar to those estimated by COJO.

##### Retrieving joint betas and standard error from SuSiE fine-mapping

To obtain conditional effect sizes ($beta$) and standard errors ($se$) for each $SNP_j$ within credible set $L$, we extracted the estimation of the effect size and its standard error from the posterior mean ($\mu_{L,j}$) and posterior second moment ($\mu 2_{L,j}$) from the SuSiE model output. For a given single effect $L$ (i.e. credible set), the effect size $beta$ and standard error $se$ were computed as:

$$beta_j = \frac{\mu_{L,j}}{s_j};\ se_j = \frac{\sqrt{\mu 2_{L,j} - \mu_{L,j}^2}}{s_j}$$

where $s_j$ denotes the column-wise scaling factor applied to genotype matrix X during standardisation (typically the standard deviation of $SNP_j$). These quantities correspond to the posterior mean and variance of regression coefficients under the SuSiE model. This formulation is mathematically equivalent to estimates obtained from a multiple (joint) linear regression model, similar to the approach used in GCTA-COJO [8].

##### Colocalisation analysis with iColoc

Colocalisation assesses whether association signals for two traits share an underlying causal variant. SuSiE-coloc first resolves signals within each trait through fine-mapping and then tests pairs of signals for colocalisation [9]. For each pair, coloc evaluates five hypotheses: no association with either trait (H0), association with the first or second trait only (H1 and H2), association with both traits through distinct causal variants (H3), or association with both traits through a shared causal variant (H4). Because our analyses focus on loci with significant associations for both traits, the principal comparison of interest is between H3 and H4.

Coloc probabilities are computed by combining lABF for all possible combinations of SNPs included in the analysis. H4 is estimated based on the elements of the diagonal of the resulting matrix, while H3 is estimated based on the off-diagonal elements. If the SNPs carrying most of the probability are the same, then probability will concentrate on the diagonal; vice-versa, if SNPs do not align, the probability will accumulate on the off diagonal elements and colocalisation will be unlikely. This representation of the problem highlights an important feature of colocalisation: if credible sets of the two traits we want to compare do not share any SNP, then colocalisation is impossible and thus there is no scope in testing it. FLANDERS uses overlap between coloc-sufficient credible sets to restrict testing to pairs that share at least one SNP. For each selected pair, colocalisation is tested using stored information from the coloc-sufficient credible set SNPs, while SNPs outside of coloc-sufficient sets are assigned the minimum lABF among its discarded variants.The same imputation approach is also applied to SNPs absent from that signal's original fine-mapping window when aligning the two variant sets. We refer to this approach as iColoc (imputed Coloc). It is conceptually similar to strategies implemented in Gentropy (https://github.com/opentargets/gentropy) where a discretionary small number is used instead and to what is done in Jesse et al., 2026 [10]. By retaining only the selected variant evidence and imputing the remainder at analysis time, iColoc greatly reduces storage requirements and decouples fine-mapping from colocalisation, allowing signals fine-mapped with different SNP windows to be compared.

#### Benchmarking of iColoc with traditional coloc

We benchmarked the proposed iColoc framework against a traditional SNP-level colocalisation approach using simulated genotype and phenotype data. Five genomic loci of 1.5Mb spanning a range of LD structures were selected for simulation (i.e. locus_1: chr2:133537-135037 kb, locus_2: chr3:186254-187754 kb, locus_3: chr11:125647-127147 kb, locus_4: chr14:64800-66300 kb and locus_5: chr19:5085-6585 kb). For each locus, two independent datasets were generated, each containing two causal variants. One causal variant was shared between the two datasets, representing a true colocalisation signal, whereas the second causal variant was unique to each dataset, representing dataset-specific association signals. Within each dataset, the two causal variants were selected to be in minimal linkage disequilibrium (LD; $r2 < 0.01$). Across datasets, the two dataset-specific causal variants were simulated under three LD scenarios, corresponding to low ($0 \leq r2 < 0.2$), moderate ($0.2 \leq r2 < 0.5$) and high ($0.5 \leq r2 < 0.7$) LD. This resulted in a total of 15 simulation scenarios (5 loci × 3 LD settings). Association summary statistics were generated from the simulated phenotypes and each region was fine-mapped using SuSiE. Fine-mapping results were stored as csAnnData objects using two alternative representations. The first retained lABF for all variants in the region and served as input for the traditional SNP-level colocalisation analysis. The second stored actual lABF values only the variants belonging to the coloc-sufficient credible sets, providing the input for iColoc. Traditional colocalisation analyses were performed for all possible pairwise combinations of credible sets, while iColoc analyses, according to the Flanders framework, were restricted to pairs of coloc-sufficient credible sets sharing at least one variant, thereby avoiding unnecessary pairwise comparisons between unrelated signals.

Across all 15 simulation scenarios, iColoc produced colocalisation posterior probabilities that very closely matched those obtained using the traditional SNP-level coloc approach (**Fig. S26**). Both methods correctly distinguished shared from dataset-specific causal variants with high accuracy (**Fig. S27)**. False-positive colocalisation calls occurred predominantly under the strongest LD scenario, where fine-mapping cannot reliably resolve independent causal variants in high LD. A notable advantage of iColoc was the reduction in the number of colocalisation tests performed: by considering only coloc-sufficient credible sets sharing at least one variant, iColoc avoided (N=2738, ~54%) comparisons that were consistently classified as true negatives (max $PP.H4=1.4x10^{-2}$) by the traditional method. Consequently, iColoc achieved equivalent benchmarking performance while halving the number of pairwise tests required, resulting in lower computational cost without loss of accuracy.

#### Signal-level multitrait colocalisation with SuSiE-HyPrColoc

Integrating molecular QTL and GWAS results across many traits requires identifying groups of association signals consistent with a shared causal variant. Although coloc evaluates two traits at a time, multitrait methods can assess evidence for sharing across larger groups. For example, moloc [11] extends the Bayesian coloc framework to multiple traits by evaluating configurations of shared and distinct causal variants, whereas mvSuSiE [12] jointly fine-maps multiple traits using a multivariate model that learns patterns of effect sharing. These approaches address related but distinct inference problems, and their computational requirements depend on the number of traits, variants and modelled signals. To organise our pairwise colocalisation results, we construct an undirected graph in which each node represents a fine-mapped association signal, identified by its credible set, and an edge connects two nodes when their pairwise colocalisation posterior probability satisfies PP.H4 > 0.75. Connected components provided candidate groups of shared signals for subsequent multitrait analysis. This representation also provides a framework for incorporating additional datasets while retaining existing pairwise results, although components affected by new nodes or edges may require reassessment.

A connected component does not, by itself, establish that all its signals share the same causal variant. Thresholded pairwise colocalisation results need not be transitive: evidence supporting colocalisation between A and B, and between B and C, does not necessarily imply sufficient evidence for colocalisation between A and C. Such patterns can arise from differences in statistical power, uncertainty in fine-mapping, or incomplete separation of nearby association signals. Consequently, neither a missing edge nor membership of the same connected component conclusively establishes whether signals have distinct or shared causal variants.

When the graph is not fully connected, it requires additional analysis to resolve, i.e., filling the missing connections and/or splitting the graph into several distinct graphs. One solution to resolve partially connected graphs is to use multitrait colocalisation approaches, such as moloc [11], which explicitly test the posterior probability of every possible combination of traits. While powerful, this strategy is computationally intensive and does not scale well as new data becomes available. We therefore used HyPrColoc [13] to assess multitrait sharing within candidate groups. HyPrColoc approximates the posterior probability that a group of traits shares a causal variant using regional association and alignment probabilities, and employs a branch-and-bound divisive clustering algorithm to identify supported subgroups. This avoids exhaustive enumeration of all possible sharing configurations and enables analysis of substantially larger numbers of traits. However, its underlying model assumes at most one causal variant per trait within the analysed region

To address this limitation, we combined SuSiE signal decomposition with HyPrColoc in a workflow termed SuSiE–HyPrColoc. Specifically, we supplied HyPrColoc with Z-scores reconstructed from the joint effect sizes and standard errors obtained from the iterative Bayesian stepwise selection (IBSS) step of SuSiE. Each input therefore represented an individual association signal rather than the full marginal association pattern of a trait. This adaptation enables the resolution of "colocalisation colliders" — cases where partial overlaps between signals artificially merge distinct causal effects — and allows us to disentangle subgroups of traits driven by different variants.

##### Joint fine-mapping

Once groups colocalised CS are defined using SuSiE-HyPrColoc, under the assumption that the underlying causal SNP is the same across all CS, we can integrate the fine-mapping information from each of them to estimate the probability of each SNP being causal to all involved traits. One approach is PIPmeta [14] which basically integrates the PIP values at each SNP:

$$\mathrm{PIP} = 1 - \prod_{k=1}^{K} (1 - \mathrm{PIP}_k)$$

This approach, although suitable, doesn't take into account the difference in strength of evidence coming from the different fine-mapping analyses. We thus instead integrate the evidence from multiple cs as:

$$lABF = \sum_{k=1}^{K} lABF_k$$

This is analogous to the formula used by coloc to estimate H4 or the probability that each SNP is causal to two different traits but extended to N traits. Given that we have already established that all credible sets in the group are all caused by the same SNP, the sum of lABF can be used as the joint log BF and thus can be used to retrieve the probability that each SNP is causal to all traits. This approach has also the advantage of using the lABFs coming from previously fine-mapping efforts while all other methods require reperforming fine-mapping from scratch every time a new phenotype is brought in.

To evaluate whether joint fine-mapping could improve the resolution of causal-variant identification across co-regulated signals, we used the previously described simulation framework. Within each simulated locus, signals showing evidence of colocalisation were grouped into modules regulated by the same causal SNP. Only modules containing more than one signal were considered for joint fine-mapping, as these provided the opportunity to combine information across multiple traits. Each eligible module was jointly fine-mapped as described to obtain a 99% joint credible set (JFM credible set). The JFM credible set for each module was therefore compared with the individual 99% credible sets obtained from standard fine-mapping of the signals included in that module (standard credible set). The comparison focused on three aspects of fine-mapping performance. First, we compared credible-set size, defined as the number of SNPs included in the 99% credible set, to assess whether joint fine-mapping reduced the number of candidate variants. Second, we compared the maximum posterior inclusion probability (PIP) within each credible set, as a measure of how strongly each approach prioritised its most likely causal variant. We also examined the PIP assigned specifically to the simulated causal SNP. Third, we assessed causal-variant recovery by calculating the proportion of simulations in which the SNP with the highest PIP corresponded to the true simulated causal SNP.

Across 335 paired comparisons, joint fine-mapping produced substantially smaller credible sets than standard fine-mapping (**Fig. S28a** and **S28b**). The median JFM credible set contained 3 SNPs, compared with 23 SNPs for the corresponding standard credible sets. This corresponded to a median reduction in credible-set size of 65.4%. Overall, the JFM credible set was shorter than the standard credible set in 84.8% of comparisons. Joint fine-mapping also assigned greater support to the simulated causal variant (**Fig. S28c**). The median PIP of the simulated causal SNP was 0.481 under JFM, compared with 0.102 under standard fine-mapping. The median absolute increase in simulated causal-SNP PIP was 0.243, and the causal SNP had a higher PIP under JFM in 95.5% of comparisons (**Fig. S28d**). The improvement in causal-variant prioritisation was also evident when considering the top-ranked SNP. Under standard fine-mapping, the SNP with the highest PIP corresponded to the simulated causal SNP in 33.7% of comparisons. In contrast, this occurred in 71.3% of JFM credible sets (**Fig. S28e**). Thus, joint fine-mapping increased recovery of the simulated causal SNP by 37.6 percentage points and more than doubled the proportion of comparisons in which the true causal SNP was ranked first. Overall, joint fine-mapping improved resolution (**i**) by reducing the number of candidate SNPs, (**ii**) increasing the posterior support assigned to the causal variant, and (**iii**) substantially improving the probability of correctly identifying the true causal SNP.

##### Software performance

Cell-type–specific cis-eQTL summary statistics comprised association results for 402,304 gene–cell type traits across 23 cell types and 4,452 individuals. These data were initially stored in 506 compressed tabular files totalling 178 GB, and were ingested into a TileDB array to support efficient storage and scalable access during fine-mapping and colocalisation. The resulting TileDB dataset occupied approximately 129 GB on disk, achieving a nearly 30% reduction in storage requirements. The FLANDERS workflow for fine-mapping and colocalisation analyses is implemented as a Nextflow pipeline and was executed on high-performance computing infrastructure, with up to four CPU cores and 64 GB of memory allocated per task. Data preprocessing, identification of association regions, fine-mapping and storage of the results in AnnData format required approximately 843 CPU-hours in total. The output comprised 114,063 credible sets, with credible-set and variant metadata and sparse matrices of log approximate Bayes factors (lABFs), effect estimates and standard errors, occupying approximately 2.2 GB.

#### Supplementary figures and legends

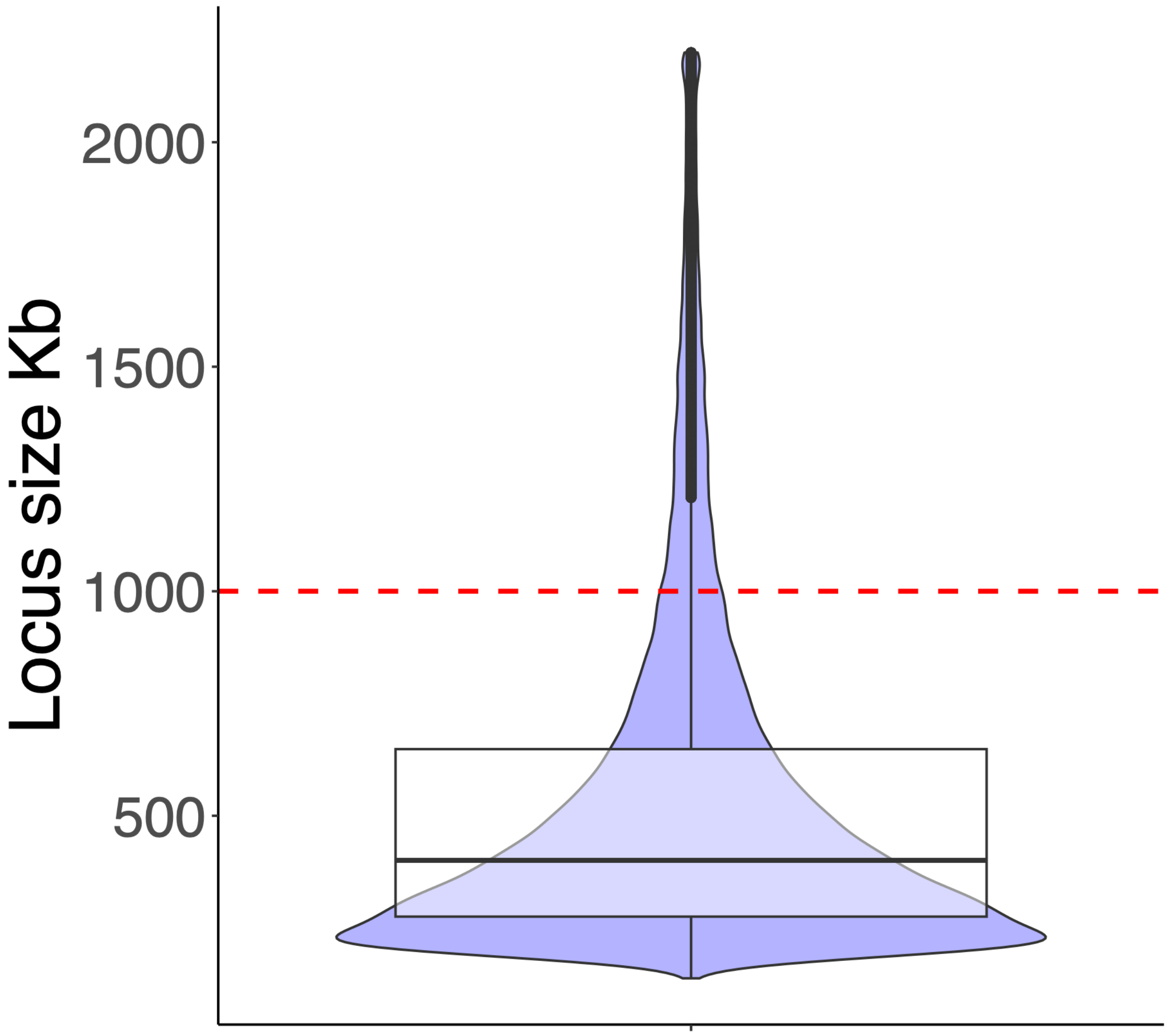

**Supplementary Figure S25 | Distribution of cis-eQTL locus sizes defined by Locusbreaker.**

Distribution of genomic region sizes after applying locus breaker to Cardinal cis-eQTL associations. The overlaid box plot indicates the median and interquartile range. The median locus size was 400 kb, with sizes ranging from 138 kb to 2.2 Mb. The red dashed line marks a locus size of 1 Mb.

**a**

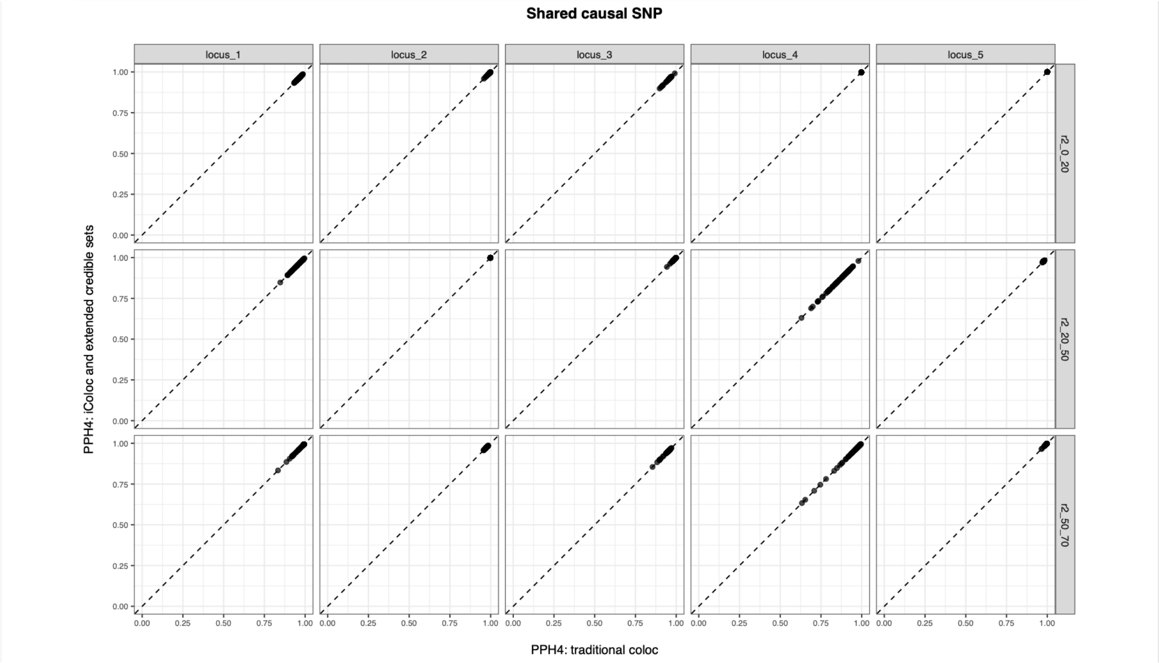

**b**

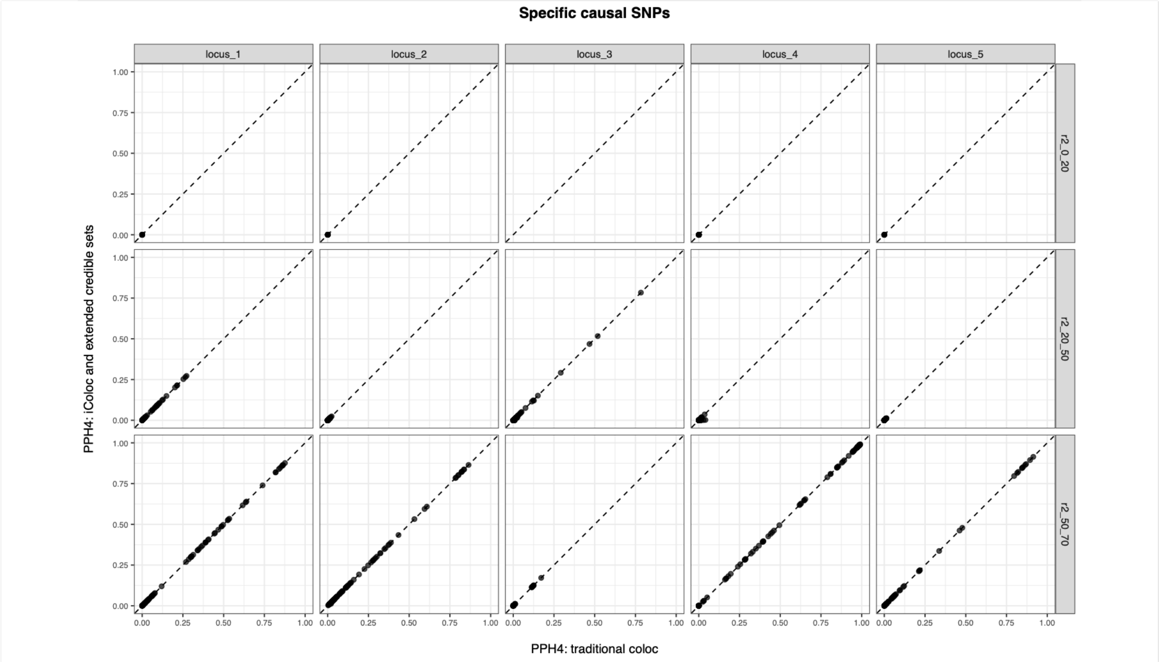

**Supplementary Figure S26. Comparison of colocalisation posterior probabilities (PPH4) obtained using the traditional SNP-level colocalisation approach and iColoc.**

Each point represents a matched colocalisation test performed by both methods. Panels correspond to the five simulated loci (columns) and the three linkage disequilibrium (LD) scenarios (rows). The dashed line indicates perfect agreement between methods. **(a)** Comparisons involving the shared causal variant. **(b)** Comparisons involving dataset-specific causal variants. Across all simulation scenarios, iColoc produced posterior probabilities that closely matched those obtained using the traditional coloc approach. Under the strongest LD scenarios ($0.5 \leq r2 < 0.7$), both methods occasionally supported colocalisation between dataset-specific signals, reflecting the reduced ability of fine-mapping to distinguish independent causal variants in strong LD.

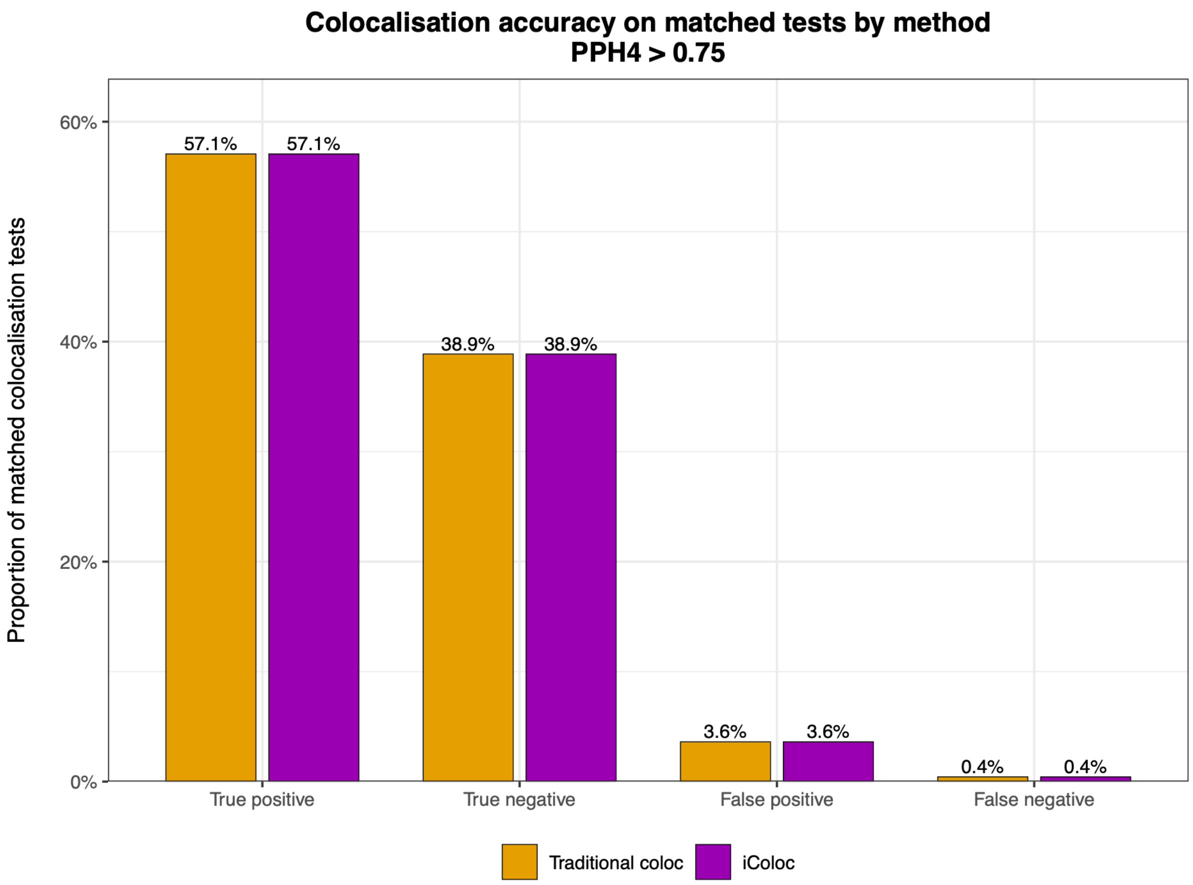

**Supplementary Figure S27. Comparison of colocalisation accuracy performance between traditional coloc and iColoc.**

Results are shown for the intersection of colocalisation tests performed by both methods. Positive colocalisation calls were defined as PPH4 > 0.75, and each comparison was classified relative to the simulated ground truth as a true positive, true negative, false positive or false negative. Both methods produced identical proportions of classification outcomes, indicating that iColoc preserves the accuracy of the traditional approach while evaluating fewer (~54%) colocalisation tests. The small proportion of false positive calls reflects limitations of fine-mapping in resolving highly correlated causal variants in the highest LD scenario.

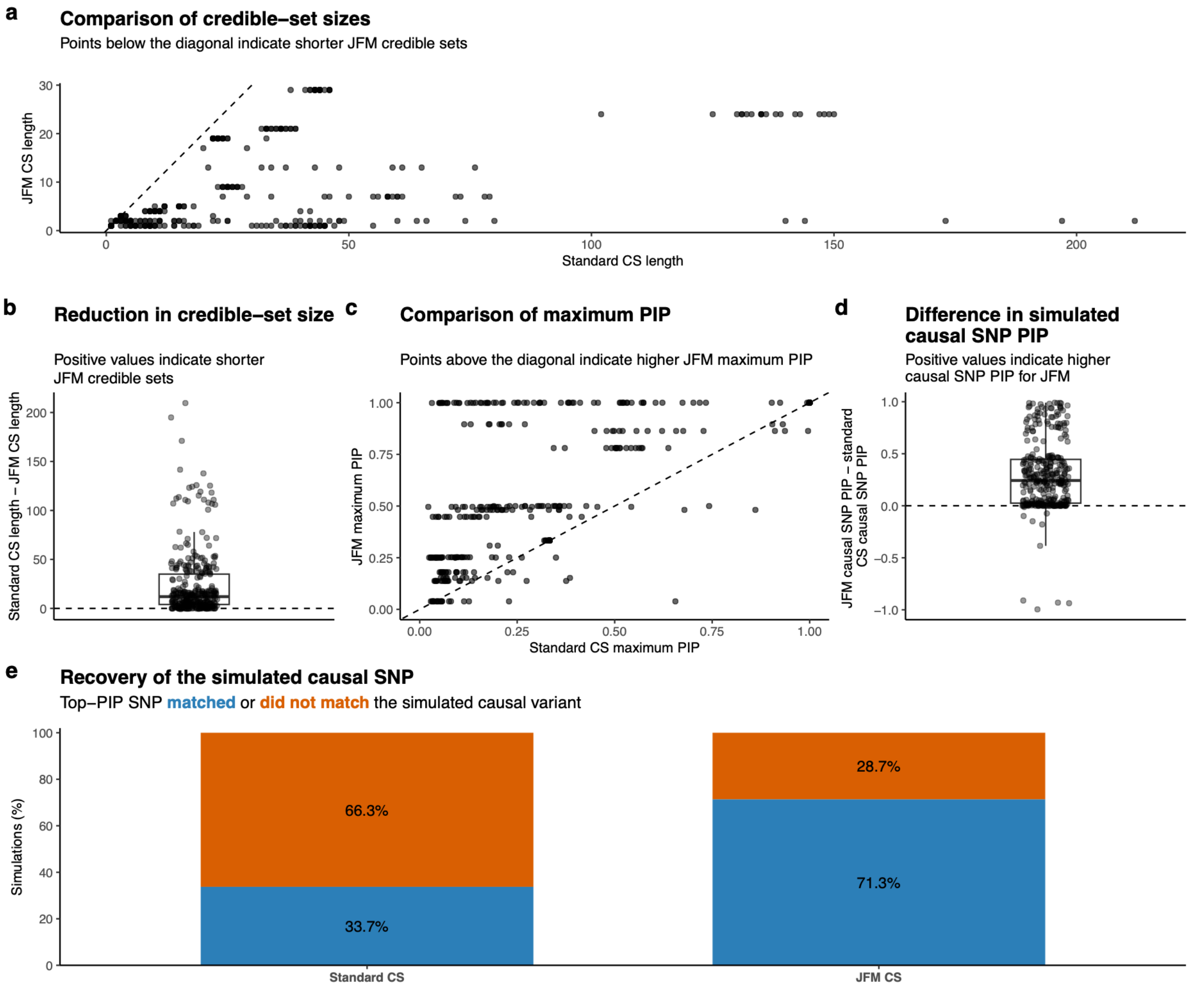

**Supplementary Figure S28. Joint fine-mapping improves credible-set resolution and causal-variant prioritisation.**

FLANDERS modules of co-regulated signals were jointly fine-mapped to derive 99% joint credible sets (JFM CS), which were compared with the corresponding 99% credible sets obtained from standard fine-mapping of the individual signals. **(a)** Comparison of credible-set size between standard and joint fine-mapping: points below the diagonal indicate shorter JFM credible sets. **(b)** Distribution of the difference in credible-set size, calculated as standard CS length minus JFM CS length: positive values indicate a reduction in the number of candidate SNPs with JFM. **(c)** Comparison of the maximum posterior inclusion probability (PIP) within each credible set: points above the diagonal indicate a higher maximum PIP under JFM. **(d)** Distribution of the difference in PIP assigned to the simulated causal SNP, calculated as JFM causal-SNP PIP minus standard causal-SNP PIP: positive values indicate greater posterior support for the causal SNP under JFM. **(e)** Proportion of simulations in which the SNP with the highest PIP matched the simulated causal variant.
